# Mapping global emergence of pathogens with epidemic and pandemic potential to inform and accelerate pandemic prevention, preparedness, readiness and response

**DOI:** 10.64898/2026.03.20.26347940

**Authors:** David M. Pigott, Huong T. Chu, Erin N. Hulland, Narmada Venkateswaran, Barbara A. Han, Adrian A. Castellanos, Oliver J. Brady, Ahyoung Lim, Diana P. Rojas, Sophie von Dobschuetz, Maria D. Van Kerkhove

## Abstract

**Introduction:** Increasing occurrence of epidemics and pandemics and concurrent emergence of different pathogens calls for multi-sectoral, multi-pathogen preparedness actions. Data on various factors that drive emergence of diverse pathogens can inform evidence-based preparedness by identifying geographies at-risk. When leveraging evidence within a One Health approach, multiple pathogens can be addressed simultaneously, thereby strengthening countries’ pandemic preparedness efforts.

**Methods:** For seventeen priority pathogens (avian influenza viruses, zoonotic coronaviruses including COVID-19, hemorrhagic fever viruses including Ebola, Henipaviruses, and arboviruses including yellow fever and Zika), we identified global evidence on animal reservoirs, vectors, environmental suitability, and reported human cases. We discriminated geospatially recorded pathogen detections from a background sample and constructed maps using these datasets to generate an evidence-based assessment of emergence risk globally.

**Results:** Seventeen pathogen-specific assessments were combined into a global composite map. Sub-Saharan Africa and South Asia have evidence supporting emergence risk for the greatest number of pathogens (included areas at-risk of all pathogens) and scored highest when strength-of-evidence weightings were factored. The Americas had the lowest tally of considered pathogens. Environmental suitability analyses received the highest weights, reservoir ranges the lowest.

**Discussion:** Preparedness and readiness must consider the range of global biological threats. Our methodology is capable of incorporating changing evidence on emergence potential for multiple pathogens to identify geographies at higher risk with different pathogen combinations. Our maps can contribute to existing decision-support structures, guiding shared interventions and strategic allocation of resources for spillover prevention and pandemic preparedness, thereby enhancing local response capacities applying a multidisciplinary approach.

**Research in Context:** *Evidence before this study:* Using PubMed, we searched for “[PATHOGEN] Preparedness Map” for each of seventeen priority pathogens to explore what resources might exist to be used to guide contemporary preparedness actions. The seventeen pathogens were: avian influenza viruses (AIV,all subtypes), chikungunya virus (CHIKV), Crimean-Congo hemorrhagic fever virus (CCHF), dengue virus (DENV), Ebola virus (EBV), Hendra virus, non-specific Henipaviruses, Lassa virus (LASV), Marburg virus (MARV), Middle East respiratory syndrome coronavirus (MERS-CoV), monkeypox virus (MPXV, all clades), Nipah virus, *Yersinia pestis*, Rift Valley fever virus (RVF), severe acute respiratory syndrome coronavirus 2 (SARS-CoV-2), yellow fever virus (YFV), and Zika virus (ZIKV). We also searched for “Emergence Preparedness Map” to try and identify a singular resource that housed all these pathogens. Searching for specific pathogens identified resources that had deployed specific approaches or types of data in answering this question, but often did not collate multiple varied evidence streams. Similarly, the more detailed resources tended to be more geographically restricted in scope. When searching for emergence resources more broadly, we identified some clusters of epidemiologically related pathogens being synthesized (for instance thinking about integrated management of vector-borne diseases), but none that spanned the full repertoire of pathogens listed. Others attempted to characterize the phenomena of emergence more broadly, but as a result lost the ability to further capitalize on pathogen-specific activities since pathogens were not a building block within a broader methodology.

*Added value of this study:* In evaluating the emergence potential for seventeen priority pathogens, we have collated the widest range of pathogens into a common map for synthesis. In doing so, we provide a support mechanism for actionable next steps for epidemic and pandemic preparedness at scale that leverage current knowledge. Contrasting to prior assessments, we leverage different types of data and provide a mechanism to differentially weight their inclusion. We outline a mechanism by which even for pathogens where comprehensive or detailed data is not present, the information currently available can be acknowledged and integrated, to provide immediate support for decision-making, while future enhancements are integrated when available and iterated upon. We also demonstrate how this modular methodology allows customized aggregations of pathogens where scopes of work necessitate - for instance, collating all pathogens with similar vectors where vector control actions can be undertaken. We show with examples of Marburg virus disease in Equatorial Guinea, how the maps demonstrated the prior evidence-base related to emergence of this disease in that geography and use that example to outline how these maps can indicate geographies of concern.

*Implications of all the available evidence:* Epidemic and pandemic preparedness is multi-faceted and multi-sectoral; some actions require pathogen specific insights, while other actions will work to counter a group of pathogens simultaneously. With this methodology, we demonstrate that it is possible to integrate data from diverse formats across different transmission routes and pathogen’s ecological dynamics globally to produce a set of resources to support local, regional, and global evidence-based decision making. Different groupings can be called upon to support different actions - pathogen specific maps where pathogen-specific vaccination schedules need to be undertaken; tracking the full pathogen-set that any given reservoir is implicated in; determining the differential diagnosis needs for a specific health facility and corresponding population it serves as a function of the implicated local pathogens or their potential future emergence; and supporting local health facilities in developing protocols, training, and necessary equipment to effectively detect and respond to possible local cases. Finally, these maps are designed to evolve alongside advancing infectious disease intelligence, allowing for continuous enhancement and resolution of data limitations across diverse surveillance systems and national contexts.

## Introduction

The COVID-19 pandemic demonstrated the massive health and economic impacts that emerging pathogens can have on society, both directly and indirectly, and has consequently renewed efforts to focus on preparing for any such future events. Concurrently, numerous epidemics of other “known” pathogens are occurring, whether mpox, itself resulting in two public health emergencies of international concern, highly pathogenic avian influenza H5N1 or Marburg virus disease (MVD) in Equatorial Guinea and Tanzania ^1–3^. These events have demonstrated that, even for pathogens that have emerged and circulated previously, we remain vulnerable to their re-emergence and enhanced transmission, particularly when such pathogens evolve, spread in previously unaffected communities (such as the men-who-have-sex-with-men community for mpox) or species (such as with the expansion of avian influenza H5N1 to land and marine mammals including dairy cattle) or in locations with no prior history of such pathogen identification (as with both mpox and the MVD cases).

The nature of our known biological threats is highly diverse whether viewed from the perspective of organisms responsible, reservoirs and vector species that sustain transmission in animals and or in humans, drivers of their emergence and spillover, including climatic factors, or transmission routes once spreading among humans, the symptoms and clinical progression associated with any resulting infections, or the health system capacities to detect and respond to such events, whether surveillance and diagnostic capacities, availability, access and affordability of medical countermeasures and policy options and implementation ^4–6^. Consequently, when assessing what the world needs to prepare for, governments must consider the multi-dimensional challenges when developing and sustaining support options involving multiple sectors. The COVID-19 pandemic emphasized the pre-existing heterogeneities and disparities across the world - whether differences in population-level comorbidity prevalence, demographic composition, existing health capacities, or in the ability to bolster existing resources or scale-up and deploy new interventions – especially those that were strengthened during past outbreaks/pandemics ^7,8^. With this in mind, a comprehensive effort to collate preemptive resources as a form of comprehensive, evidence-based and actionable health intelligence related to pathogen emergence is essential, particularly when paired with the need to take specific actions to prepare today, that align with the necessary timelines to implement these changes ^9^.

The Joint External Evaluation (JEE) and the State Party Self-Assessment Annual Reporting (SPAR) tool, developed to support the standards set by the International Health Regulations (IHR, 2005), represent the initial global systematic efforts to comprehensively evaluate national preparedness capacities for infectious hazards ^10^. These evaluations have been complemented by assessments from non-UN affiliated organizations ^11,12^. Collectively, these resources establish crucial baselines for preparedness activities and enable countries to monitor implementation progress through iterative assessments. Nonetheless, there remains a significant need for precise, quantitative data to effectively guide targeted pandemic preparedness and readiness actions, identify gaps in existing capabilities, and address specific vulnerabilities within national and global response systems. Similarly, under the Universal Health Coverage agenda, there remains an underexplored intersection between promoting health security for rare or potential biological threats including Disease X, and the ongoing improvements of the health of individuals day-to-day with the same package of actions ^13,14^.

Further, when considering specific known epidemic- and pandemic-prone pathogens, our understanding of the basic biology and epidemiology of these pathogens changes with more insights and information becoming available over time, and the context in which communities interact with these pathogens and their transmission routes is dynamically changing as a function of pathogen evolution, interventions implemented, climate change, or evolutionary drivers and selective pressures acting on that organism or affecting its surrounding animal-human-environment interface in a defined geographic space ^15,16^.

The increasing occurrence of epidemics and pandemics over the last several decades and the concurrent emergence or re-emergence of different pathogens calls for multi-pathogen prevention and preparedness approaches. WHO and others have identified the need to generate global composite maps that combine information of different pathogens with epidemic and pandemic potential in order to optimize decision-making on local preparedness activities, building upon and learning from disease specific maps and models already available ^17,18^. The consolidation of the latest knowledge, a clear understanding of current gaps in this knowledge, and the critical use of existing resources in making real time decisions is a necessary part of the preparedness landscape. For many pathogens with epidemic and pandemic potential, the scientific community is in the intelligence gathering phase as we have limited knowledge on factors driving emergence or spillover, and the breadth of vectors or reservoir species. For other pathogens, we have better global estimates of the populations affected, their historical exposure, and existing health system capacities to detect and treat as well as population level vulnerabilities, and as a result can be more confident in what prevention and response actions to take where. It is crucial to acknowledge the diversity of biological threats the world is facing today and consider multi-sectoral, multi-pathogen in addition to pathogen-specific approaches to decision making in epidemic and pandemic preparedness efforts.

Our main objective was to build on existing mapping exercises to generate updated evidence-based risk maps that demonstrate the value in integrating varied evidence types for 17 priority pathogens with epidemic and pandemic potential, differentiating their ability to discriminate areas at-risk, and prompting allocation of resources to existing approaches and decision-support structures for zoonotic spillover prevention and pandemic preparedness. We aimed to construct a methodology that is not only adaptable to dynamic environmental conditions but also responsive to emerging scientific insights, evolving environmental conditions like climate change and capable of integrating novel forms of intelligence such as those derived from the detection and characterization of newly emerging pathogens (or disease X), the identification of new reservoir species, or novel model-based evaluations of specific risk dimensions. By considering a wide array of pathogens known to have epidemic or pandemic potential, these results demonstrate the opportunity space for synergistic actions, whether focusing on shared interventions such as considering principles aligned with collaborative surveillance, use of multi-pathogen diagnostic panels or integrated pest management or vector control, shared health system stressors, shared population health vulnerabilities, shared animal reservoirs and shared risks.

## Methods

### Pathogen selection and dataset collation

Using existing prioritization exercises including the WHO Blueprint pathogen prioritization effort (updated 2017)^19^, knowledge of available global resources, and iterating with World Health Organization technical teams, we aimed to select a set of critical pathogens that not only have epidemic and pandemic potential ^20^, but also are diverse enough to demonstrate how the subsequent methodology and maps could be used to evaluate pathogens with different epidemiological traits, ecological contexts, and taxonomic backgrounds. The following pathogens were included in our analysis: avian influenza virus (AIV, all subtypes), chikungunya virus (CHIKV), Crimean-Congo hemorrhagic fever virus (CCHF), dengue virus (DENV), Ebola virus (EBV), Hendra virus, non-specific Henipaviruses, Lassa virus (LASV), Marburg virus (MARV), Middle East respiratory syndrome coronavirus (MERS-CoV), monkeypox virus (MPXV, all clades), Nipah virus, *Yersinia pestis*, Rift Valley fever virus (RVF), severe acute respiratory syndrome coronavirus 2 (SARS-CoV-2), yellow fever virus (YFV), and Zika virus (ZIKV). The datasets include a diversity of pathogens, including those with bird or mammal reservoirs, transmission routes as varied as vector-borne, blood-borne or respiratory, and differing causal pathogen biology including viruses and a bacterium (*Yersinia pestis*).

For each included pathogen, we identified and analyzed existing globally comprehensive assessments for pathogen occurrence or environmental suitability that considered a variety of different evidence types. We aimed to compile relevant and available data at continental or global scales, focusing on the following variables where applicable: (i) countries or first administrative divisions where human cases have been reported to United Nations-level organizations or other open sources of epidemiological data, (ii) distributional data related to vector species and animal hosts, (iii) environmental suitability (modelling that assesses the relationship of covariates including temperature, precipitation, normalized difference vegetation index (NDVI), enhanced vegetation index (EVI), elevation/altitude and other environmental variables with pathogen detections) assessments delimiting populations-at-risk for exposure to the pathogen (iv) geopositioned detections of pathogens in human and animal sources through published research.

Countries and regions reporting human cases infected with the pathogen were sourced from the Food and Agriculture Organization of the United Nations’ EMPRES-I databases, supplemented where relevant, including results of molecular and serologic field published studies. For mammals and birds identified as possible reservoir hosts, we differentiated the strength of the evidence supporting these assumptions as ranging from detection of infection with the pathogen of interest in any context to a detected infection in a non-experimentally induced context or in a wild animal, considering a detected infection as determined by Reverse Transcription-Polymerase Chain Reaction (RT-PCR), genotyping, or other advanced, non-serological assay or technique. We used range maps as outlined by BirdLife International or the International Union for Conservation of Nature (IUCN) ^21,22^ to define spatial ranges of the implicated wild animals. For vector distribution we incorporated a model-based estimate of vector presence/absence. We identified relevant environmental suitability assessments and geopositioned records of pathogen detections from published literature or governmental resources. Details of the specific evidence used, their provenance and global distribution, and additional evidence-specific processing are outlined for each pathogen in the Supplementary Information.

Each evidence base was converted into a binary measure, either since the data was already categorical (e.g. a country has reported a case, or it has not; a location has the home range of one of a list of susceptible species, or it does not) or by applying a threshold value (e.g. applying consistent thresholds of 0.2, 0.4, 0.6, 0.8 to all environmental suitability indices identified, or a threshold that characterizes a specific percentile of all elements). All evidence was summarized at a 5km x 5km raster resolution, matching the most granular resolution of included inputs.

### Evidence synthesis and aggregation

Once evidence was compiled and processed, each element was compared to the spatially annotated database of reported occurrences of the pathogen of interest, to determine the precision value of such evidence base. Ultimately, such evidence aims to guide decisions and in-country operations by quantifying the ability of each to discriminate between locations where such pathogens have been reported, compared to a baseline set of control locations. For this assessment, a random sample was taken uniformly from across the world to provide these details which served as a common reference baseline used for each pathogen. Within such a methodology, an evidence-base is penalized for having too broad a characterization of risk (i.e. too many locations where there have been no instances of the pathogen are included). For plague and SARS-CoV-2 where no global geopositioned database of disease occurrences were identified, the mean value of such evidence type in all other pathogens was used instead. For each individual pathogen, a global pathogen-specific map was produced by taking, for any given location, the sum of each binary evidence-base multiplied by the precision weight of that evidence type.

Given that the maximum value possible for a specific pathogen was a function of the number of evidence-types included, prior to aggregating all pathogen-specific maps into a multi-pathogen composite map, we standardized each layer so that they would all have the same maximum value, 1. To do this, each layer was divided by the maximum possible value, the tally of the total types of evidence considered. With this methodology, therefore, pathogens with inconsistent evidence will have a lower score than those with consistent evidence; similarly those where the only used evidence is weak (as determined by their precision score) will have a lower score than a pathogen system where the used evidence is strong.

## Results

Our results demonstrate the complex mosaic of spillover risk that critical epidemic-prone pathogens represent across the globe. Our results suggest that populations in all countries may be exposed to at least one of the pathogens considered. In some territories, the pathogen profile is varied and needs a varied suite of actions to prevent and prepare.

### Geographic range of emergence risk for epidemic- and pandemic-prone pathogens

When considering the risk for individual pathogens derived from the synthesis of all pathogen-specific inputs, SARS-CoV-2 and avian influenza virus were widespread in their geographic risk distributions as compared to other pathogens (Supplementary Appendix). Underpinning this broad risk profile is the number of unique species (44 and 303) that have been documented as having been infected by SARS-CoV-2 and HPAI strains, respectively. In contrast, our results demonstrate that MERS-CoV has the smallest geographic risk range of any of the pathogens considered, primarily focused within the Arabian Peninsula, and also spanning North Africa, the Horn of Africa, and as far west as the Indian subcontinent.

When evidence was overlaid for multiple pathogens in a composite map, our results demonstrate that the greatest potential for emergence of the largest number of pathogens are in locations in sub-Saharan Africa. Our results provide evidence that some areas of South Sudan, for example, are at risk of exposure to emergence of all priority pathogens considered (Figure 1). Large geographic areas in Western Africa, including Nigeria and Ghana, in Central and in Southern Africa (from Gabon and Cameroon, to the United Republic of Tanzania and Mozambique) similarly have areas with evidence to support emergence potential for the different pathogens evaluated, with important foci for Lassa virus (LASV), *Yersinia pestis*, monkeypox virus (MPXV), Rift Valley fever virus (RVF), Ebola virus (EBV), and Marburg virus (MARV) present among others.

**Figure 1:**
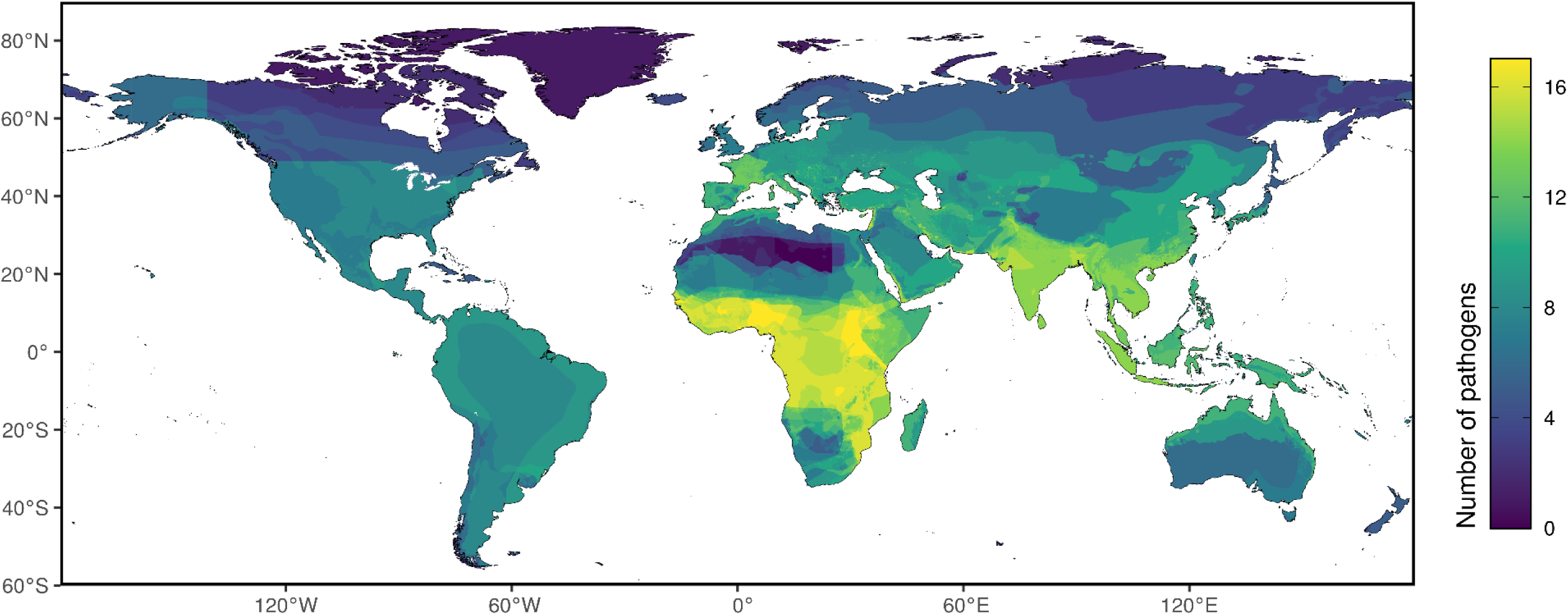
Global distribution of the number of pathogens with any evidence to support their presence in a given location. Areas in yellow have the highest count of pathogens while areas in dark blue have the fewest.

Outside of Africa, in South Asia our results illustrate support for the emergence potential for the highest numbers of pathogens in Western India, specifically Gujarat and Rajasthan, in South-East Asia, in the Mekong Delta (Cambodia, Lao People’s Democratic Republic, and Viet Nam), Indonesia, and in southern China where there is evidence of emergence potential of a multitude of pathogens, including key foci for DENV, non specific Henipaviruses, CHIKV, and ZIKV. In Europe, the highest risk for emergence potential was identified in France and the Balkan Peninsula, with again approximately a dozen implicated pathogens, such as CCHF. The Americas was the region with the lowest risk for the considered pathogens, with parts of Northern Canada having some of the lowest number of pathogens at risk of emergence with only one or two, often SARS-CoV-2 and HxNx avian influenza, priority pathogens with any evidence to support that these populations were at risk.

While the macroecological patterns of many of these pathogens are relatively widely known, we are also able to reflect the subnational heterogeneity in possible pathogen exposure. Our results show strong signals in variation across northern nations such as Canada, Sweden, and the Russian Federation, where the southernmost latitudes had evidence to support potential exposure to more pathogens than others. In other countries, such as Nigeria and Australia, as we look at the more arid regions of these countries, the number of implicated pathogens decline.

Figure 2 explores how, once we factor in the predictive power of different forms of evidence, we see a further detailing of the spatial variation in possible emergence potential. Countries such as Guinea, Sierra Leone, and Liberia have the highest composite score considering the differential weighting of each evidence type of each pathogen. In Ghana, Nigeria, Cameroon, and Gabon, while all having evidence to support at-risk status for emergence of multiple pathogens, there are heightened foci at sub-national levels, in specific provinces and regions, when factoring in the strength of the collective evidence. Subnational detailing of differential exposure is heightened when poorly discriminating evidence types are downweighed in importance. We see a reversal in status of countries in the Horn of Africa however; South Sudan which had the highest number of pathogens present, when the evidence to support that status was recalibrated to account for the differential strength of the local evidence a composite score lower than that of other continental peers resulted. The Indian subcontinent across to southern China remain the areas of Asia with the highest aggregate risk. In Europe and North America we see a far larger reduction in weighted composite risk than when looking at the binary status of any evidence to support possible pathogen exposure – in the United States of America in particular, subnational variation becomes more pronounced, with the southern states such as Texas and Florida remaining at more elevated risk as compared to Northern and Western counterparts. In South America, we see that while the number of pathogens present was less, their risk status is underpinned by comparatively robust data – Brazil, Colombia, and the Bolivarian Republic of Venezuela all have a higher combined score than all of Europe and North America, while in pathogen tally space shown in Figure 1, these latter regions were higher.

**Figure 2:**
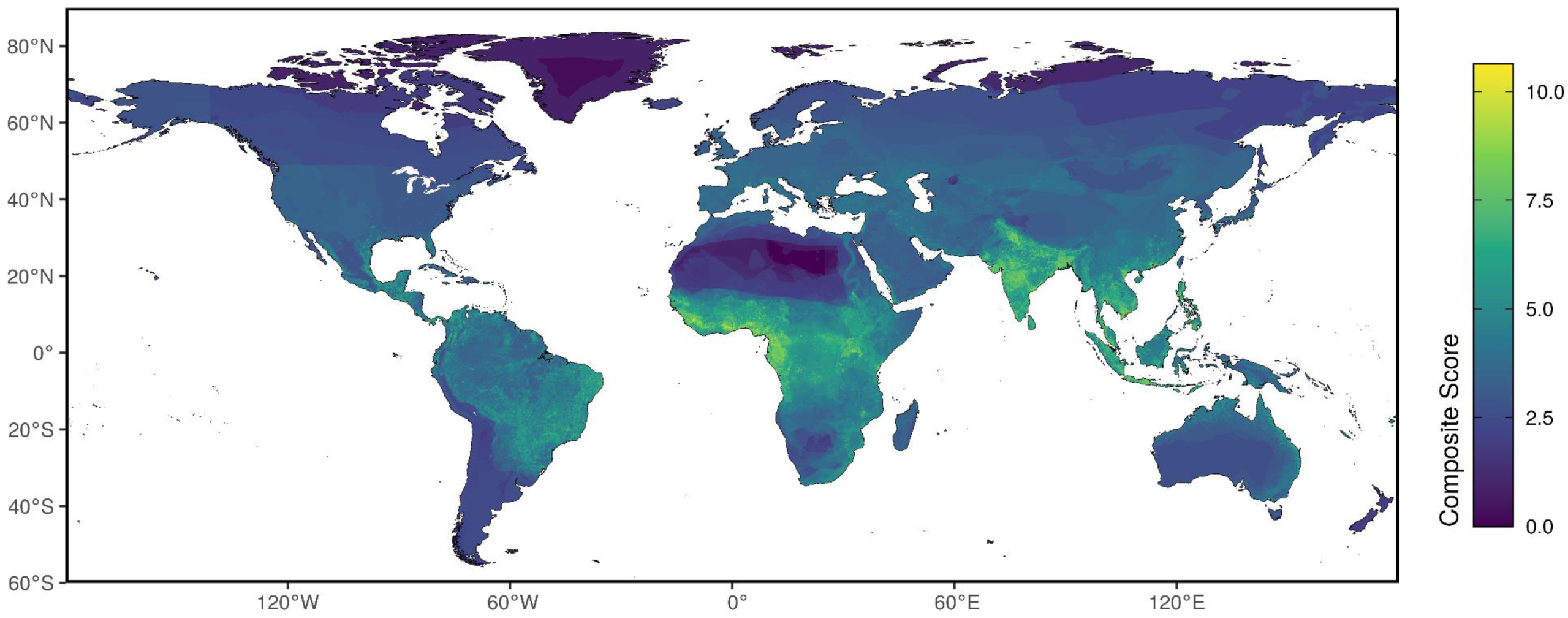
Final composite score combining standardized pathogen-specific evidence scores. Areas in yellow have the highest aggregate score while areas in dark blue have the lowest.

### Evaluating critical groupings of related pathogens

By virtue of evaluating risks for such a breadth of pathogens, we can provide insights into the global distribution of various critical groupings, whether aligning with aspects relevant to their surveillance, localizing interventions, or how different populations and health systems may prepare or respond. Figure 3a builds upon recent work ^18^, and collates the pathogens relevant to *Aedes*-borne transmission, namely dengue, chikungunya, Zika, and yellow fever, where we see highest scores in tropical climates. Many regions have risk for all four pathogens (Supplementary Appendix) however, we do see a broader area of risk north and south of the equator related to the presence of the *Aedes* mosquito outside of locations where the pathogens have been detected. Figure 3b demonstrates the areas at risk of rodent-borne pathogens, plague, mpox, and Lassa fever. Here we see an emphasis in West and Equatorial Africa, where the presence of all three pathogens is supported by multiple evidence types. Given the essential role that rodent hosts play for pathogen maintenance and spillover into humans and considering their ubiquitous presence, we see far broader areas of risk identified across the globe, with no-zero composite scores for most nations. In particular, a large number of the rodent hosts are implicated in plague transmission, and therefore possible population-level risk is present in North America, South America, and Europe and North Asia – in many of these regions, plague cases have also been reported previously.

**Figure 3:**
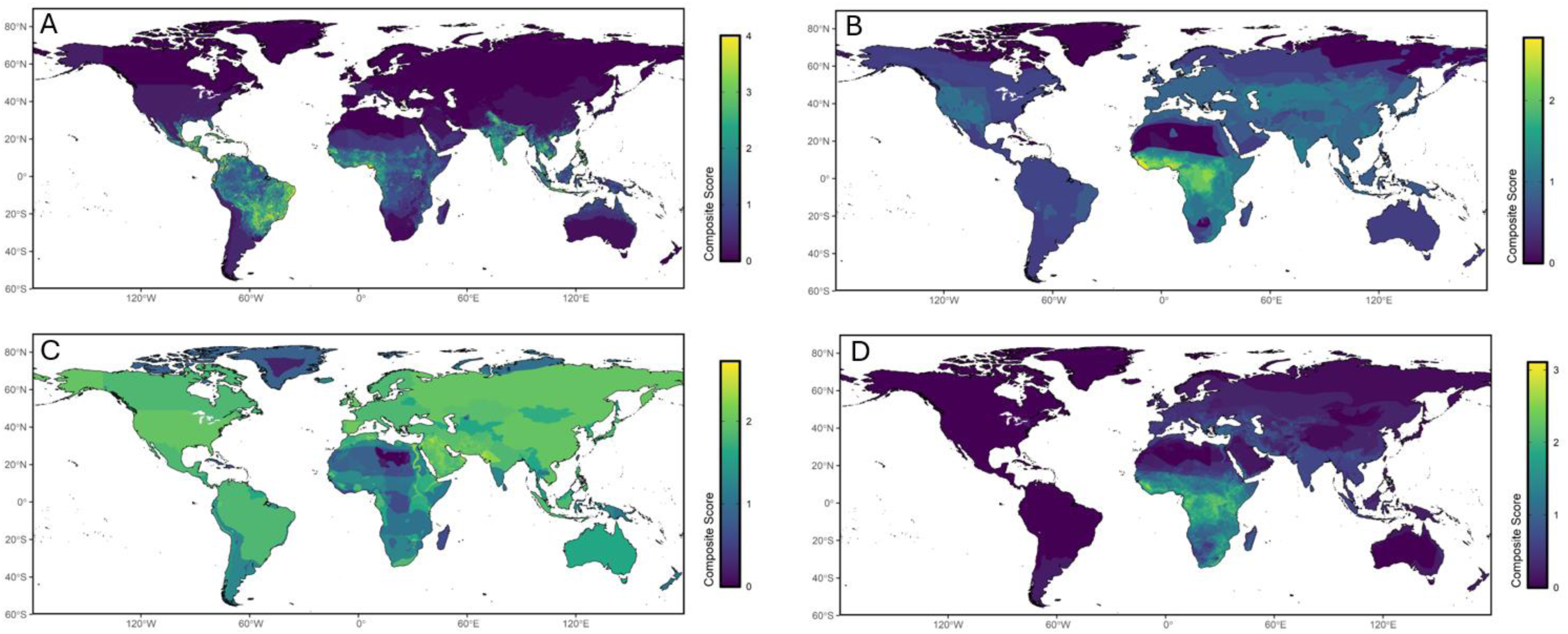
Composite scores for specific collections of pathogens related to (a) Aedes-borne pathogens (dengue virus (DENV), chikungunya virus (CHIKV), yellow fever virus (YFV), and Zika virus (ZIKV)) (b) rodent-borne pathogens (monkeypox virus (MPXV, all clades), *Yersinia pestis*, and Lassa virus (LASV)) (c) respiratory pathogens (Middle East respiratory syndrome coronavirus (MERS-CoV), avian influenza viruses (AIV, all subtypes), and severe acute respiratory syndrome coronavirus 2 (SARS-CoV-2)) and (d) hemorrhagic fevers (Crimean-Congo hemorrhagic fever virus (CCHF), Ebola virus (EBV), Marburg virus (MARV), Rift Valley fever virus (RVF) and LASV).

Figures 3c and 3d reflect characteristics of the pathogens when circulating within human populations, versus traits associated with the broader life-history of the pathogen. Figure 3c includes respiratory pathogens (MERS-CoV, avian influenza, and SARS-CoV-2) where initial symptoms in humans are indistinguishable and human-to-human transmission occurs in a similar manner, and therefore healthcare workers need to consider similar containment strategies in triaging and treating suspected or confirmed patients. These pathogens are present across the globe with most nations having a similar composite scoring. The broad range of mammalian and avian hosts as noted above results in a near global risk profile for this grouping. Figure 3d looks at another cluster of pathogens with shared clinical syndromes, the hemorrhagic fevers, where again it is important for healthcare workers to implement high quality infection prevention and control strategies when treating suspected and confirmed patients. Here, the emergence potential for Ebola, Rift Valley fever, Crimean-Congo haemorrhagic fever, Marburg, and Lassa fever, are focused in the sub-Saharan African Region, while parts of Eurasia have non-zero risk scores due to the presences of several mammalian hosts having their home range also in these territories.

### Robustness in various forms of evidence

We have seen how weighting different evidence types by their precision scores changes the global composition from Figure 1 to Figure 2. Table 1 quantifies the different scores present among the various evidence types and pathogens. In general, we found that the environmental suitability layers provided the greatest discriminatory power among the data types considered. For all pathogens where an environmental suitability assessment was sourced, with the exception of mpox, this evidence type received the highest weighting. The most stringent threshold version of the suitability layers received the highest average weight across all the priority pathogens (mean of 0.8356). In contrast, the reservoir range layers generally received the lowest weightings; the broadest reservoir definition category of any type of reported infection among animals received a mean weighting of 0.29689 across all the pathogens. Using prior cases (which for avian influenza and MERS-CoV was differentiated to be among humans and animals separately), received the most inconsistent weighting when comparing across the pathogens, ranging from 0.2279 for chikungunya virus to 0.8442 for MERS-CoV.

**Table 1:**
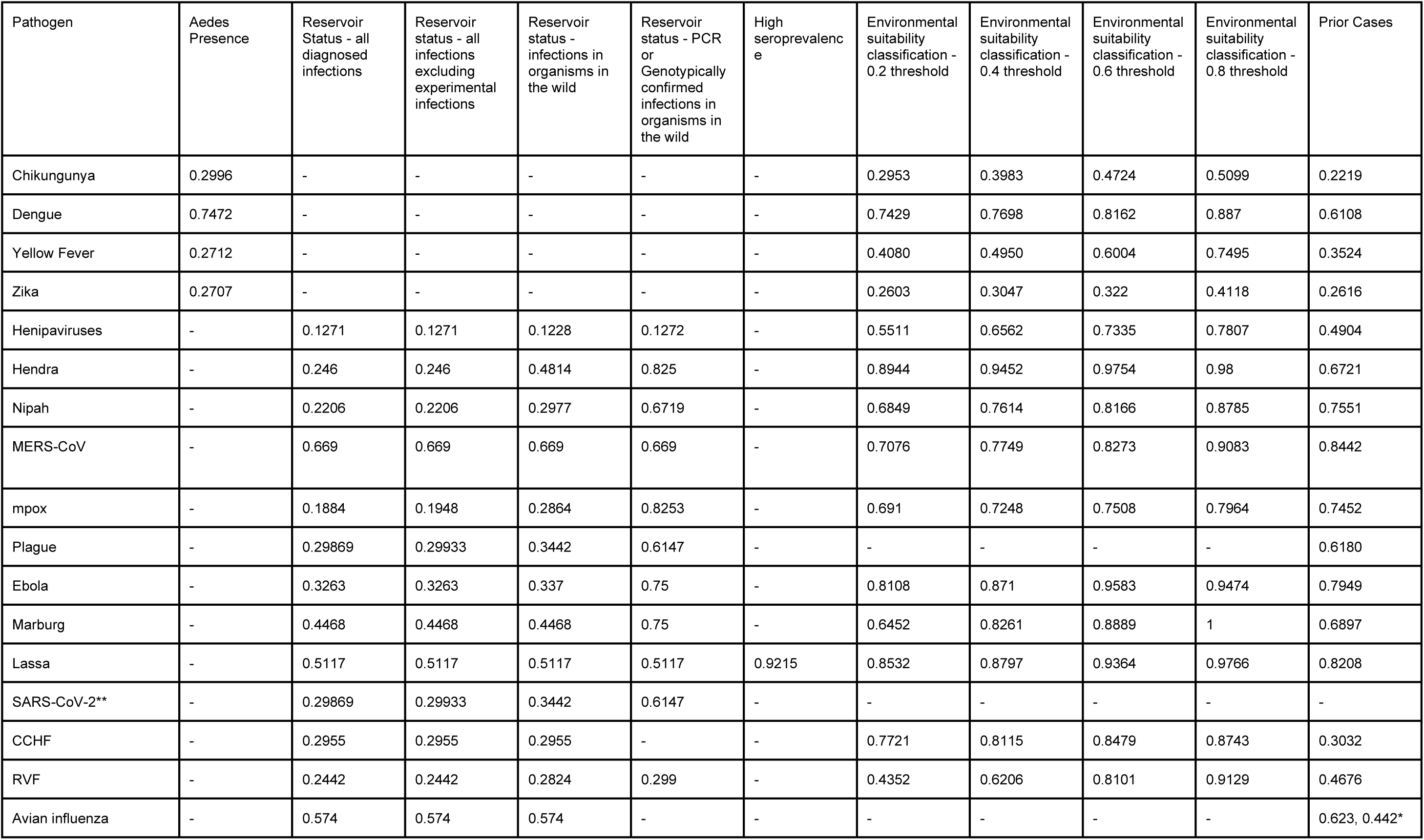
Evidence types used for specific pathogens and corresponding precision weights. - denotes evidence type not incorporated in this iteration. *avian influenza had two different prior case datasets - one for reported animal infections, the other for reported human cases. ** Precision weights for these pathogens are derived as the mean of all other pathogens’ precision weights for the specific evidence type.

| Pathogen | Aedes Presence | Reservoir Status - all diagnosed infections | Reservoir status - all infections excluding experimental infections | Reservoir status - infections in organisms in the wild | Reservoir status - PCR or Genotypically confirmed infections in organisms in the wild | High seroprevalence | Environmental suitability classification - 0.2 threshold | Environmental suitability classification - 0.4 threshold | Environmental suitability classification - 0.6 threshold | Environmental suitability classification - 0.8 threshold | Prior Cases |
| --- | --- | --- | --- | --- | --- | --- | --- | --- | --- | --- | --- |
| Chikungunya | 0.2996 | - | - | - | - | - | 0.2953 | 0.3983 | 0.4724 | 0.5099 | 0.2219 |
| Dengue | 0.7472 | - | - | - | - | - | 0.7429 | 0.7698 | 0.8162 | 0.887 | 0.6108 |
| Yellow Fever | 0.2712 | - | - | - | - | - | 0.4080 | 0.4950 | 0.6004 | 0.7495 | 0.3524 |
| Zika | 0.2707 | - | - | - | - | - | 0.2603 | 0.3047 | 0.322 | 0.4118 | 0.2616 |
| Henipaviruses | - | 0.1271 | 0.1271 | 0.1228 | 0.1272 | - | 0.5511 | 0.6562 | 0.7335 | 0.7807 | 0.4904 |
| Hendra | - | 0.246 | 0.246 | 0.4814 | 0.825 | - | 0.8944 | 0.9452 | 0.9754 | 0.98 | 0.6721 |
| Nipah | - | 0.2206 | 0.2206 | 0.2977 | 0.6719 | - | 0.6849 | 0.7614 | 0.8166 | 0.8785 | 0.7551 |
| MERS-CoV | - | 0.669 | 0.669 | 0.669 | 0.669 | - | 0.7076 | 0.7749 | 0.8273 | 0.9083 | 0.8442 |
| mpox | - | 0.1884 | 0.1948 | 0.2864 | 0.8253 | - | 0.691 | 0.7248 | 0.7508 | 0.7964 | 0.7452 |
| Plague | - | 0.29869 | 0.29933 | 0.3442 | 0.6147 | - | - | - | - | - | 0.6180 |
| Ebola | - | 0.3263 | 0.3263 | 0.337 | 0.75 | - | 0.8108 | 0.871 | 0.9583 | 0.9474 | 0.7949 |
| Marburg | - | 0.4468 | 0.4468 | 0.4468 | 0.75 | - | 0.6452 | 0.8261 | 0.8889 | 1 | 0.6897 |
| Lassa | - | 0.5117 | 0.5117 | 0.5117 | 0.5117 | 0.9215 | 0.8532 | 0.8797 | 0.9364 | 0.9766 | 0.8208 |
| SARS-CoV-2** | - | 0.29869 | 0.29933 | 0.3442 | 0.6147 | - | - | - | - | - | - |
| CCHF | - | 0.2955 | 0.2955 | 0.2955 | - | - | 0.7721 | 0.8115 | 0.8479 | 0.8743 | 0.3032 |
| RVF | - | 0.2442 | 0.2442 | 0.2824 | 0.299 | - | 0.4352 | 0.6206 | 0.8101 | 0.9129 | 0.4676 |
| Avian influenza | - | 0.574 | 0.574 | 0.574 | - | - | - | - | - | - | 0.623, 0.442* |

### Real-time validation of the methodology

During the development of these maps, two countries (Equatorial Guinea and the United Republic of Tanzania) reported their first cases of MVD. Despite the lack of prior human cases, both countries, as evidenced in this study, had multiple types of evidence to support a non-zero risk status. Both Kientem, Equatorial Guinea, and the Kagera Region of Tanzania are homes to bats implicated as reservoirs for the virus, inclusive of the Egyptian fruit bat, *Rousettus aegyptiacus,* which has had PCR detections of the virus. Similarly, these regions are present in the 0.2, 0.4 and 0.6 thresholded environmental suitability maps (Figure 4); indeed Kientem was named as a region for further surveillance for the virus as far back as 2015 ^23^. Thinking about other similar regions, we can leverage our maps to identify locations that have a comparable, or higher, score than Kientem (Kagera already had a higher MARV specific score) and have not already seen outbreaks of the virus (Figure 4). In doing so, we identify areas of Sierra Leone, Liberia, Cameroon, Gabon, and Central African Republic, as well as Zambia and Ethiopia (Figure 4).

**Figure 4:**
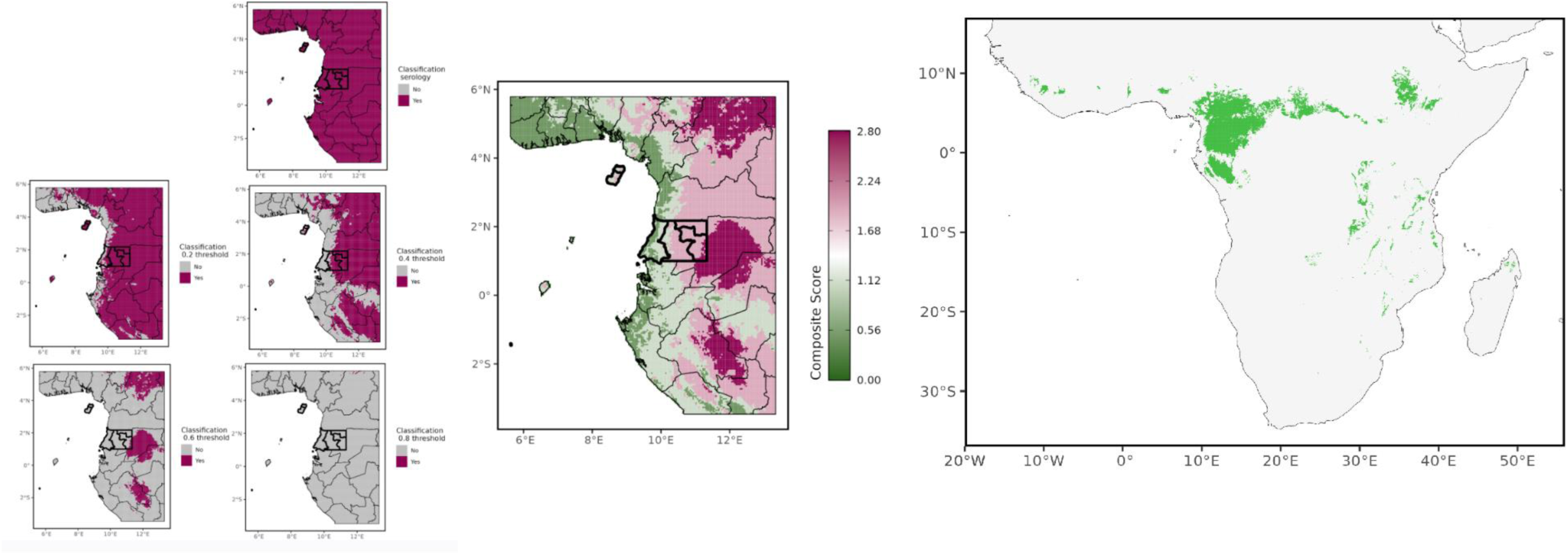
Marburg virus (MARV) assessment in the Equatorial Guinea context. On the left hand side, areas in purple denote regions where the evidence type supports potential risk status, including reservoir bat species, and the four different environmental suitability assessments. The central map shows the composite weighted score for MARV after weighting each data type by their precision value. In both the left and central panels Equatorial Guinea is denoted with thicker black borders. The rightmost panel shows all other regions of the world that have comparable or higher MARV-specific composite scores than Kientem, Equatorial Guinea.

## Discussion

Our results show significant global variation in emergence potential of priority pathogens as indicated by a diverse set of evidence. Pandemic preparedness activities must factor in current known evidence to provide support to those populations most likely to be impacted by pathogen emergence. We have shown that a variety of data exists that can be leveraged today to continue to develop national preparedness plans and target activities ^9,24,25^ to reflect geographic vulnerability to a range of different pathogens and provide an evidence-based foundation to build-upon with additional data and analysis. Importantly, previous pathogen-specific assessments inadvertently perpetuate the false assumption that countries need to only prepare for those pathogens with which they have experienced outbreaks in the past. When a broader spectrum of evidence is considered, particularly the geographic distribution of animal hosts and vectors involved in disease transmission, it becomes clear that the potential footprint for disease emergence extends far beyond areas with previously reported cases. While it is fair to say that these broader forms of evidence are less predictive (as evidenced by their weighting in this study), they are nonetheless greater than zero and can be used to advocate for strengthening local capacities in countries at risk. The Marburg experiences in Equatorial Guinea and the United Republic of Tanzania serve as a reminder of this phenomena, where the maps in this paper highlighted the elevated risk these regions had compared to others, and provided preemptive evidence to support ensuring local capacity was capable of this pathogen’s detection. As outlined in Figure 4, countries and regions can begin to consider risk as an equivalency calculation, identifying communities that have a comparable epidemiological and ecological context to regions that have seen cases, but to-date may have not seen outbreaks arise due to the overall relative rarity of some of these pathogens. They can consider the different state-of-the-art in terms of evidence and make decisions accordingly as to whether to proceed with further strengthening based upon this, or in the context of many other pathogens.

These maps represent a baseline for taking targeted key actions to mitigate not only individual pathogens, where the specific countermeasures may be relevant (such as using the Ebola specific resources in the Supplemental Appendix to stockpile Ebola virus vaccines in anticipation of a future outbreak ^26^), but also multiple pathogens (such as protecting cold chain systems to ensure delivery of any type of vaccine when needed ^27^, or building trust and awareness among communities indicated as being at-risk ^28^). Actions taken to address vaccine delivery for a possible Ebola outbreak will also benefit yellow fever, chikungunya, mpox, or yet-to-be-developed vaccines ^29^, and as a consequence, combining insights from potential spillover for all these pathogens simultaneously can indicate where maximal impact could be achieved. Such decisions can occur not only in global planning activities, where the constellation of microbial threats in each country can be considered, but also within countries where state-level, or community-level compilations of evidence can be reviewed. All of these can be evaluated in the context of additional population health outcomes or health-system level resources, such as current vaccine-coverage for childhood diseases, or health facility assessments of existing storage capabilities.

Such pluralistic approaches can benefit a variety of activities: vector control efforts to address pathogens with vectors in common, such as *Aedes* mosquitoes and their roles in CHIKV, DENV, YFV and ZIKV transmission, can utilize these resources to identify regions where all four are present (Figure 3a). Animal surveillance activities can leverage these geographically resolved resources to identify for given surveys sites, which pathogens could have the potential to be circulating among animals they may encounter or if focused on the capture of a specific animal, they can cross-reference the ledgers of reservoir status in order to determine which serological assays or PCR primers to run tests upon given the current evidence to support reservoir status ^30^ (Figure 3b). Human surveillance efforts can benefit from considering these resources, whether ensuring that laboratory systems are sufficiently resourced to test for these pathogens, or to consider these pathogens within broader serosurveillance efforts, or indeed within more novel approaches such as metagenetic sequencing or wastewater surveillance.

Taking this further, we can build custom aggregations that complement activities designed to strengthen health systems for prevention, preparedness, readiness and response. Across the microbial spectrum pathogens present with similar syndromes, such as febrile illnesses, or with similar signs such as coughing - custom aggregations of these provide context for settings where rapid differential diagnoses of undiagnosed patients is a necessary precursor to clear and appropriate response actions, particularly when paired with contextual information on the general prevalence of those syndromes or most common causes of that syndrome ^31,32^. Similarly, we can consider the shared stressors that certain groupings of pathogens put on a health system, whether the provisioning of Personal Protective Equipment for healthcare workers for pathogens that are highly transmissible, or where supporting general infection prevention and control measures is going to be most relevant to a wide array of relevant pathogens, not just a singular microbial threat that may have served as the most recent reminder of the importance of these principles (Figure 3d). Clustering pathogens by shared comorbidity profiles similarly can be a motivating feature, particularly given the need to act over prolonged periods of time to affect a change on, e.g., obesity, versus the more immediate and consequential impact that providing a vaccine, or delivering a drug can achieve. During the COVID-19 pandemic assessments of population vulnerability using population-level prevalence of key co-morbidities served as important contextual resources for planning and were supported by resources that existed prior to the pandemic ^33^. We can similarly use these results to support the development of multi-pathogen profiles of at-risk populations, and cross-reference their ability to have timely access to high-quality care, or specialist facilities necessary to treat cases of specific pathogens, or test relevant samples ^34,35^. While Figure 3 focused on a limited subset of possible options, the necessary inputs to generate any other aggregation are presented here.

Finally, we can cross-reference this work when considering where next to enhance research into these pathogens supporting ongoing WHO efforts for epidemic and pandemic research preparedness ^36^. When thinking about different types of evidence, the environmental suitability assessments represented the most powerful means of differentiating areas of realized risk from a random background sample. As such, assessments that attempt to further explore the geographic variation present in prior cases is critical. Lassa virus represented a particularly compelling example, since it was the one pathogen for which we were able to identify a map of human seroprevalence across a comprehensive geographic range ^37^. The precision weighting of this resource (0.9215) was comparable to those of the 0.6 and 0.8 threshold environmental suitability layers. This is particularly compelling given the potential for the environmental suitability model to be overfit to the geographically resolved occurrence data used, while the seroprevalence layer was parameterized using a distinct resource. Expanding the geographic coverage of comparable efforts for other emerging pathogens that are priorities in the regional level (e.g. Oropouche virus (Americas), Ross River virus (Western Pacific) among others) if having comparable discriminatory capabilities, could prove transformative in a next generation of our maps ^38^.

In contrast, diseases such as plague, or avian Influenza indicate the potential for considerable benefit in terms of pinpointing areas of emphasis should global efforts to further map transmissibility be updated or made available. We hope that others can use this work to promote the revision of their models, or more widespread sharing of necessary data to feed into such kinds of assessments, as well as providing support for researchers and organizations to go and collate such data, or produce such estimates when existing resources are out-of-date, demonstrably lack precision, or are currently absent. The ongoing utility of such maps is contingent on the ability to integrate new information, and engineering systems that can leverage resources that provide close to real-time reporting of suspected and confirmed cases, such as ProMED ^39^, to automatically update and re-estimate emergence assessments ^40^. Only by doing so can we provide up-to-date guidance to support in-country decision making processes.

Future iterations of these maps would benefit from as up-to-date as possible assessments. While pre-existing resources are not insufficient for characterizing contemporary emergence, as evidenced by the Marburg example (Figure 4), other events, such as recent mortality events in sea lions and other species due to avian influenza ^41^, indicate that global assessments need to perpetually incorporate the most recent insights and dynamic environmental changes. While more contemporary assessments may exist than those integrated in this paper, not all were publicly available, and centralizing and standardizing otherwise disparate models and maps is an important challenge for the future. Our assessment of the discriminatory power of different evidence bases to differentiate reported detections from a background sample can be influenced by the global variation in detection capacities. Future iterations could explore how this varies and either sample using different kinds of bias grids, or re-sample detections to account for clustering in some highly surveilled populations versus comparative sparsity in others, or gaps that are merely artefacts of the lack of the ability to report. This evaluation used equal weighting for the final pathogen-specific aggregation. Future iterations should investigate the differential endpoints resulting from the use of other weights derived from burden statistics such as Disability Adjusted Life Years ^42^, economic impacts, or underreporting-adjusted report tallies. Along these lines, regional-specific weightings could be leveraged instead of a global standard, which might also accommodate variations in surveillance capabilities in different regions. For instance, the requirement for differentiation in higher-surveilled populations you would want higher than in settings where a lower percentage of all true cases are accurately identified.

## Conclusion

The broad health and societal impacts of pathogens with epidemic and pandemic potential necessitate the collation of an array of resources that touch upon a range of factors related to our understanding of their emergence potential. We recognize that for many diseases such insights already exist in some manner - by collating these different pieces and aligning summaries with ongoing preparedness activities and strengthening of surveillance systems, we hope to encourage continuous refinement of our core health intelligence resources, as well as provide a support tool for decisions related to where and how to prepare.

The emergence potential maps described here have been incorporated into WHO dashboards, e.g. for Dynamic Preparedness Metrics (DPM; https://extranet.who.int/sph/dpm) ^43,44^ and available online (composite and individual pathogens: https://experience.arcgis.com/experience/1236ef068f234bea90fe404f8308d5b6/page/Aedes-(Stegomyia)-borne-diseases/; by mode of transmission: https://experience.arcgis.com/experience/128eaafff2a14817962999263a9c111f), and are used in WHO’s Global Programme of Work 14 (GPW14) outcome indicators framework. Further to this, use of the emergence potential maps is currently explored for the development of a risk-needs metric for Pandemic Fund resources allocation.

## Data Availability

All data produced are available online at

https://github.com/beoutbreakprepared

## Acknowledgements

This work was concluded in 2024 and supported by the United States Agency for International Development (USAID), the Germany Agency for International Cooperation (GIZ) and the Government of France.

We express our gratitude to countries and partners for their surveillance efforts and to the ongoing efforts of various institutions for making their various data inputs available.

