## Supplemental materials for "Mapping global emergence of pathogens with epidemic and pandemic potential to inform and accelerate pandemic prevention, preparedness, readiness and response"

### Pathogen-specific Evidence

#### Avian Influenza virus

Overall assessment

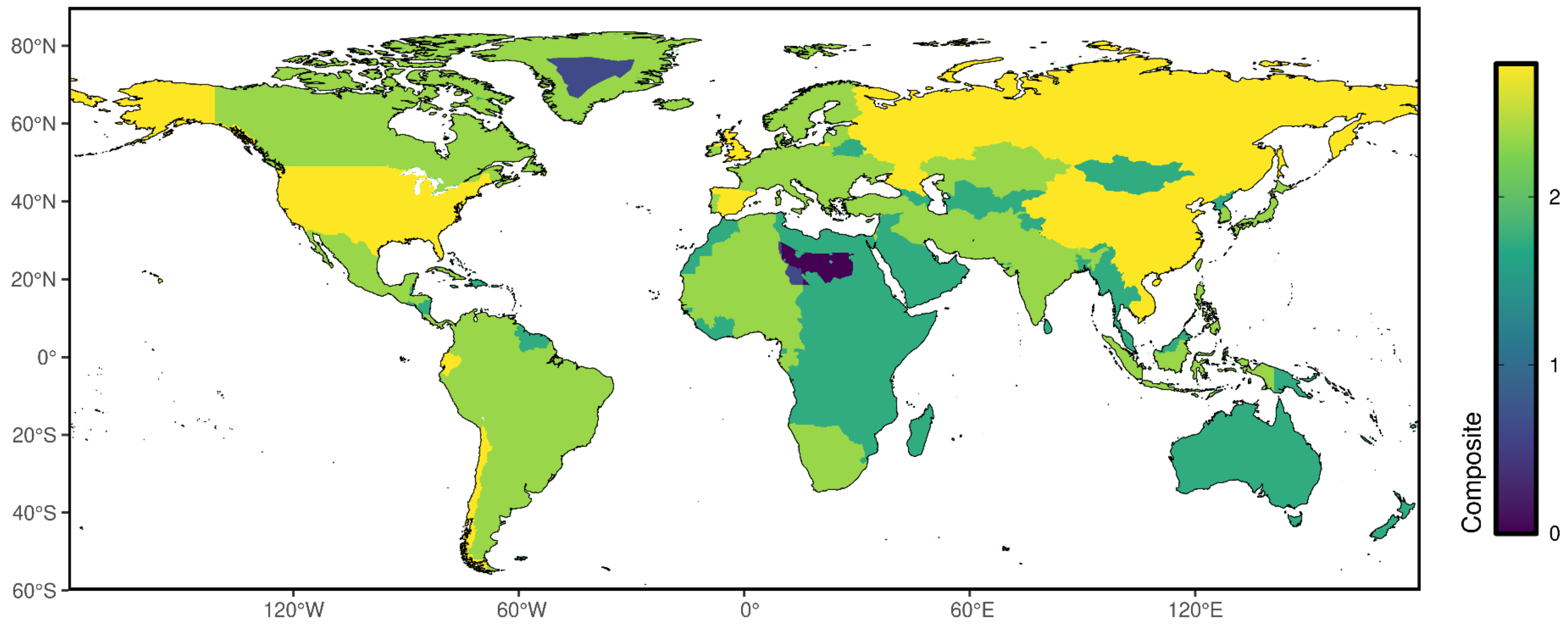

### Reservoir presence

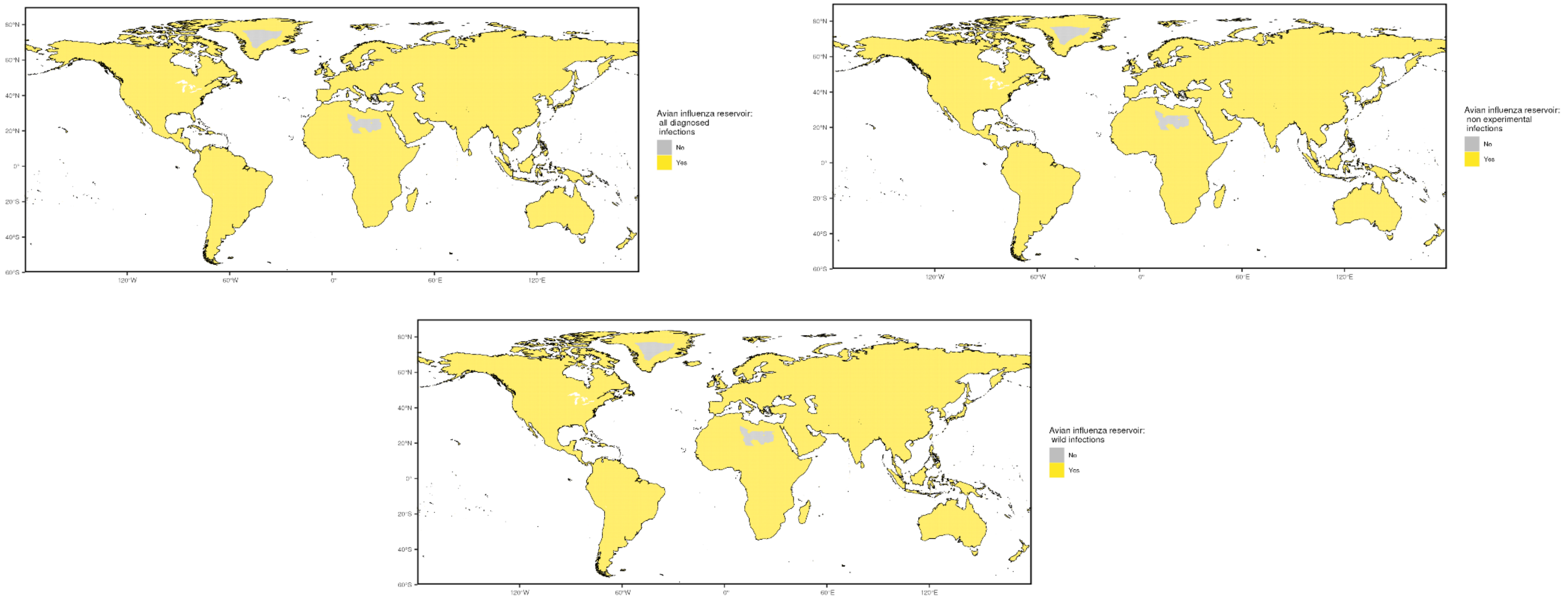

### Prior cases

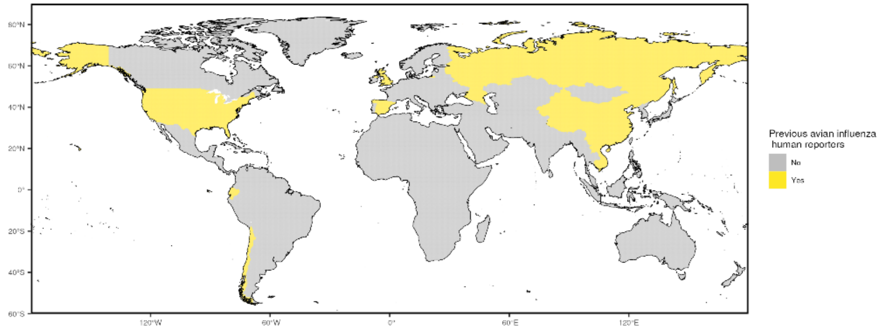


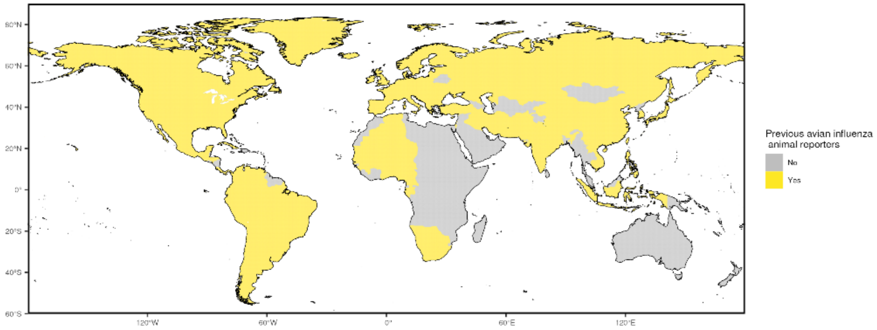


### Occurrence data

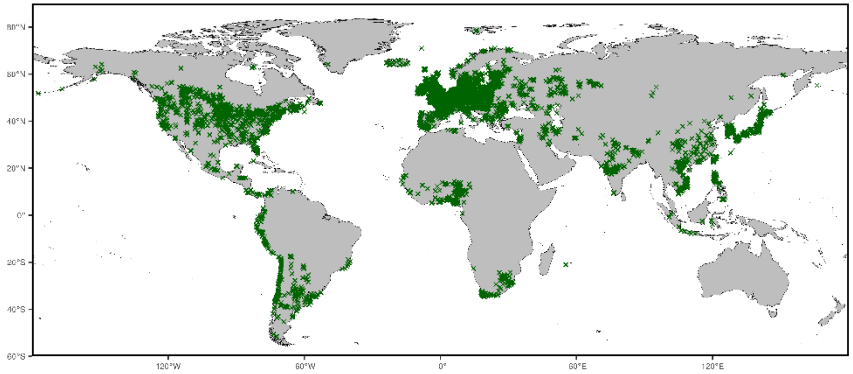

NOTE: centroids of polygons were used to map administrative and buffered occurrences, where relevant

### Precision weightings

| Precision categories | Precision weightings |
|---|---|
| All diagnosed infections | 0.574 |
| Non-experimental infections | 0.574 |
| Wild infections | 0.574 |
| Prior cases- animals | 0.623 |
| Prior cases- humans | 0.442 |

#### Data sources

Evidence data for Avian influenza disease were acquired from three main sources. First, we obtained a list of countries with transmission reports of avian influenza in animals and humans respectively compiled by the EMPRES-i database from 2021 until 31st May 2023. The EMPRES-i database, as well as the USDA-APHIS 2022-2023 databases for Highly Pathogenic Avian Influenza in Mammals and Wild Birds, were also used to compile a list of mammalian and avian non-human species with evidence of avian influenza used to create a zoonotic transmission layer. These species were stratified into three groupings: all reported species, all non-experimental reports, and all wild infections. The geographic range for each mammalian species was obtained from the International Union for Conservation of Nature's (IUCN) Red List of Threatened Species, and the ranges for each avian species was obtained from the BirdLife International and Handbook of the Birds of the World.

Occurrence data for obtaining precision estimates from each layer utilized both animal and human case data, which were obtained from the EMPRES-i database.

| **Evidence category** | **Citation** |
|---|---|
| **Countries with reported human cases** | FAO . 2023. EMPRES-i - Global Animal Disease Information System. Available at: https://empres-i.apps.fao.org/. (Accessed 31 May 2023) |
| **Countries with reported animal cases** | FAO . 2023. EMPRES-i - Global Animal Disease Information System. Available at: https://empres-i.apps.fao.org/. (Accessed 31 May 2023) |
| **Reservoir status determination** | 1. FAO . 2023. EMPRES-i - Global Animal Disease Information System. Available at: https://empres-i.apps.fao.org/. (Accessed 31 May 2023)<br>2. USDA-APHIS . 2023. 2022-2023 Detections of Highly Pathogenic Avian Influenza in Mammals. Available at: https://www.aphis.usda.gov/aphis/ourfocus/animalhealth/animal-disease-information/avian/avian-influenza/hpai-2022/2022-hpai-mammals/. (Accessed 10 May 2023)<br>3. USDA-APHIS . 2023. 2022-2023 Detections of Highly Pathogenic Avian Influenza in Wild Birds. Available at: https://www.aphis.usda.gov/aphis/ourfocus/animalhealth/animal-disease-information/avian/avian-influenza/hpai-2022/2022-hpai-wild-birds/. (Accessed 10 May 2023) |
| **Human and animal geopositioned occurrences** | FAO . 2023. EMPRES-i - Global Animal Disease Information System. Available at: https://empres-i.apps.fao.org/. (Accessed 31 May 2023) |

### Pathogen-specific Evidence

#### Crimean-Congo haemorrhagic fever

Overall assessment

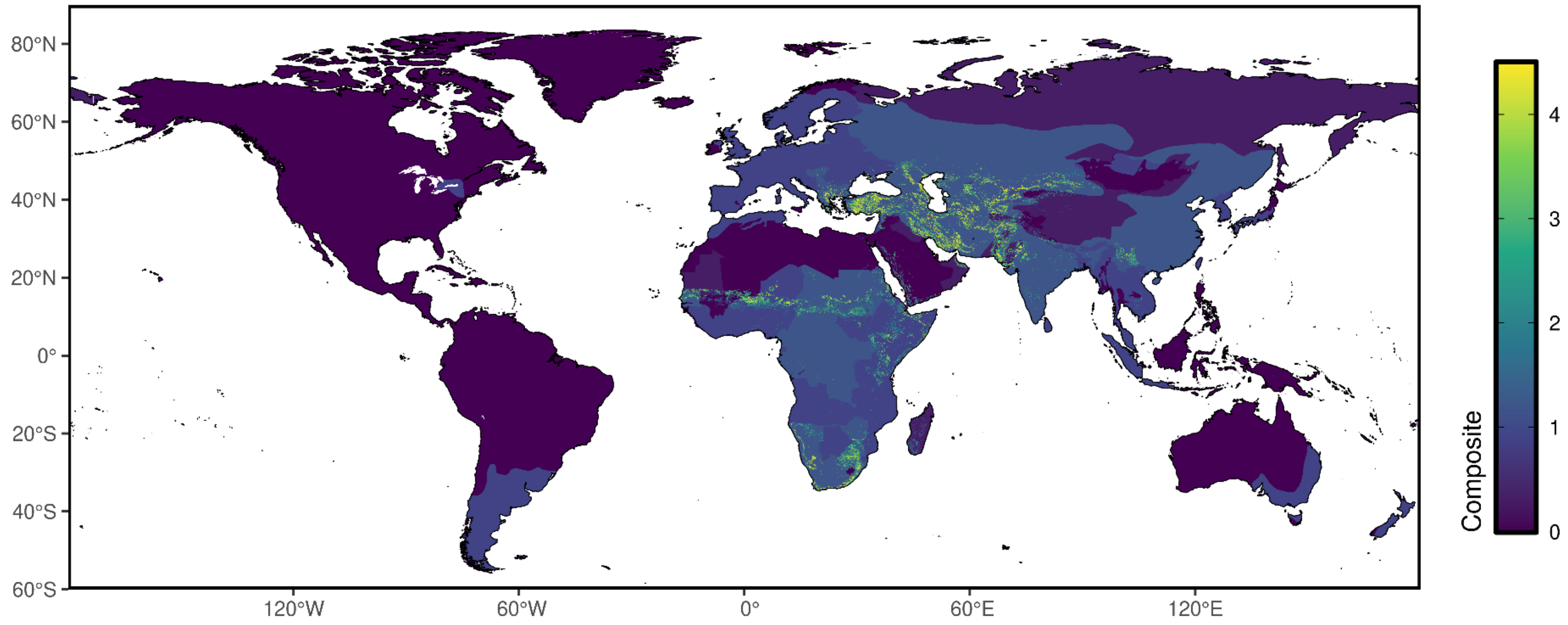

### Reservoir presence

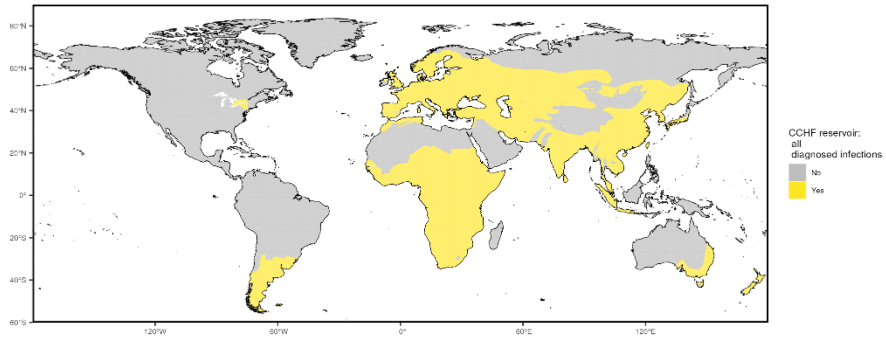


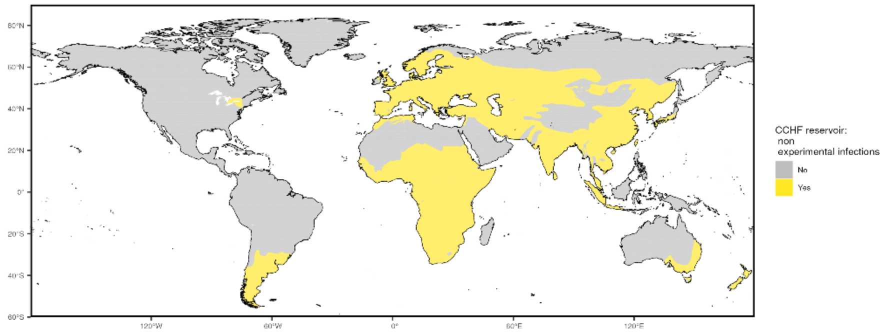


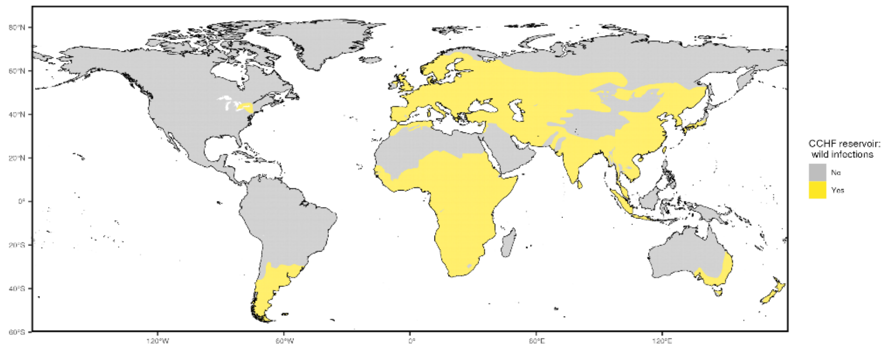

### Niche classifications

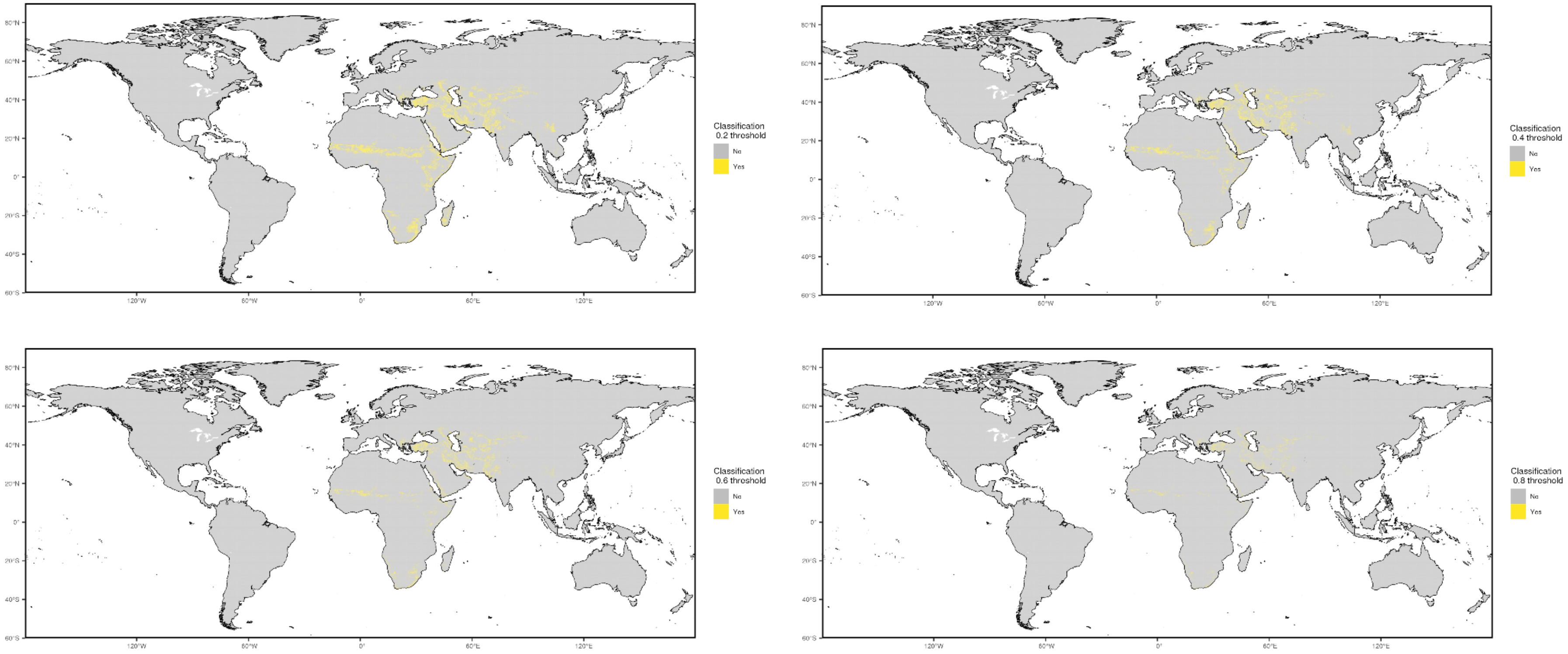

#### Prior cases

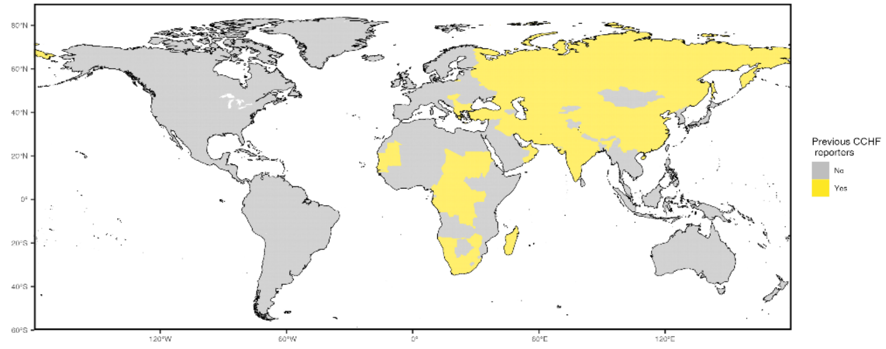


#### Occurrence data

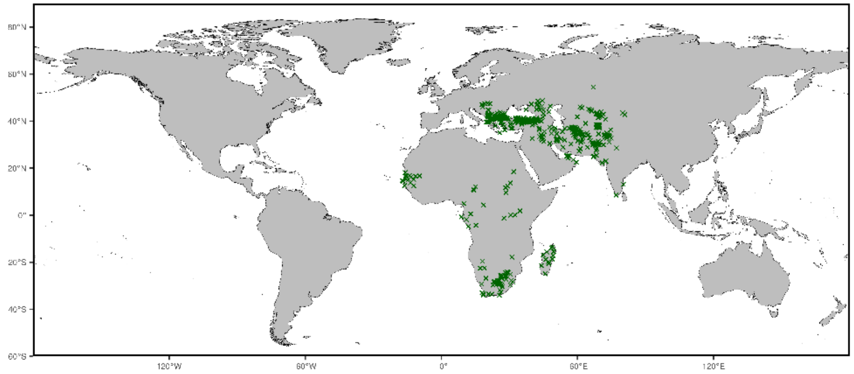


NOTE: centroids of polygons were used to map administrative and buffered occurrences, where relevant

Precision weightings

| Precision categories | Precision weightings |
|---|---|
| Prior cases | 0.3032 |
| Niche classifications (0.2) | 0.7721 |
| Niche classifications (0.4) | 0.8115 |
| Niche classifications (0.6) | 0.8479 |
| Niche classifications (0.8) | 0.8743 |
| All diagnosed infections | 0.2955 |
| Non-experimental infections | 0.2955 |
| Wild infections | 0.2955 |
| Prior cases | 0.3032 |

#### Data sources

Evidence data for Crimean-Congo Haemorrhagic Fever (CCHF) were acquired from two main sources. First, we obtained a list of countries with historic transmission reports of CCHF compiled by Messina et al. (2015). Second, an environmental suitability synoptic niche map with continuous suitability estimates from Messina et al. was used to create evidence suitability layers for CCHF with cutoff scores of 0.2, 0.4, 0.6, and 0.8. This suitability layer was constructed using human and mammalian occurrence data and relevant environmental covariates (namely, mean annual enhanced vegetation index and the SD of this mean, and shrub or grass landcover) to predict the estimated environmental suitability for CCHF transmission. Third, mammalian non-human species with evidence of Crimean-Congo Haemorrhagic Fever infections as reported to GIDEON were used were used in creating a zoonotic transmission layer, and these species were stratified into four groupings: all reported species, all non-experimental reports, all wild infections, and all wild, PCR positive infections. The geographic range for each species was obtained from the International Union for Conservation of Nature's (IUCN) Red List of Threatened Species.

Occurrence data for obtaining precision estimates from each layer were obtained from Messina et al. (2015) and were collated using a comprehensive, systematic review and data extraction process.

| Evidence category | Citation |
|---|---|
| **Countries with reported human cases** | Messina et al. (2015) "The global distribution of Crimean-Congo hemorrhagic fever" Trans Roy Soc Trop Med Hyg https://pubmed.ncbi.nlm.nih.gov/26142451/ |
| **Environmental suitability** | Messina et al. (2015) "The global distribution of Crimean-Congo hemorrhagic fever" Trans Roy Soc Trop Med Hyg https://pubmed.ncbi.nlm.nih.gov/26142451/ |
| **Reservoir status determination** | Berger. (2005) "GIDEON: a comprehensive Web-based resource for geographic medicine" Int. J. Health Geogr. 4, 10 |
| **Human geopositioned occurrences** | Messina et al. (2015) "The global distribution of Crimean-Congo hemorrhagic fever" Trans Roy Soc Trop Med Hyg https://pubmed.ncbi.nlm.nih.gov/26142451/ |

### Pathogen-specific Evidence

#### Chikungunya virus

Overall assessment

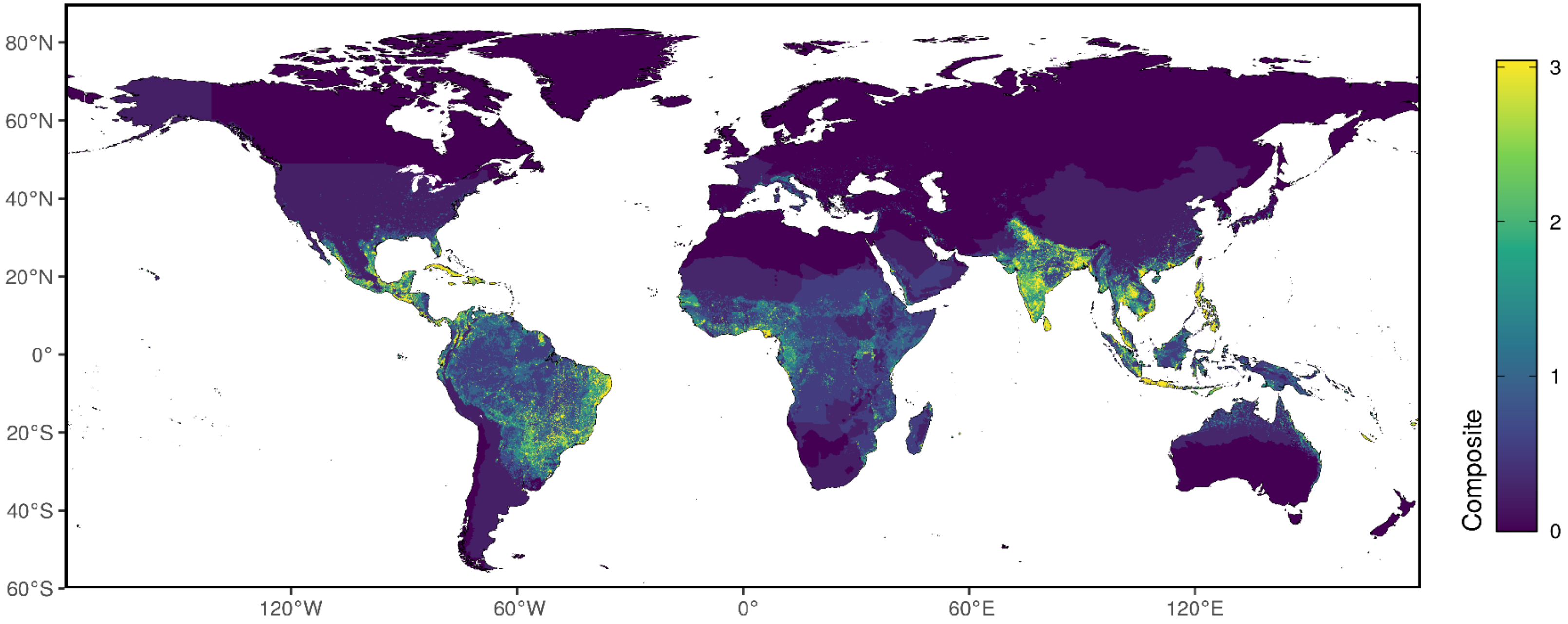

Aedes presence

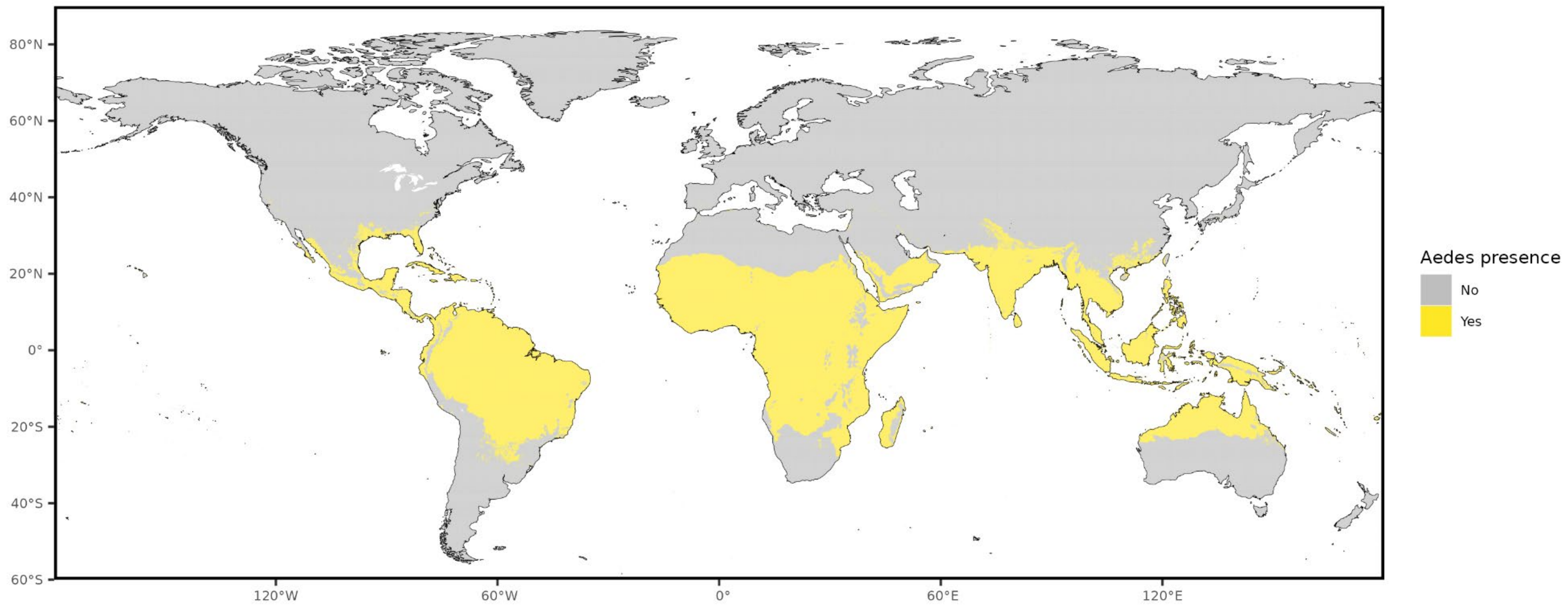

### Niche classifications

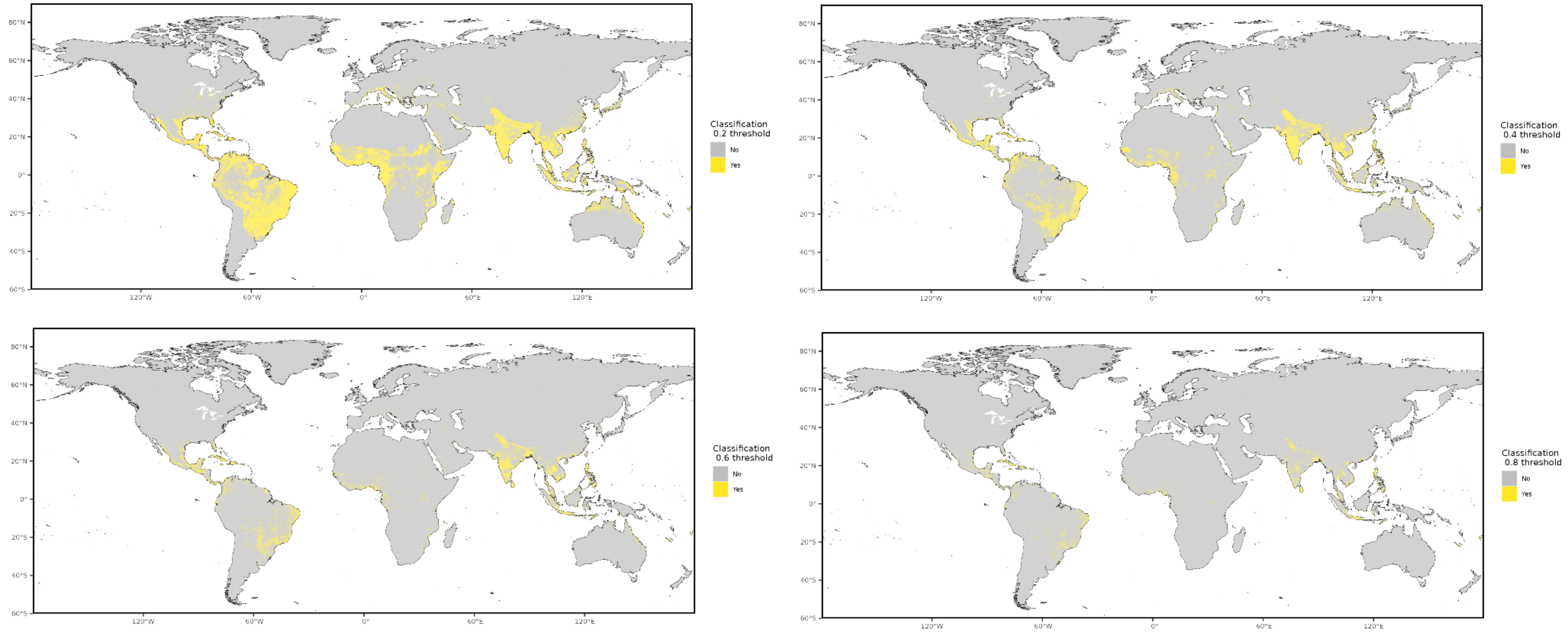

#### Prior cases

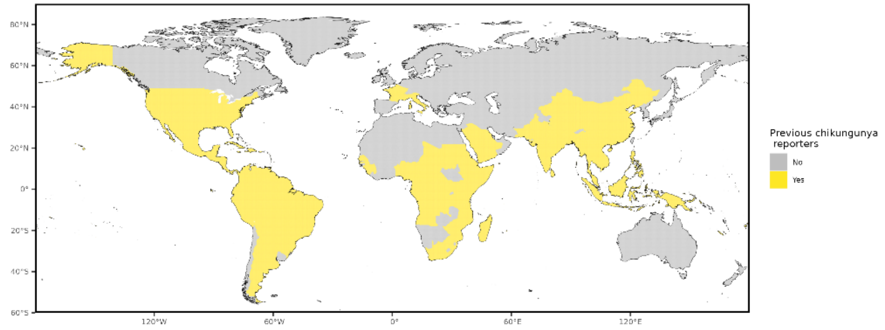


#### Occurrence data

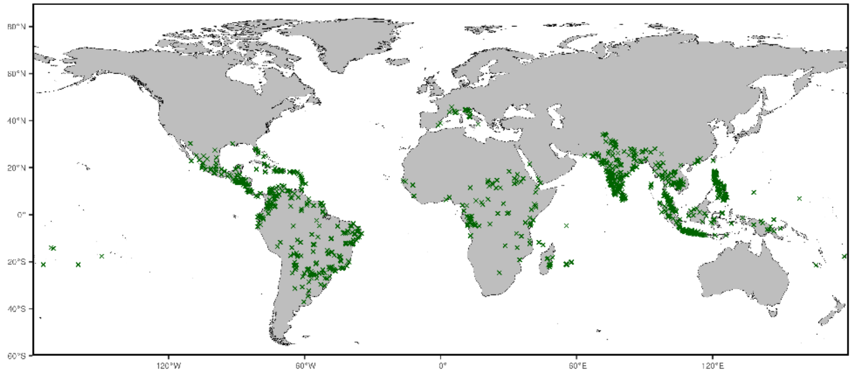


NOTE: centroids of polygons were used to map administrative and buffered occurrences, where relevant

#### Precision weightings

| **Precision categories** | **Precision weightings** |
| --- | --- |
| Aedes presence | 0.2891 |
| Prior cases | 0.2239 |
| Niche classifications (0.2) | 0.4240 |
| Niche classifications (0.4) | 0.5696 |
| Niche classifications (0.6) | 0.7048 |
| Niche classifications (0.8) | 0.8270 |

#### Data sources

Evidence data for Chikungunya were acquired from three main sources. First, we obtained a list of countries with historic transmission reports of aedes-borne pathogens, subset specifically to Chikungunya, compiled by the WHO. Second, an environmental suitability synoptic niche map with continuous suitability estimates from Lim et al. (2025) was used to create evidence suitability layers for Chikungunya virus with cutoff scores of 0.2, 0.4, 0.6, and 0.8. Third, the global distribution of arbovirus vectors *Aedes aegypti* and *Aedes albopictus* as published by Kraemer et al. in 2015 were included as a vector layer.

Occurrence data for obtaining precision estimates from each layer were obtained from Lim et al. (2025).

| **Evidence category** | **Citation** |
|---|---|
| **Countries with reported human cases** | World Health Organization internal database |
| **Environmental suitability** | Lim, A., Shearer, F.M., Sewalk, K. *et al.* The overlapping global distribution of dengue, chikungunya, Zika and yellow fever. *Nat Commun* **16**, 3418 (2025). https://doi.org/10.1038/s41467-025-58609-5 |
| **Vector distribution** | Kraemer et al. (2015) "The global distribution of the arbovirus vectors Aedes aegypti and Ae. albopictus" eLife https://elifesciences.org/articles/08347 |
| **Human geopositioned occurrences** | Lim, A., Shearer, F.M., Sewalk, K. *et al.* The overlapping global distribution of dengue, chikungunya, Zika and yellow fever. *Nat Commun* **16**, 3418 (2025). https://doi.org/10.1038/s41467-025-58609-5 |

### Pathogen-specific Evidence

#### Dengue fever

Overall assessment

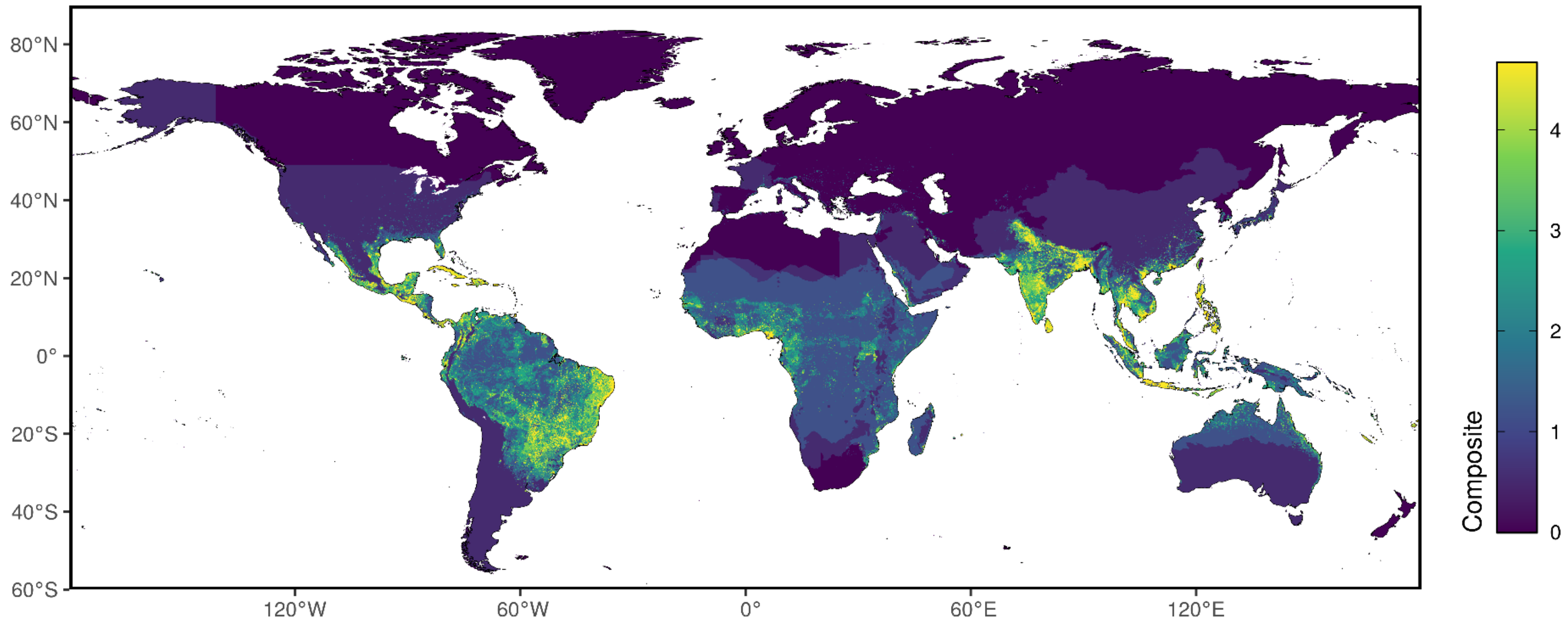

Aedes presence

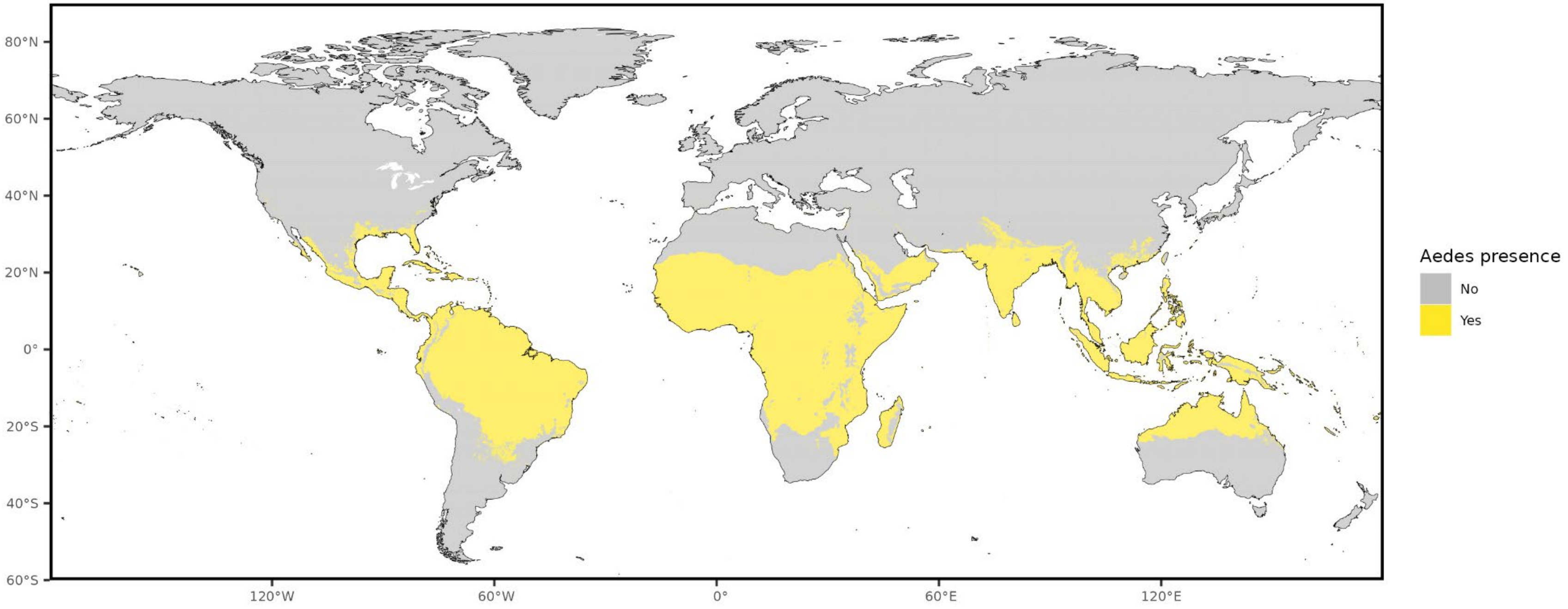

### Niche classifications

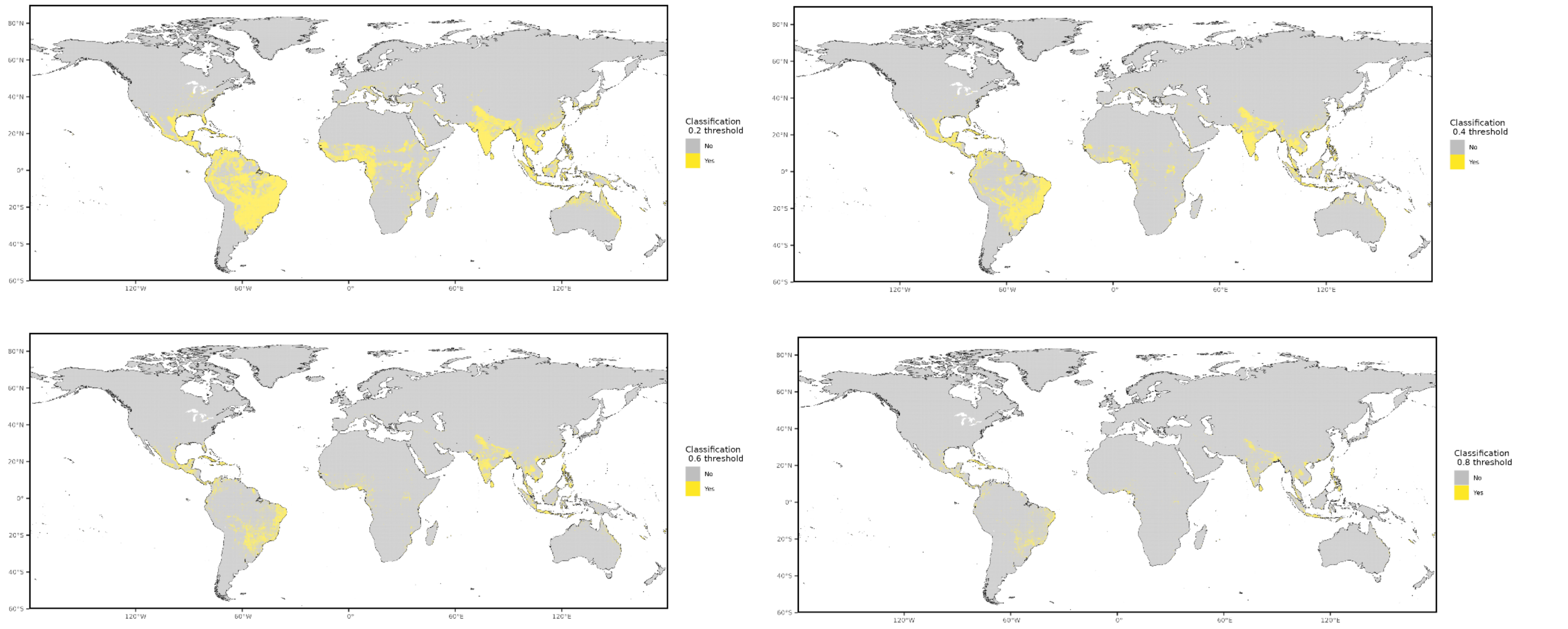

#### Prior cases

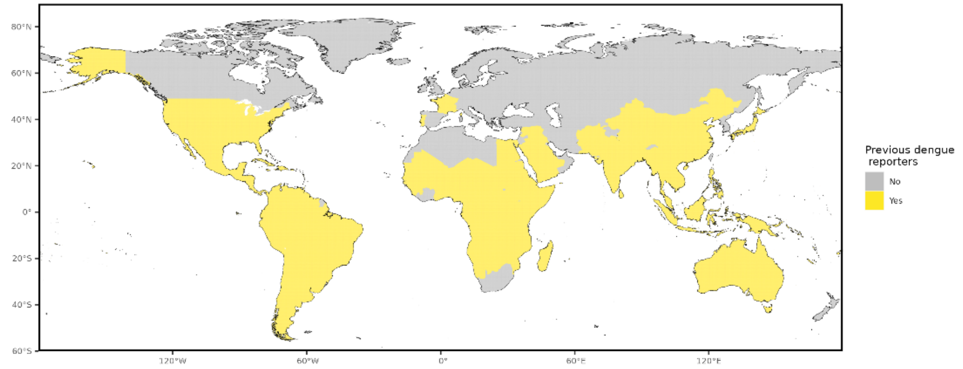


#### Occurrence data

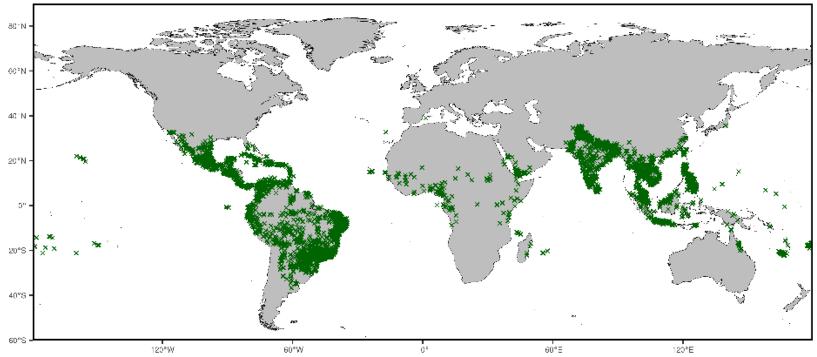

NOTE: centroids of polygons were used to map administrative and buffered occurrences, where relevant

#### Precision weightings

| **Precision categories** | **Precision weightings** |
|---|---|
| Aedes presence | 0.6448 |
| Prior cases | 0.5119 |
| Niche classifications (0.2) | 0.7813 |
| Niche classifications (0.4) | 0.8537 |
| Niche classifications (0.6) | 0.9148 |
| Niche classifications (0.8) | 0.9586 |

#### Data sources

Evidence data for Dengue were acquired from three main sources. First, we obtained a list of countries with historic transmission reports of aedes-borne pathogens, subset specifically to Dengue, compiled by the WHO. Second, an environmental suitability synoptic niche map with continuous suitability estimates from Lim et al. (2025) was used to create evidence suitability layers for Dengue virus with cutoff scores of 0.2, 0.4, 0.6, and 0.8. Third, the global distribution of arbovirus vectors *Aedes aegypti* and *Aedes albopictus* as published by Kraemer et al. in 2015 were included as a vector layer.

Occurrence data for obtaining precision estimates from each layer were obtained from Lim et al. (2025)).

| **Evidence category** | **Citation** |
|---|---|
| **Countries with reported human cases** | World Health Organization internal database |
| **Environmental suitability** | Lim, A., Shearer, F.M., Sewalk, K. *et al.* The overlapping global distribution of dengue, chikungunya, Zika and yellow fever. *Nat Commun* **16**, 3418 (2025). https://doi.org/10.1038/s41467-025-58609-5 |
| **Vector distribution** | Kraemer et al. (2015) "The global distribution of the arbovirus vectors Aedes aegypti and Ae. albopictus" eLife https://elifesciences.org/articles/08347 |
| **Human geopositioned occurrences** | Lim, A., Shearer, F.M., Sewalk, K. *et al.* The overlapping global distribution of dengue, chikungunya, Zika and yellow fever. *Nat Commun* **16**, 3418 (2025). https://doi.org/10.1038/s41467-025-58609-5 |

### Pathogen-specific Evidence

#### Ebola virus

Overall assessment

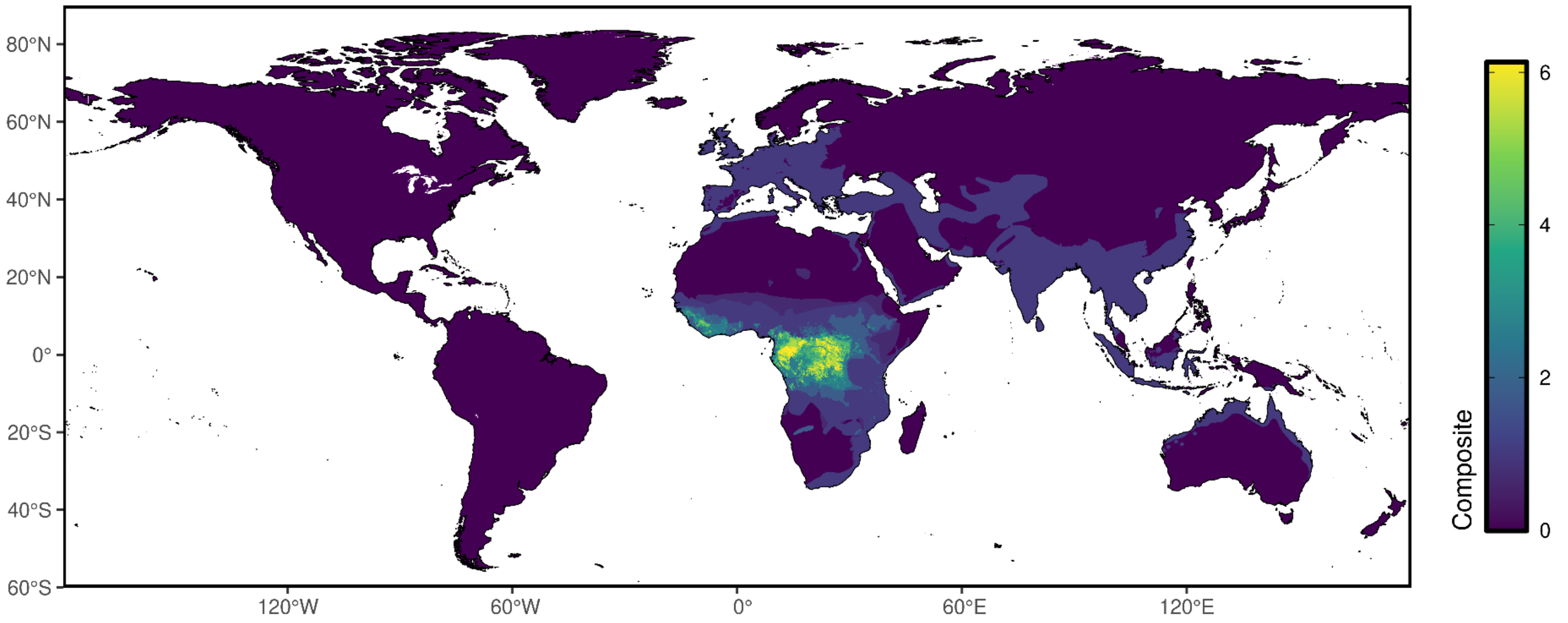

### Reservoir presence

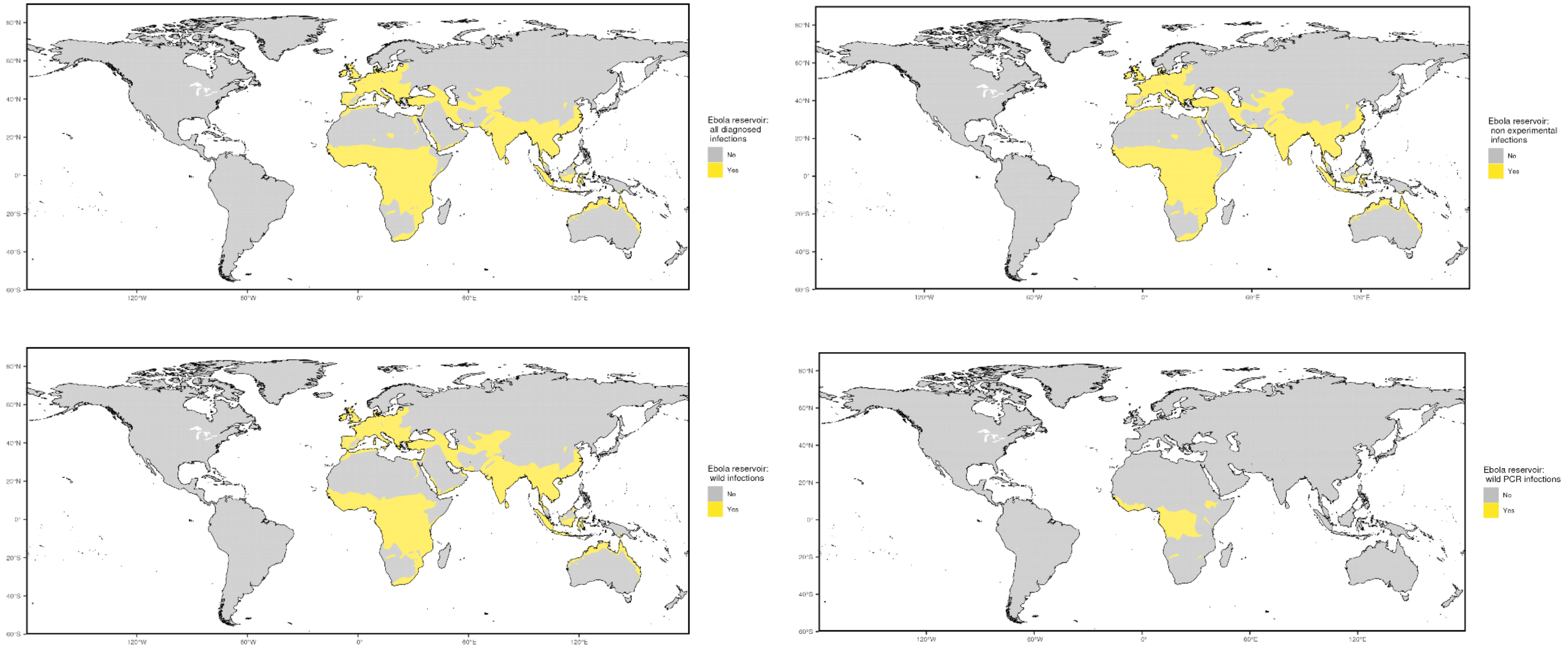

### Niche classifications

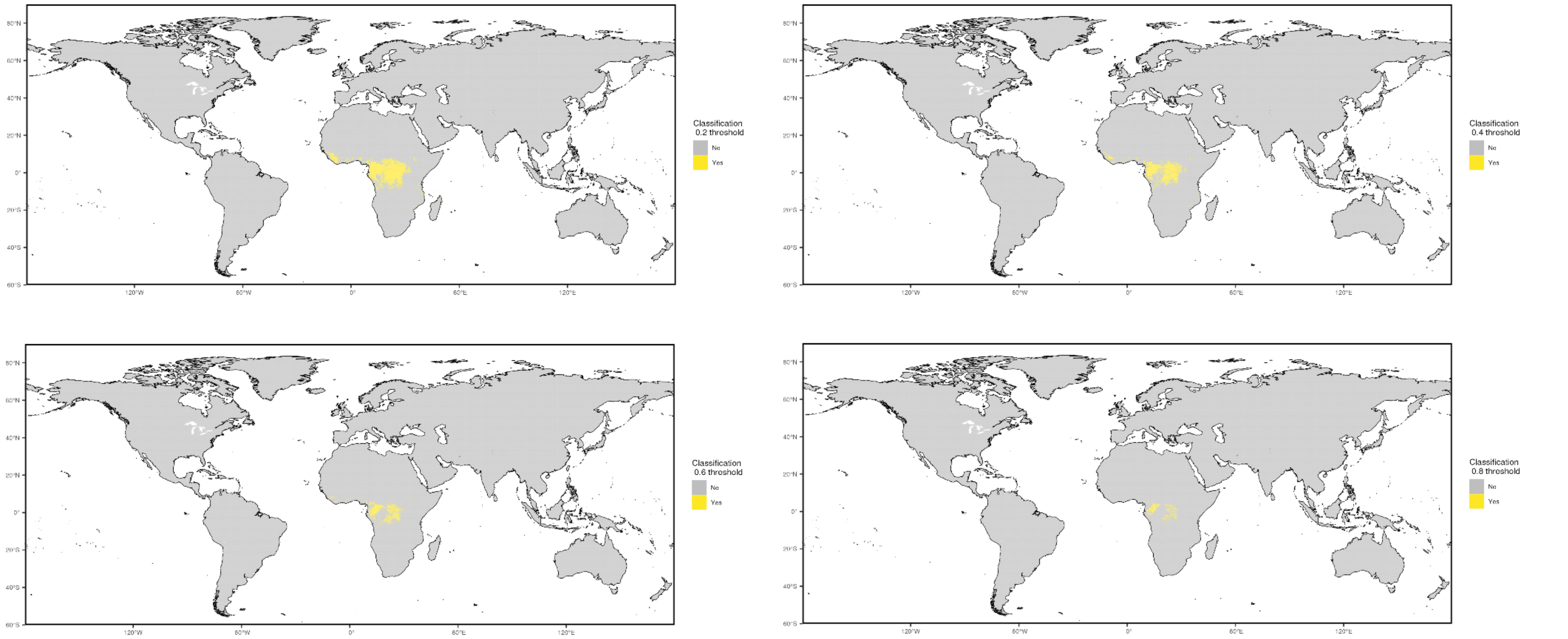

#### Prior cases

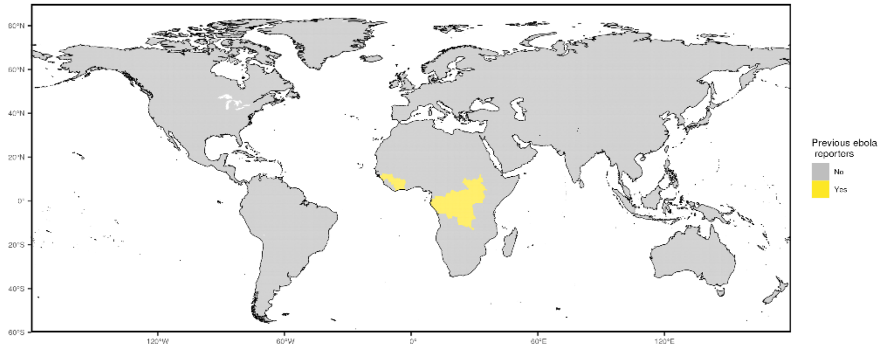


#### Occurrence data

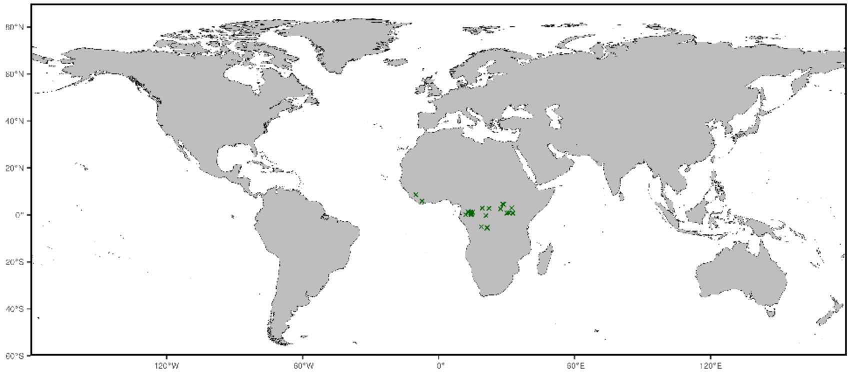


NOTE: centroids of polygons were used to map administrative and buffered occurrences, where relevant

#### Precision weightings

| **Precision categories** | **Precision weightings** |
|---|---|
| Prior cases | 0.7949 |
| Niche classifications (0.2) | 0.8108 |
| Niche classifications (0.4) | 0.871 |
| Niche classifications (0.6) | 0.9583 |
| Niche classifications (0.8) | 0.9474 |
| All diagnosed infections | 0.3263 |
| Non-experimental infections | 0.3263 |
| Wild infections | 0.337 |
| Wild/PCR+ infections | 0.75 |

#### Data sources

Evidence data for Ebola virus disease were acquired from three main sources. First, we obtained a list of countries with historic transmission reports of Ebola compiled by the CDC. Second, an environmental suitability synoptic niche map with continuous suitability estimates from Pigott et al. was used to create evidence suitability layers for Ebola with cutoff scores of 0.2, 0.4, 0.6, and 0.8. This suitability layer was constructed using human and mammalian occurrence data and relevant environmental covariates (namely, enhanced vegetation index, land surface temperature, elevation, potential evapotranspiration, and the distributions of implicated bat reservoir species) to predict the estimated environmental suitability for Ebola transmission. Third, mammalian non-human species with evidence of Ebola infections, identified by Han et al. (2016) were used in creating a zoonotic transmission layer, and these species were stratified into four groupings: all reported species, all non-experimental reports, all wild infections, and all wild, PCR positive infections. The geographic range for each species was obtained from the International Union for Conservation of Nature's (IUCN) Red List of Threatened Species.

Occurrence data for obtaining precision estimates from each layer were obtained from Pigott et al. (2016).

| **Evidence category** | **Citation** |
|---|---|
| **Countries with reported human cases** | US Centers for Disease Control - https://www.cdc.gov/vhf/ebola/history/chronology.html |
| **Environmental suitability** | Pigott et al. (2016) "Updates to the zoonotic niche map of Ebola virus disease in Africa" eLife 5 e16412 |
| **Reservoir status determination** | Han et al. (2016) "Undiscovered Bat Hosts of Filoviruses" PLoS NTDs 10(7) e0004815 |
| **Human geopositioned occurrences** | Pigott et al. (2016) "Updates to the zoonotic niche map of Ebola virus disease in Africa" eLife 5 e16412 |

### Pathogen-specific Evidence

#### Hendra virus

Overall assessment

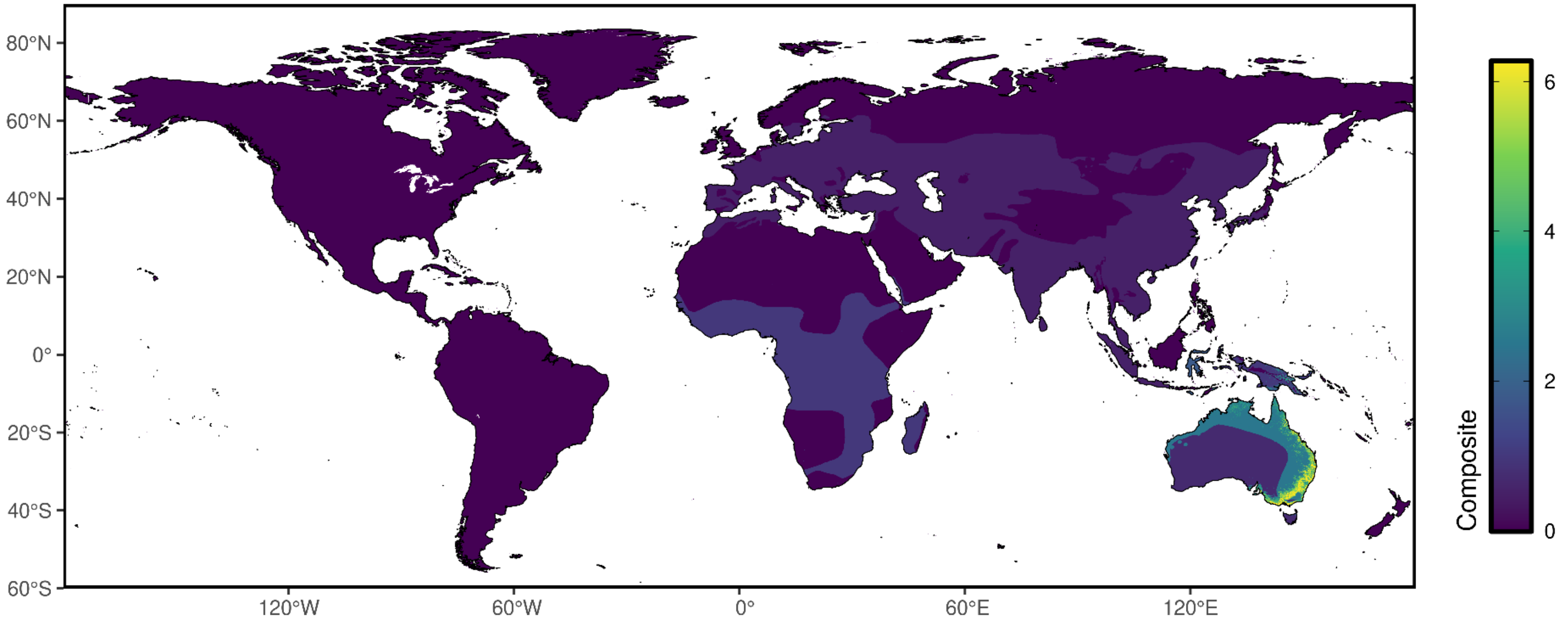

### Reservoir presence

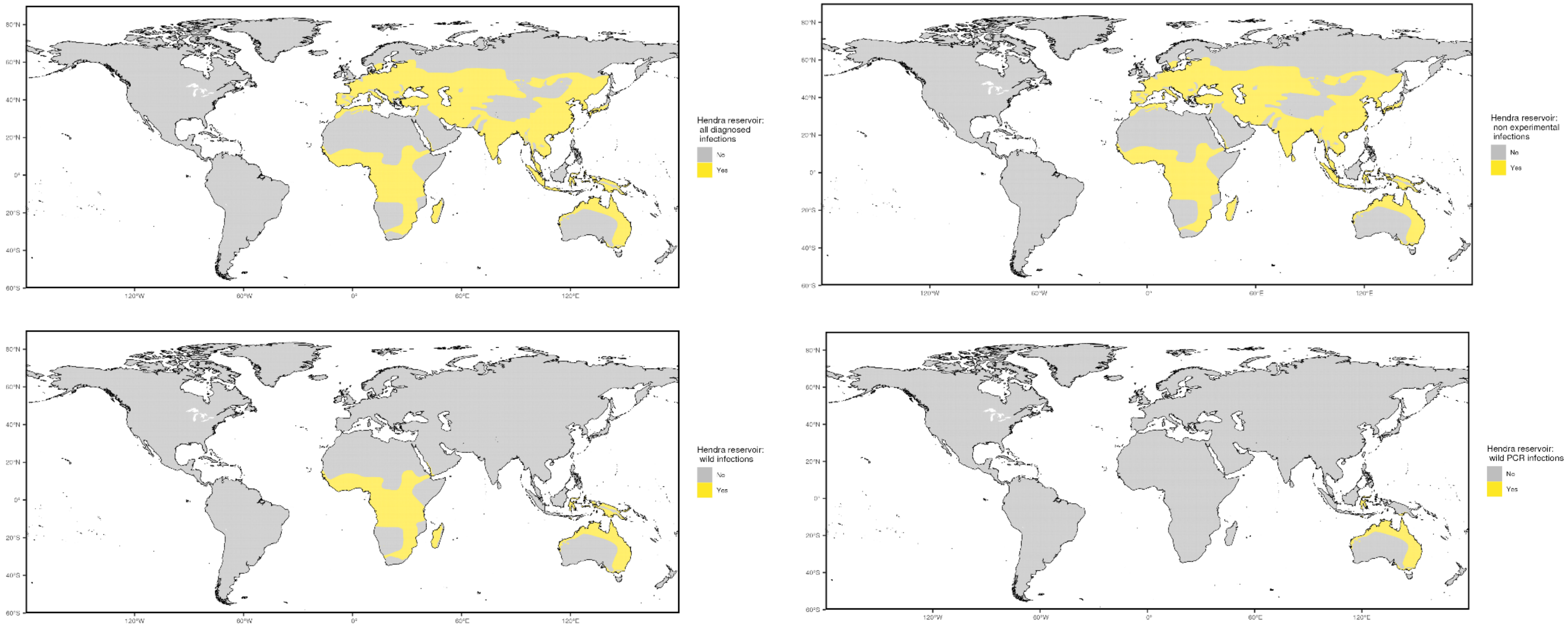

Niche classifications

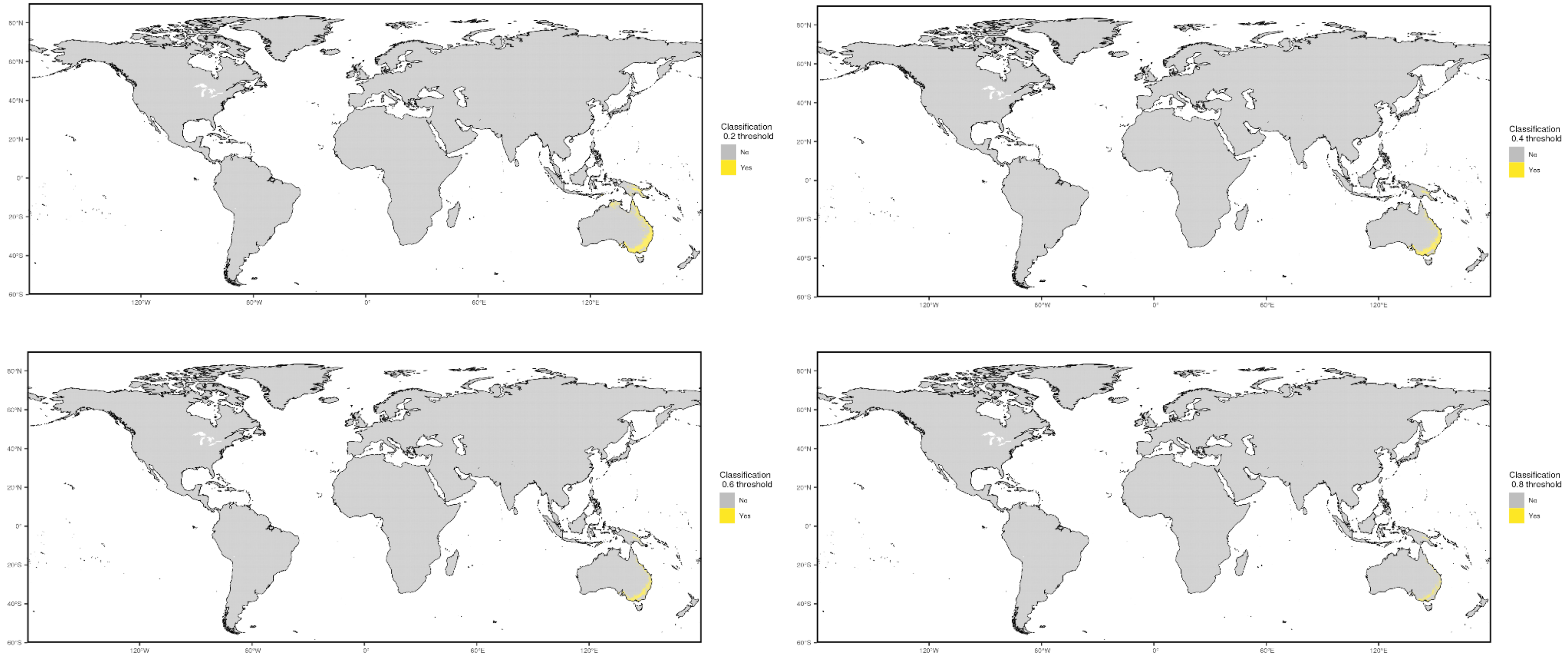


Prior cases

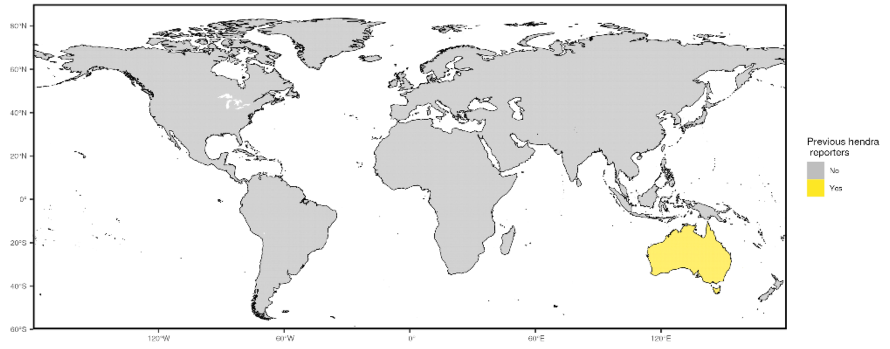


#### Occurrence data

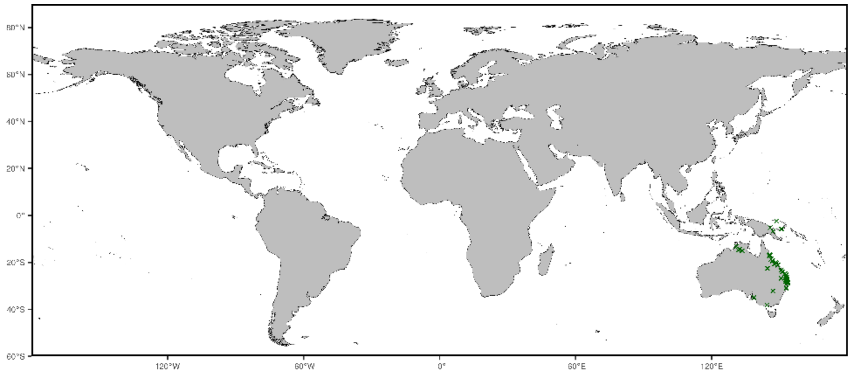


NOTE: centroids of polygons were used to map administrative and buffered occurrences, where relevant

#### Precision weightings

| **Precision categories** | **Precision weightings** |
|---|---|
| Prior cases | 0.6721 |
| Niche classifications (0.2) | 0.8944 |
| Niche classifications (0.4) | 0.9452 |
| Niche classifications (0.6) | 0.9754 |
| Niche classifications (0.8) | 0.98 |
| All diagnosed infections | 0.246 |
| Non-experimental infections | 0.246 |
| Wild infections | 0.4814 |
| Wild/PCR+ infections | 0.825 |

#### Data sources

Evidence data for Hendra were acquired from three main sources. First, we obtained a list of countries with historic transmission reports of Hendra compiled by the WHO. Second, an environmental suitability synoptic niche map with continuous suitability estimates from unpublished research by Pigott et al. was used to create evidence suitability layers for Hendra virus with cutoff scores of 0.2, 0.4, 0.6, and 0.8. This suitability layer was constructed using human and mammalian occurrence data through 2017 and relevant environmental covariates (namely, enhanced vegetation index, cumulative precipitation, mean temperature, tasseled cap brightness, tasseled cap wetness, and a consolidated indicator of species distribution models of implicated bat species) to predict the estimated environmental suitability for Hendra transmission. Third, non-human species with evidence of Hendra as reported to GIDEON were used in creating a zoonotic transmission layer, and these species were stratified into four groupings: all reported species, all non-experimental reports, all wild infections, and all wild, PCR positive infections. The geographic range for each species was obtained from the International Union for Conservation of Nature's (IUCN) Red List of Threatened Species.

Occurrence data for obtaining precision estimates from each layer were obtained from unpublished Pigott et al. and were collated using a comprehensive, systematic review and data extraction process.

| Evidence category | Citation |
|---|---|
| **Countries with reported human cases** | World Health Organization https://www.who.int/health-topics/hendra-virus-disease#tab=tab_1 |
| **Environmental suitability** | Pigott et al. *Unpublished.* "Mapping the environmental suitability for the Henipaviruses: Hendra and Nipah" |
| **Reservoir status determination** | Berger. (2005) "GIDEON: a comprehensive Web-based resource for geographic medicine" Int. J. Health Geogr. 4, 10 |
| **Human geopositioned occurrences** | Pigott et al. *Unpublished.* "Mapping the environmental suitability for the Henipaviruses: Hendra and Nipah" |

### Pathogen-specific Evidence

#### Henipaviruses

Overall assessment

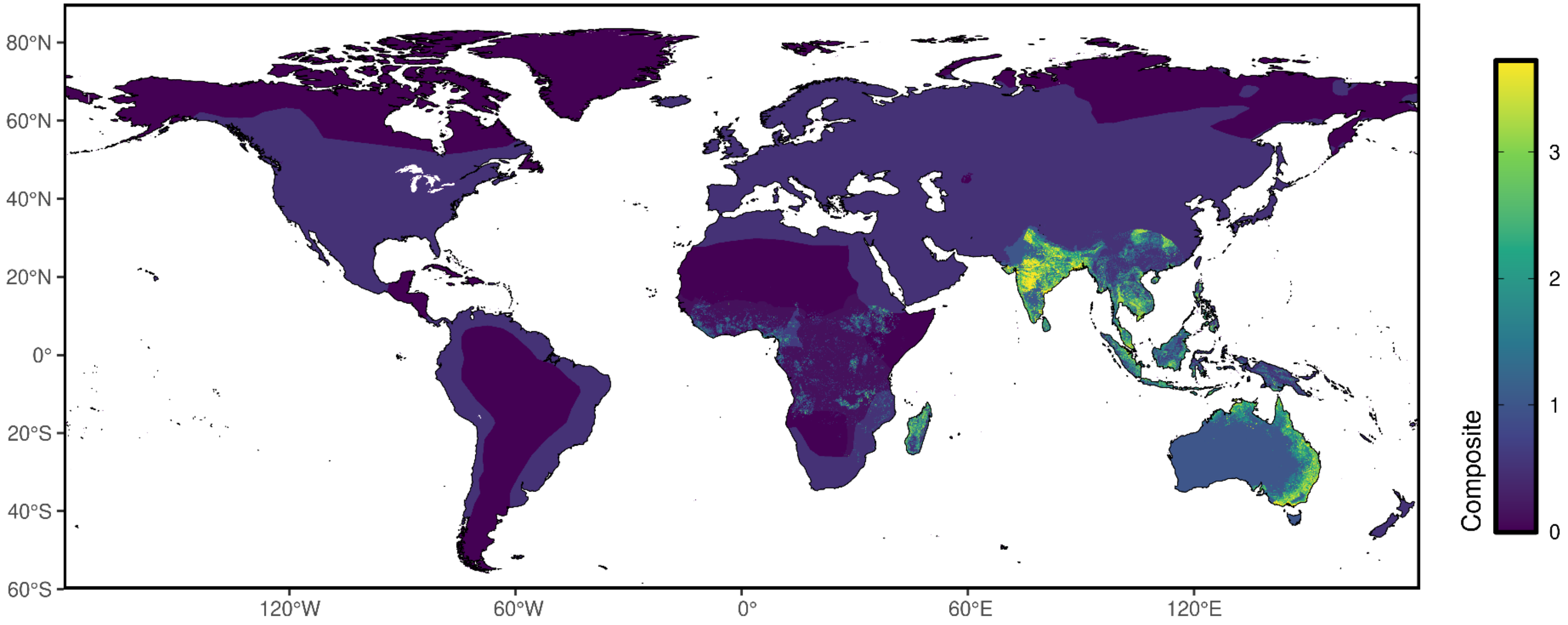

### Reservoir presence

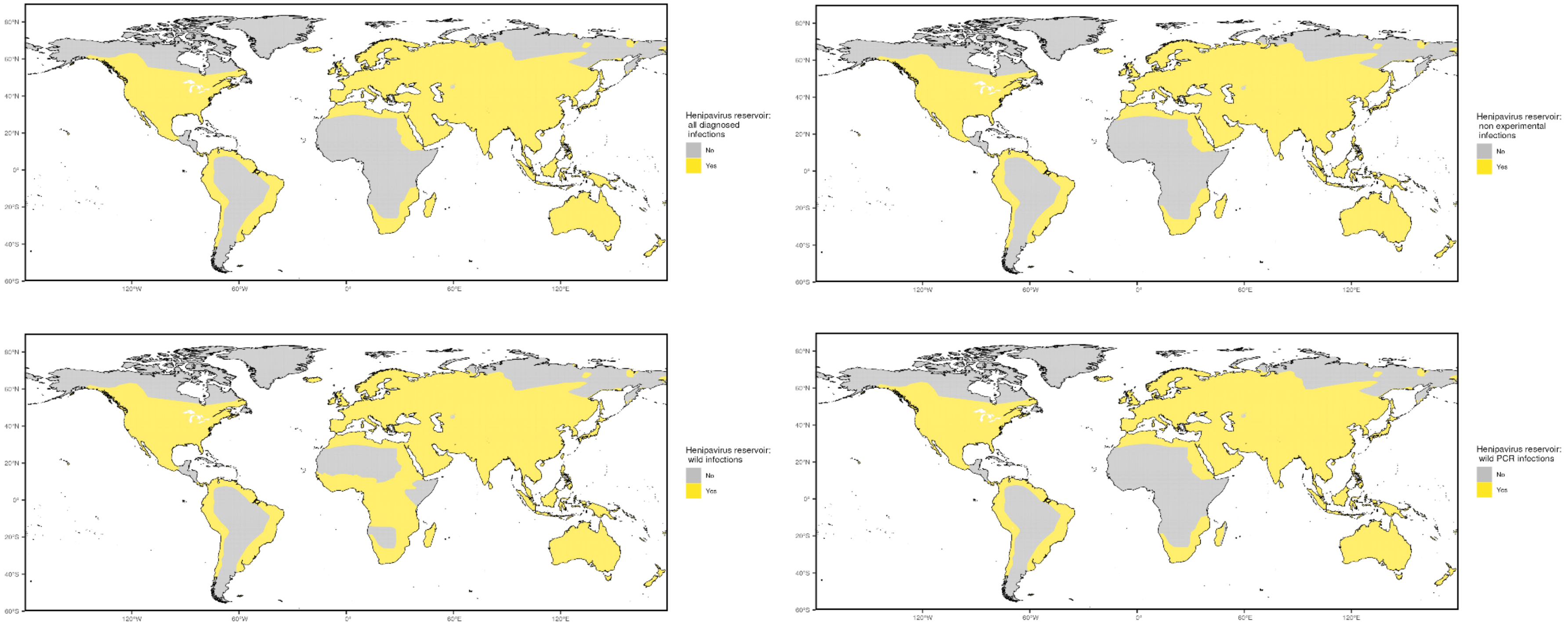

### Niche classifications

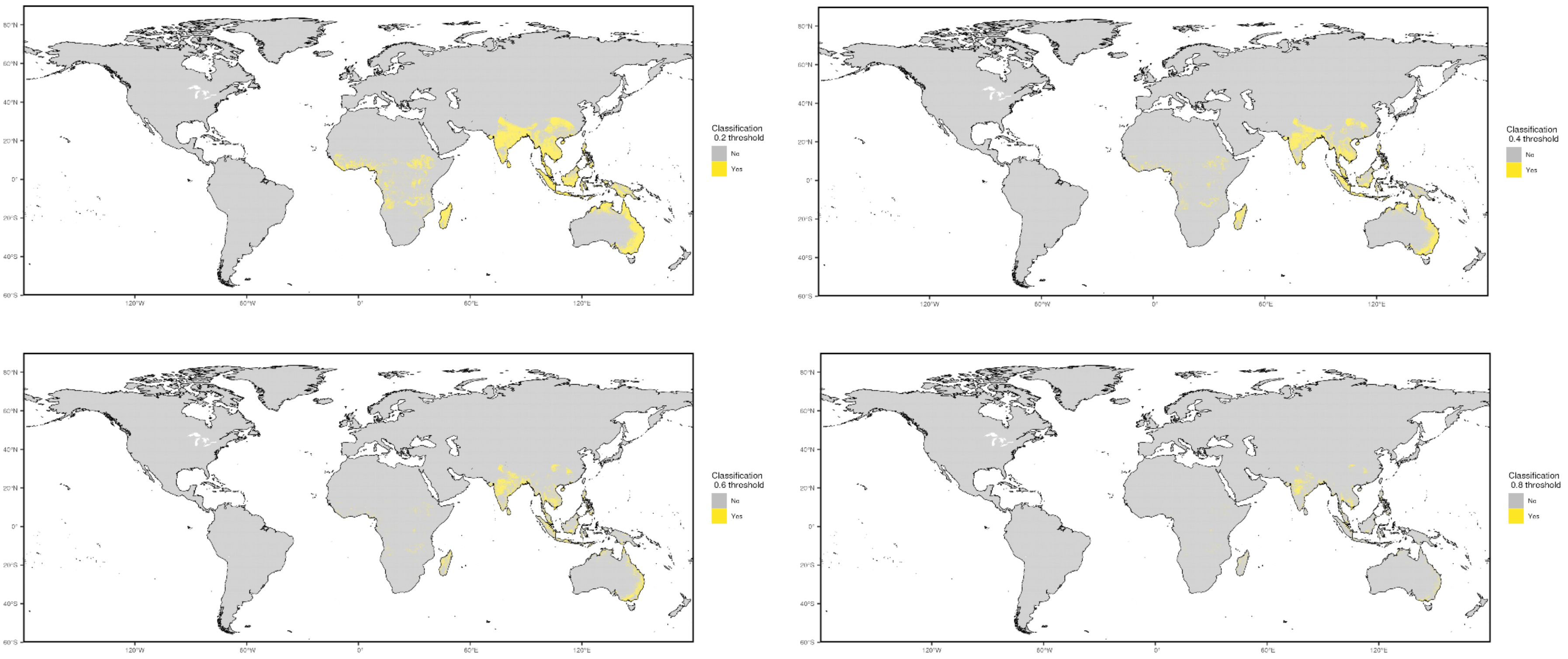

#### Prior cases

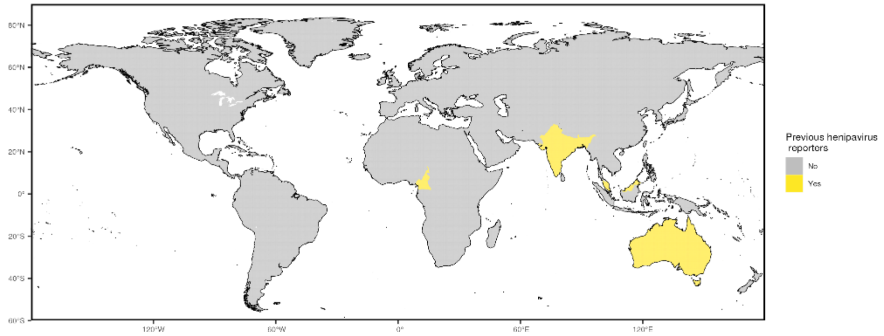


#### Occurrence data

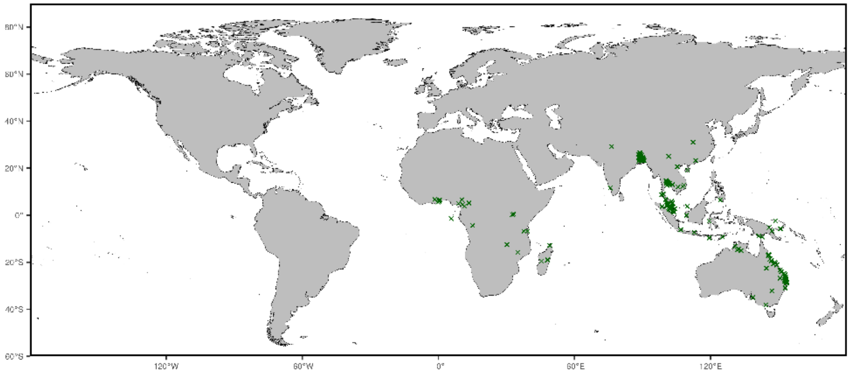


NOTE: centroids of polygons were used to map administrative and buffered occurrences, where relevant

Precision weightings

| **Precision categories** | **Precision weightings** |
|---|---|
| Prior cases | 0.4904 |
| Niche classifications (0.2) | 0.5511 |
| Niche classifications (0.4) | 0.6562 |
| Niche classifications (0.6) | 0.7335 |
| Niche classifications (0.8) | 0.7807 |
| All diagnosed infections | 0.1271 |
| Non-experimental infections | 0.1271 |
| Wild infections | 0.1228 |
| Wild/PCR+ infections | 0.1272 |

### Data sources

Evidence data for all Henipaviruses were acquired from three main sources. First, we obtained a list of countries with historic transmission reports of both Nipah and Hendra compiled by the WHO; we supplemented this list with all human non-specific Henipavirus cases from the comprehensive literature review compiled in the unpublished Pigott et al. manuscript "Mapping the environmental suitability for the Henipaviruses: Hendra and Nipah". Second, an environmental suitability synoptic niche map with continuous suitability estimates from the unpublished research by Pigott et al. was used to create evidence suitability layers for all Henipaviruses with cutoff scores of 0.2, 0.4, 0.6, and 0.8. This suitability layer was constructed using human and mammalian occurrence data through 2017 and relevant environmental covariates (namely, enhanced vegetation index, cumulative precipitation, mean temperature, tasseled cap brightness, tasseled cap wetness, and a consolidated indicator of species distribution models of implicated bat species) to predict the estimated environmental suitability for Henipaviral transmission. Third, non-human species with evidence of Nipah, Hendra or any non-specific Henipavirus as reported to GIDEON were used in creating a zoonotic transmission layer, and these species were stratified into four groupings: all reported species, all non-experimental reports, all wild infections, and all wild, PCR positive infections. The geographic range for each species was obtained from the International Union for Conservation of Nature's (IUCN) Red List of Threatened Species.

Occurrence data for obtaining precision estimates from each layer were obtained from unpublished Pigott et al. and were collated using a comprehensive, systematic review and data extraction process.

| Evidence category | Citation |
|---|---|
| **Countries with reported human cases** | World Health Organization https://www.who.int/news-room/fact-sheets/detail/nipah-virus and https://www.who.int/health-topics/hendra-virus-disease#tab=tab_1 and Pigott et al. *Unpublished.* "Mapping the environmental suitability for the Henipaviruses: Hendra and Nipah" |
| **Environmental suitability** | Pigott et al. *Unpublished.* "Mapping the environmental suitability for the Henipaviruses: Hendra and Nipah" |
| **Reservoir status determination** | Berger. (2005) "GIDEON: a comprehensive Web-based resource for geographic medicine" Int. J. Health Geogr. 4, 10 |

| | |
|---|---|
| **Human geopositioned occurrences** | Pigott et al. *Unpublished.* "Mapping the environmental suitability for the Henipaviruses: Hendra and Nipah" |

### Pathogen-specific Evidence

#### Lassa fever

Overall assessment

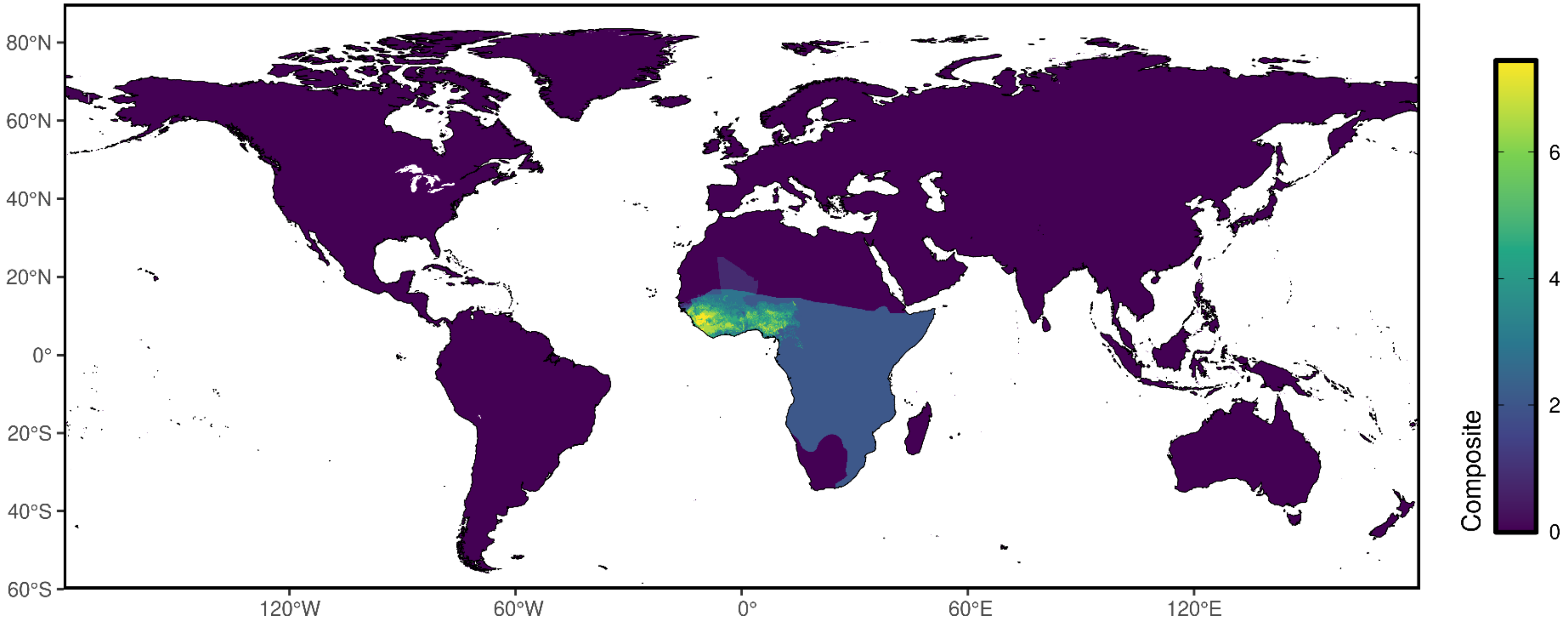

### Reservoir presence

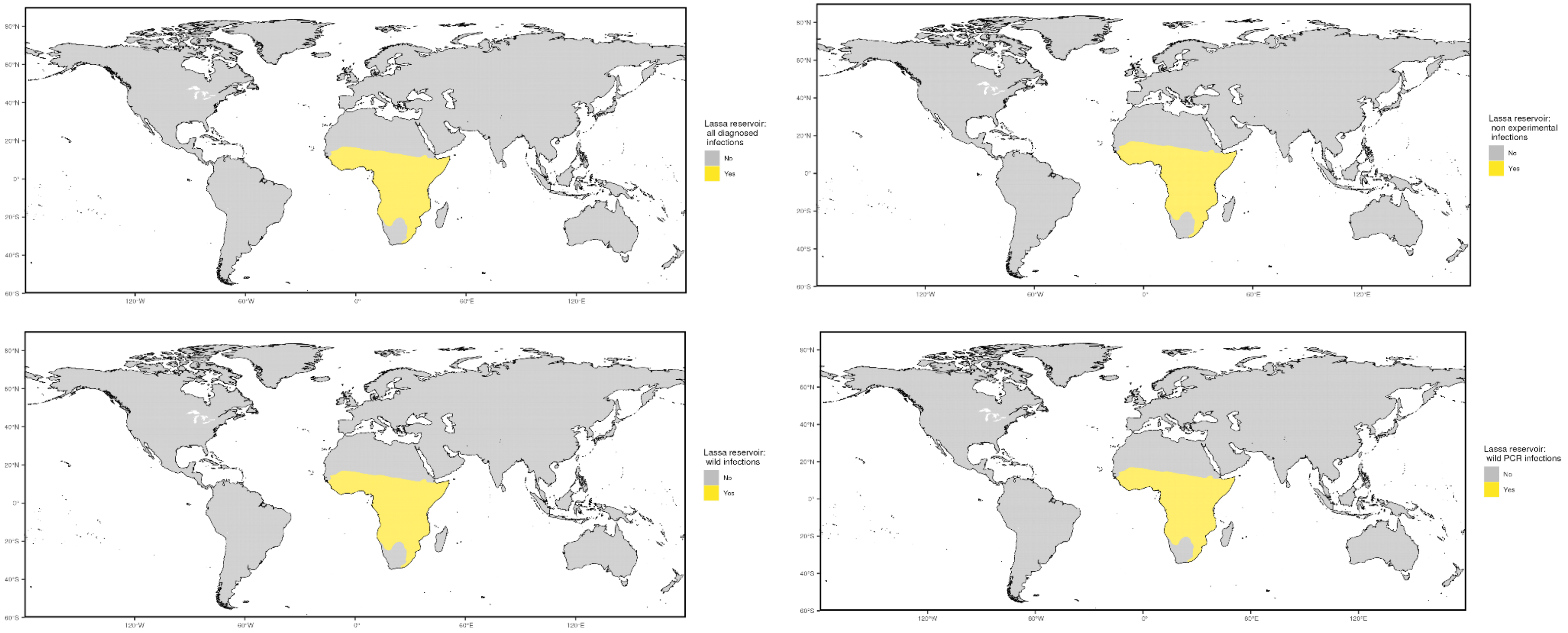

#### Niche classifications

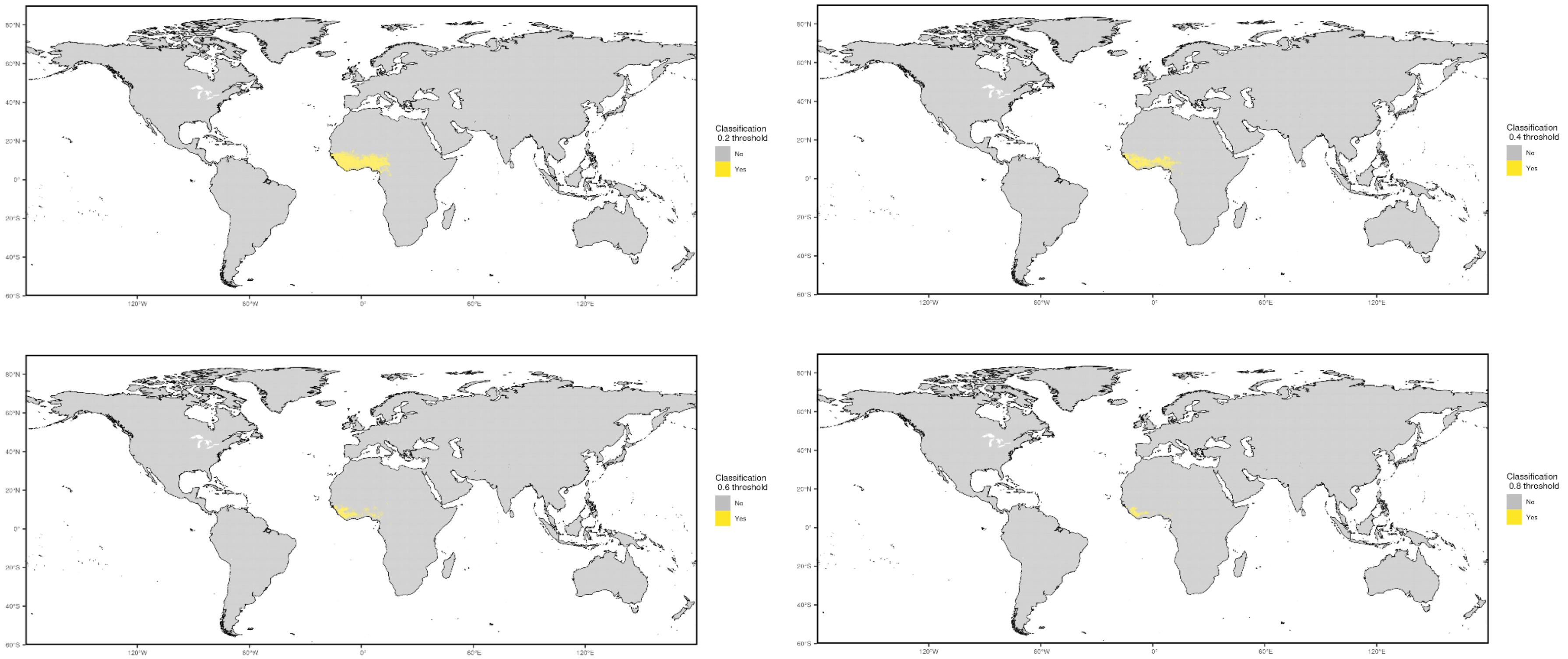

Seroprevalence

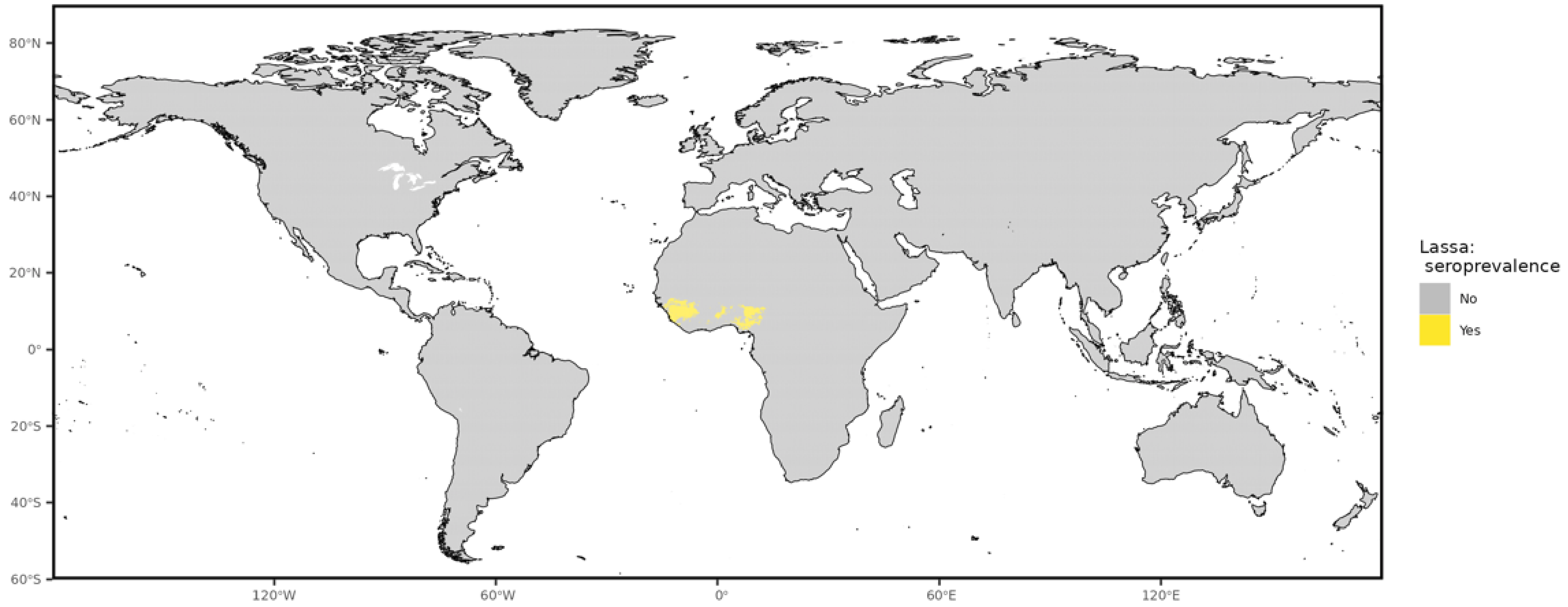

Prior cases

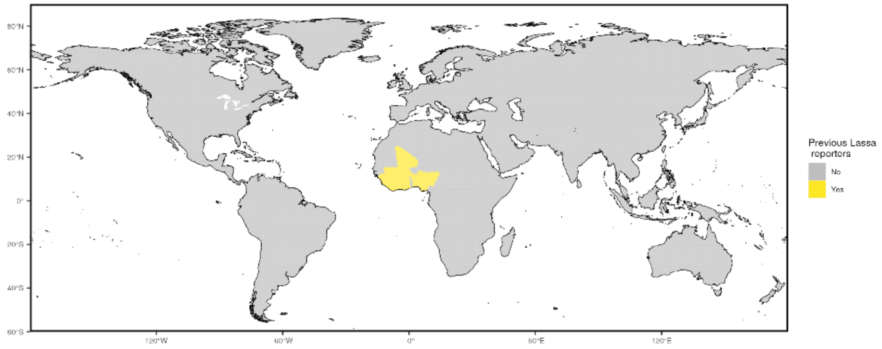


Occurrence data

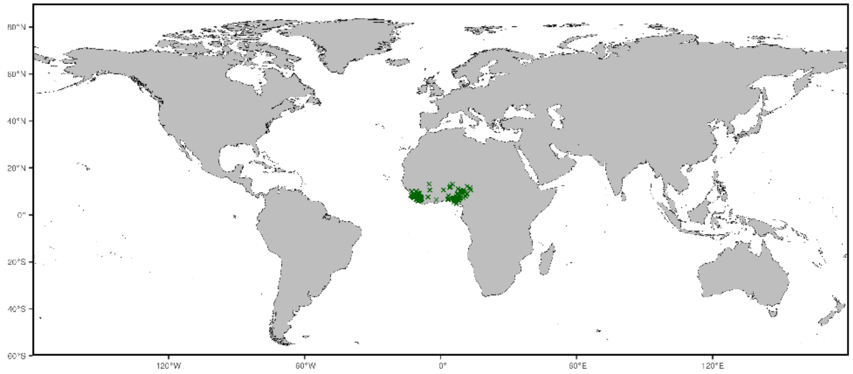


NOTE: centroids of polygons were used to map administrative and buffered occurrences, where relevant

#### Precision weightings

| Precision categories | Precision weightings |
|---|---|
| Prior cases | 0.8208 |
| Niche classifications (0.2) | 0.8532 |
| Niche classifications (0.4) | 0.8797 |
| Niche classifications (0.6) | 0.9364 |
| Niche classifications (0.8) | 0.9766 |
| High seroprevalence | 0.9369 |
| All diagnosed infections | 0.5117 |
| Non-experimental infections | 0.5117 |
| Wild infections | 0.5117 |
| Wild/PCR+ infections | 0.5117 |

#### Data sources

Evidence data for Lassa fever disease were acquired from four main sources. First, we obtained a list of countries with historic transmission reports of Lassa fever compiled by the CDC. Second, an environmental suitability synoptic niche map with continuous suitability estimates from Mylne et al. was used to create evidence suitability layers for Lassa fever with cutoff scores of 0.2, 0.4, 0.6, and 0.8. This suitability layer was constructed using human and mammalian occurrence data and relevant environmental covariates (namely, enhanced vegetation index, land surface temperature, elevation, potential evapotranspiration, and a covariate of the implicated *Mastomys natalensis*) to predict the estimated environmental suitability for Lassa transmission. Third, a high human seroprevalence layer was created by utilizing two primary datasets published by Basinski et al. in 2021: reported human serosurvey data and an estimated risk layer comprising of three distinct elements, incorporating data from human lassa serosurveys, generated predictions of human lassa seroprevalence, and incidence throughout West Africa. We used the human serosurvey studies to calculate the 75th percentile threshold value of reported proportion seropositives, indicating “high” seropositivity among studies. For the coordinates of these high seropositivity studies, we extracted the values of the estimated seroprevalence raster by Basinski et al. 2021, to determine a floor threshold value that if applied to the seroprevalence raster would produce a map that included 75% of all “high” seroprevalence studies. This high seroprevalence threshold (0.1106193) was then used to convert the raster values of the Combined_Risk_Layer into a binary format, resulting in the creation of a high human seroprevalence layer used in this analysis. Finally, the mammalian non-human species with evidence of Lassa fever infections (*Mastomys natalensis*) was used in creating a zoonotic transmission layer stratified into four groupings: all reported species, all non-experimental reports, all wild infections, and all wild, PCR positive infections. The geographic range for *Mastomys natalensis* was obtained from the International Union for Conservation of Nature’s (IUCN) Red List of Threatened Species.

Occurrence data for obtaining precision estimates from each layer were obtained from Mylne et al. (2015) and were collated using a comprehensive, systematic review and data extraction process.

| Evidence category | Citation |
|---|---|
| **Countries with reported human cases** | US Centers for Disease Control - https://www.cdc.gov/vhf/lassa/outbreaks/index.html |
| **Environmental suitability** | Mylne et al. (2015) "Mapping the zoonotic niche of Lassa fever in Africa" Trans Roy Soc Trop Med Hyg https://www.ncbi.nlm.nih.gov/pmc/articles/PMC4501400/ |

| | |
|---|---|
| **Reservoir status determination** | 1.McCormick JB, Webb PA, Krebs JW et al.. A prospective study of the epidemiology and ecology of Lassa fever. J Infect Dis 1987;155:437–44.<br><br>2. Safronetz D, Lopez JE, Sogoba N et al.. Detection of Lassa virus, Mali. Emerg Infect Dis 2010;16:1123–6.<br><br>3. Wulff H, Fabiyi A, Monath TP. Recent isolations of Lassa virus from Nigerian rodents. Bull World Health Organ 1975;52:609–13. |
| **Human geopositioned occurrences** | Mylne et al. (2015) "Mapping the zoonotic niche of Lassa fever in Africa" Trans Roy Soc Trop Med Hyg https://www.ncbi.nlm.nih.gov/pmc/articles/PMC4501400/ |

### Pathogen-specific Evidence

#### Marburg virus

Overall assessment

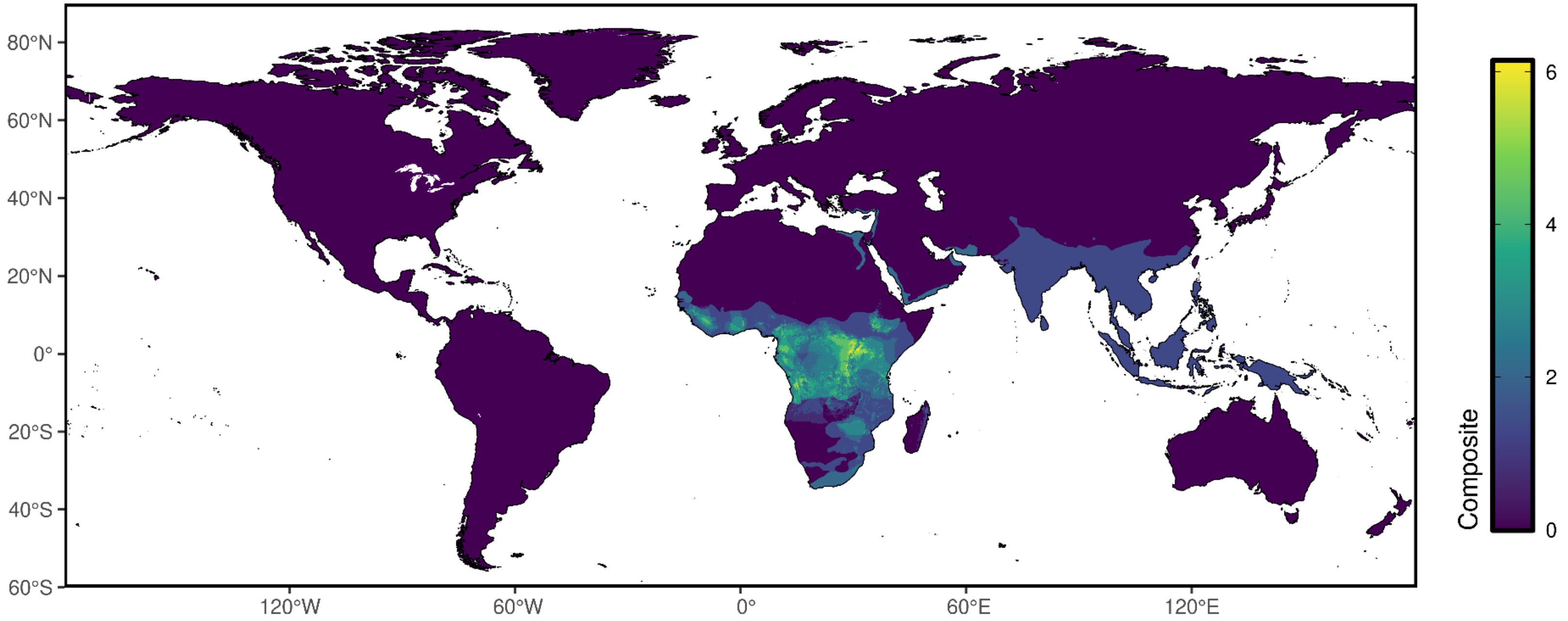

### Reservoir presence

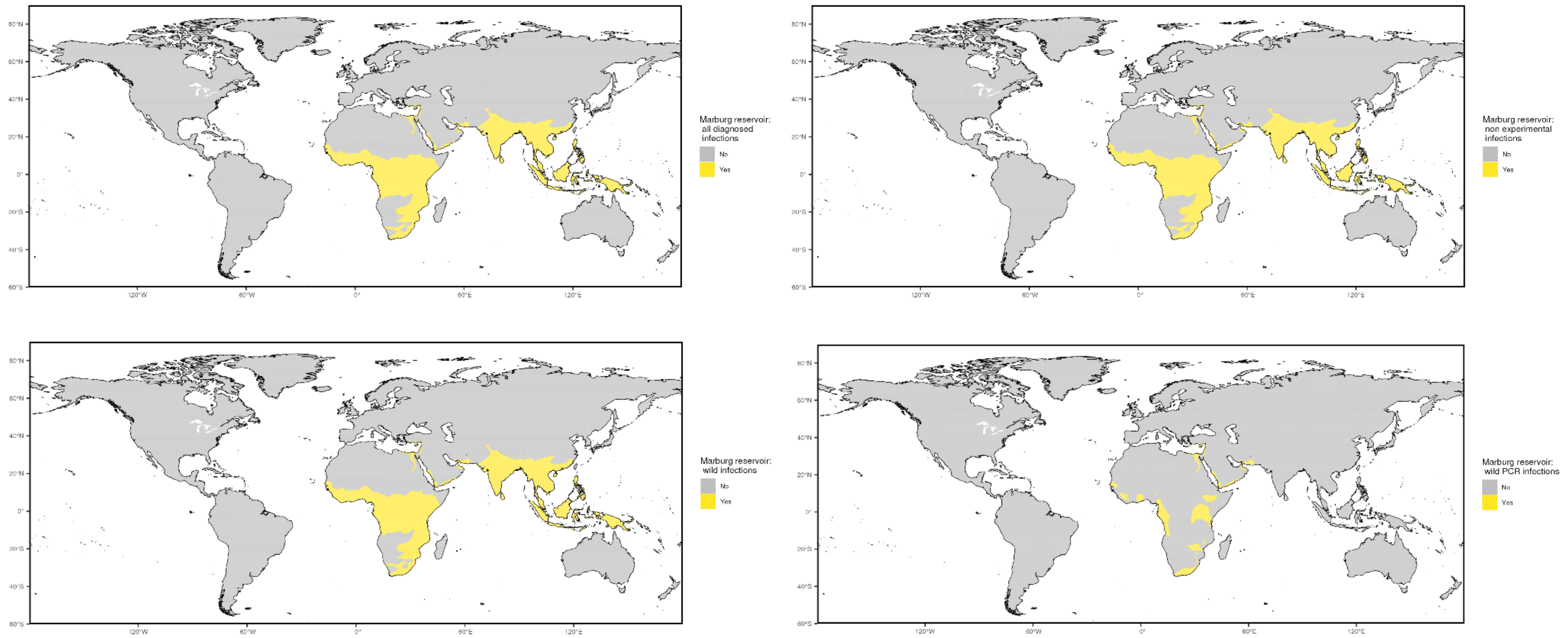

### Niche classifications

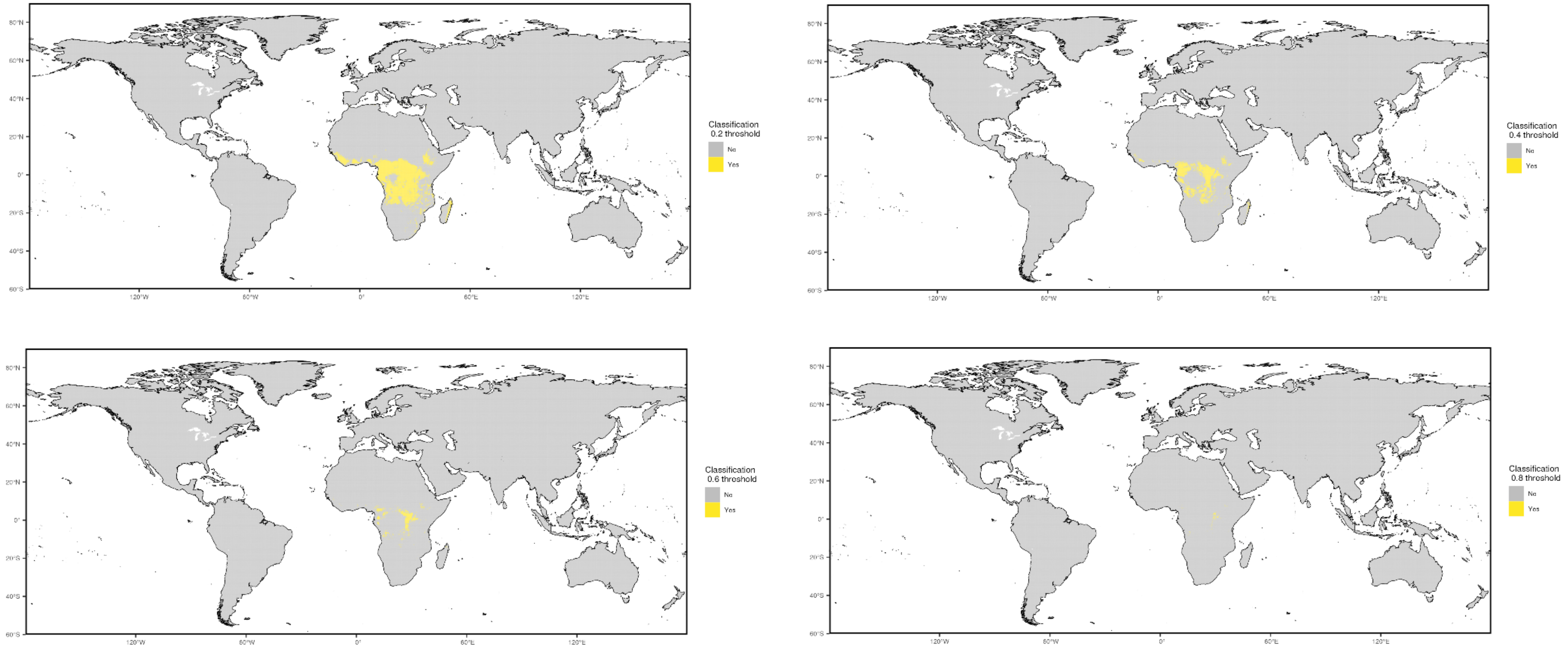

#### Prior cases

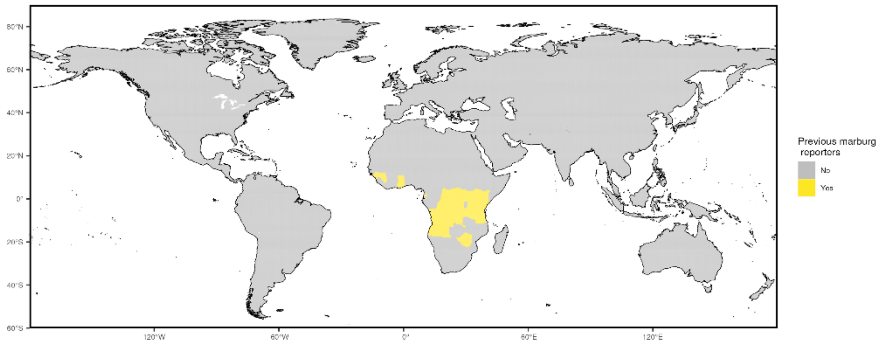


#### Occurrence data

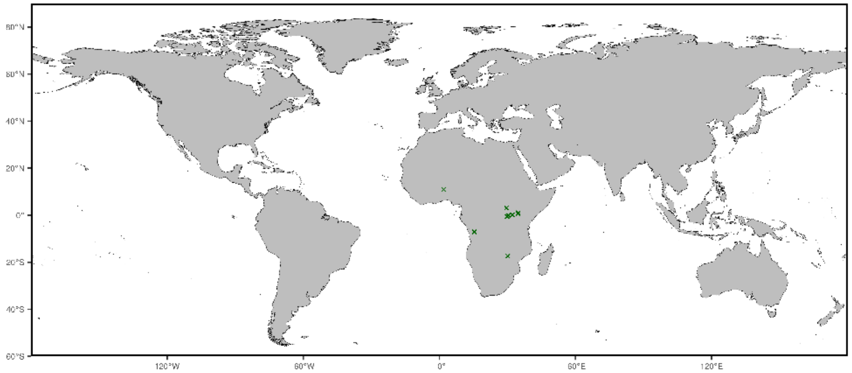


NOTE: centroids of polygons were used to map administrative and buffered occurrences, where relevant

#### Precision weightings

| **Precision categories** | **Precision weightings** |
|---|---|
| Prior cases | 0.6897 |
| Niche classifications (0.2) | 0.6452 |
| Niche classifications (0.4) | 0.8261 |
| Niche classifications (0.6) | 0.8889 |
| Niche classifications (0.8) | 1 |
| All diagnosed infections | 0.4468 |
| Non-experimental infections | 0.4468 |
| Wild infections | 0.4468 |
| Wild/PCR+ infections | 0.75 |

#### Data sources

Evidence data for Marburg virus disease were acquired from three main sources. First, we obtained a list of countries with historic transmission reports of Marburg compiled by the CDC. Second, an environmental suitability synoptic niche map with continuous suitability estimates from Pigott et al. was used to create evidence suitability layers for Marburg with cutoff scores of 0.2, 0.4, 0.6, and 0.8. This suitability layer was constructed using human and mammalian occurrence data and relevant environmental covariates (namely, enhanced vegetation index, land surface temperature, elevation, potential evapotranspiration, and distance to karst landforms) to predict the estimated environmental suitability for Marburg transmission. Third, mammalian non-human species with evidence of Marburg infections, identified by Han et al. (2016) were used in creating a zoonotic transmission layer, and these species were stratified into four groupings: all reported species, all non-experimental reports, all wild infections, and all wild, PCR positive infections. The geographic range for each species was obtained from the International Union for Conservation of Nature's (IUCN) Red List of Threatened Species.

Occurrence data for obtaining precision estimates from each layer were obtained from Pigott et al. (2015), with additions of three outbreaks since publication, namely the outbreaks in Ghana, Equatorial Guinea, and Tanzania

| Evidence category | Citation |
|---|---|
| **Countries with reported human cases** | US Centers for Disease Control - https://www.cdc.gov/vhf/marburg/outbreaks/chronology.html |
| **Environmental suitability** | Pigott et al. (2015) "Mapping the zoonotic niche of Marburg virus disease in Africa" Trans R Soc Trop Med Hyg 109(6) 366-78 |
| **Reservoir status determination** | Han et al. (2016) "Undiscovered Bat Hosts of Filoviruses" PLoS NTDs 10(7) e0004815 |
| **Human geopositioned occurrences** | Pigott et al. (2015) "Mapping the zoonotic niche of Marburg virus disease in Africa" Trans R Soc Trop Med Hyg 109(6) 366-78 with supplemental additions of details of index case in Ghana, Equatorial Guinea, and Tanzania |

### Pathogen-specific Evidence

#### Middle East respiratory syndrome

Overall assessment

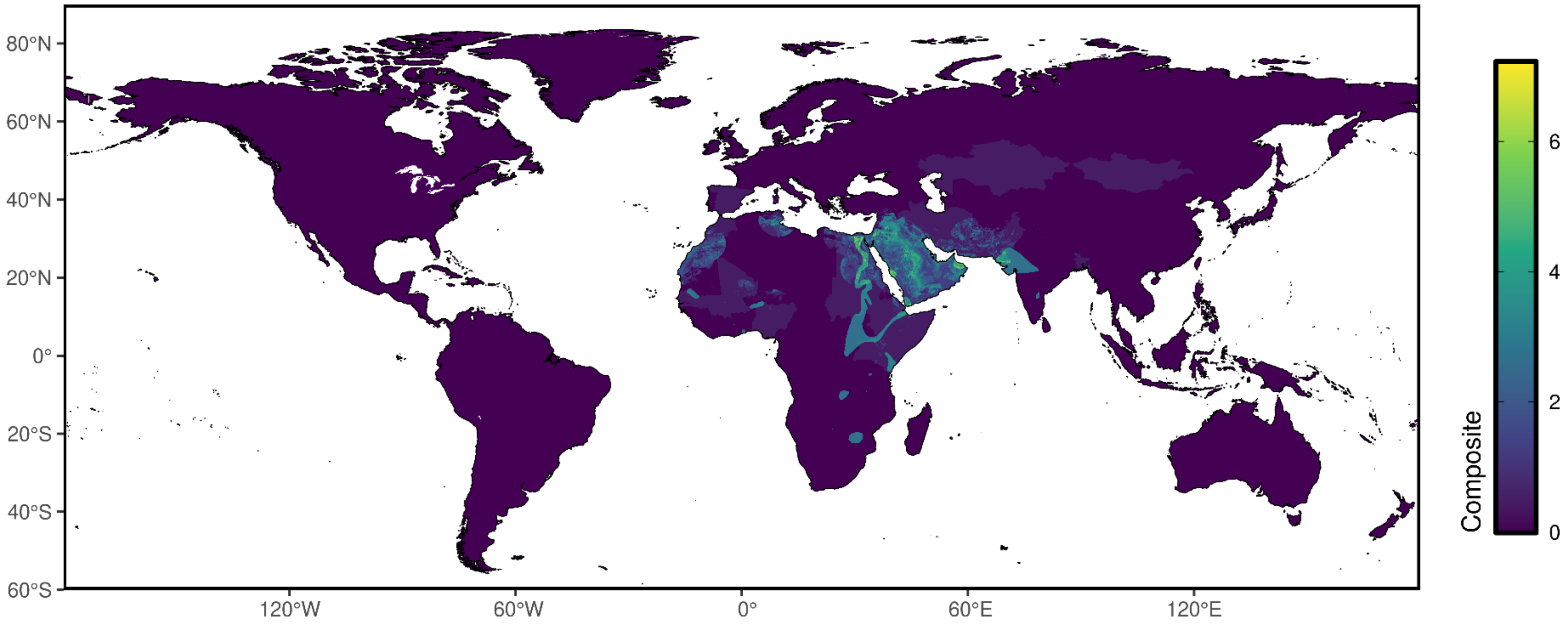

### Reservoir presence

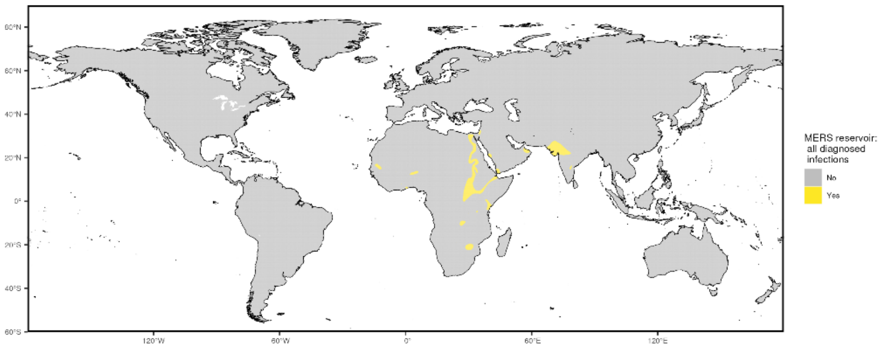


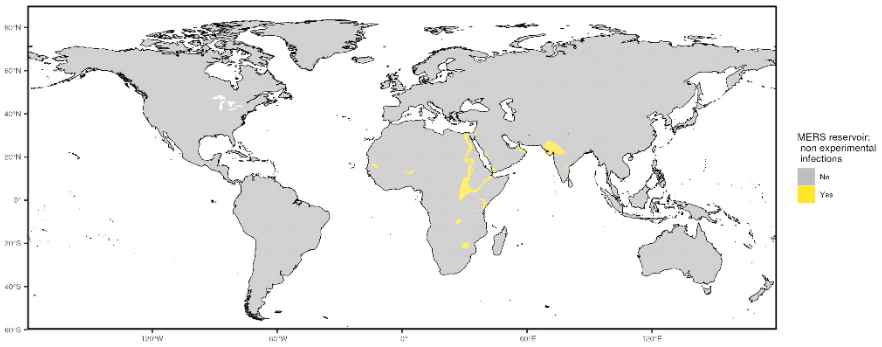


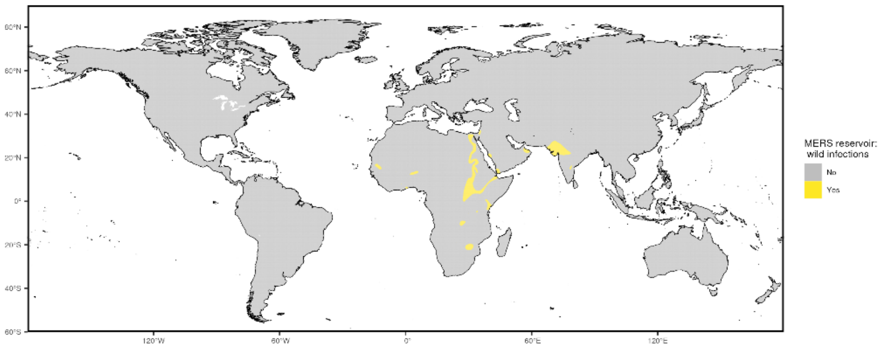


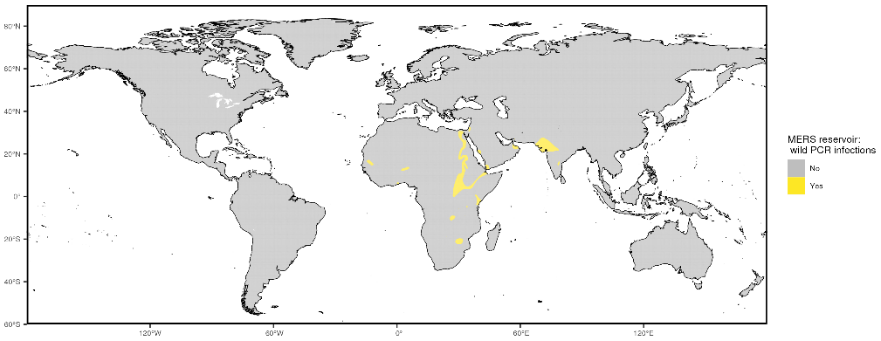

### Niche classifications

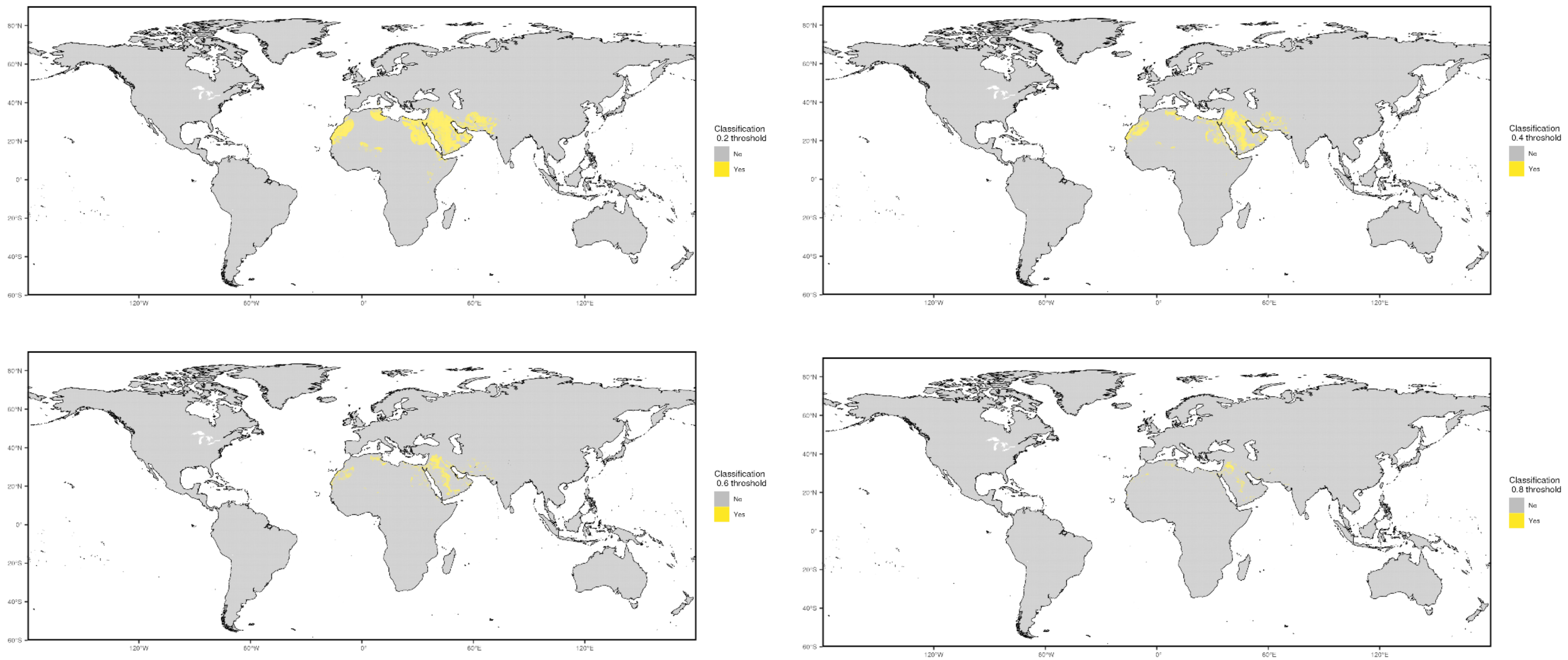

#### Prior cases

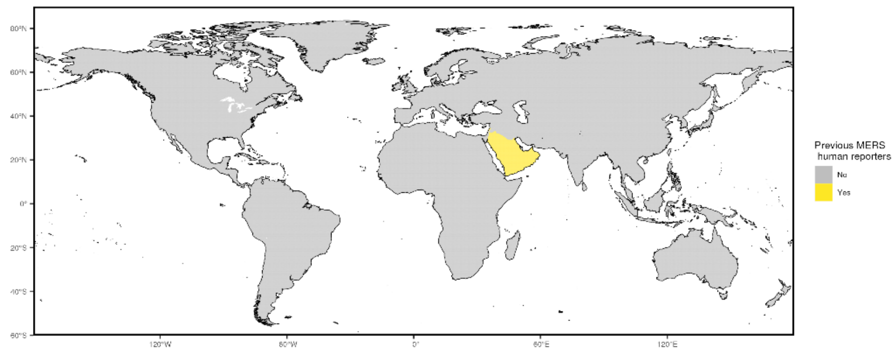


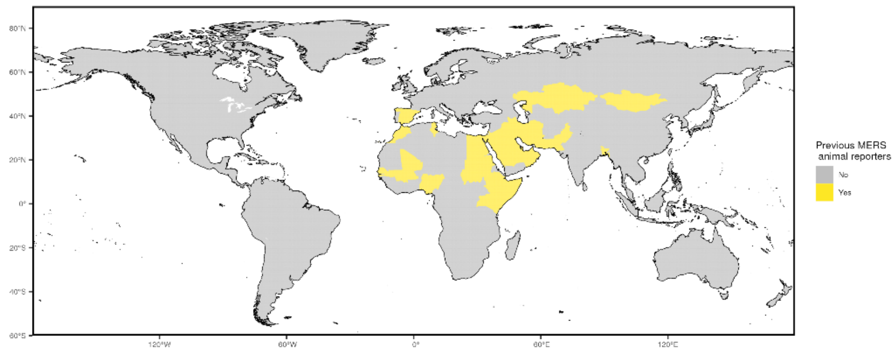


#### Occurrence

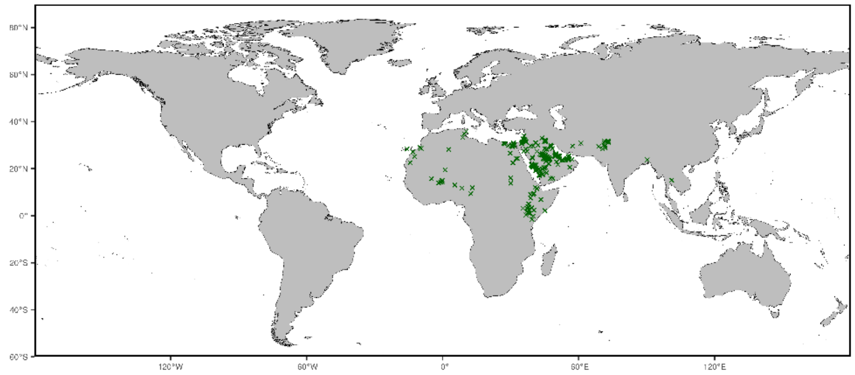


NOTE: centroids of polygons were used to map administrative and buffered occurrences, where relevant

Precision weightings

| **Precision categories** | **Precision weightings** |
|---|---|
| Prior cases- humans | 0.8442 |
| Prior cases- animals | 0.4767 |
| Niche classifications (0.2) | 0.7076 |
| Niche classifications (0.4) | 0.7749 |
| Niche classifications (0.6) | 0.8273 |
| Niche classifications (0.8) | 0.9083 |
| All diagnosed infections | 0.669 |
| Non-experimental infections | 0.669 |
| Wild infections | 0.669 |
| Wild/PCR+ infections | 0.669 |

#### Data sources

Evidence data for MERS-CoV were acquired from three main sources. First, we obtained a list of countries with historic transmission reports of MERS-CoV in humans compiled by the WHO. We also obtained a list of countries with historic transmission reports of MERS-CoV in humans compiled by the WHO and FAO. Second, an environmental suitability synoptic niche map with continuous suitability estimates from unpublished research by Osborne et al. was used to create evidence suitability layers for MERS-CoV with cutoff scores of 0.2, 0.4, 0.6, and 0.8. This suitability layer was constructed using human and mammalian occurrence data through February 2018 and relevant environmental covariates (namely, enhanced vegetation index, aridity, tasseled capped brightness, monthly averaged daily mean temperature, and tasseled cap wetness, elevation, and camel head count per hectare) to predict the estimated environmental suitability for MERS-CoV transmission. Third, mammalian non-human species with evidence of MERS-CoV as reported to GIDEON were used in creating a zoonotic transmission layer, and these species were stratified into four groupings: all reported species, all non-experimental reports, all wild infections, and all wild, PCR positive infections. The geographic range for each species was obtained from the International Union for Conservation of Nature's (IUCN) Red List of Threatened Species.

Occurrence data for obtaining precision estimates from each layer were obtained from Ramshaw et al. (2019) "A database of geopositioned Middle East Respiratory Syndrome Coronavirus occurrences" and the niche suitability modeling from Osborne et al. and were collated using a comprehensive, systematic review and data extraction process.

| Evidence category | Citation |
|---|---|
| **Countries with reported human cases** | World Health Organization Internal Database |
| **Countries with reported animal cases** | World Health Organization and Food and Agricultural Organization Internal Databases |
| **Environmental suitability** | Osborne, J.C.P. et al. *Unpublished*. "Mapping the potential geographic extent of Middle East respiratory syndrome coronavirus." |

| | |
|---|---|
| **Reservoir status determination** | Berger. (2005) “GIDEON: a comprehensive Web-based resource for geographic medicine” Int. J. Health Geogr. 4, 10 |
| **Human geopositioned occurrences** | Ramshaw RE, Letourneau ID, Hong AY, Hon J, Morgan JD, Osborne JCP, Shirude S, Van Kerkhove MD, Hay SI, Pigott DM. A database of geopositioned Middle East Respiratory Syndrome Coronavirus occurrences. Sci Data. 2019 Dec 13;6(1):318. doi: 10.1038/s41597-019-0330-0. PMID: 31836720; PMCID: PMC6911100 and Osborne, J.C.P. et al. *Unpublished*. “Mapping the potential geographic extent of Middle East respiratory syndrome coronavirus.” |

### Pathogen-specific Evidence

#### Monkeypox

Overall assessment

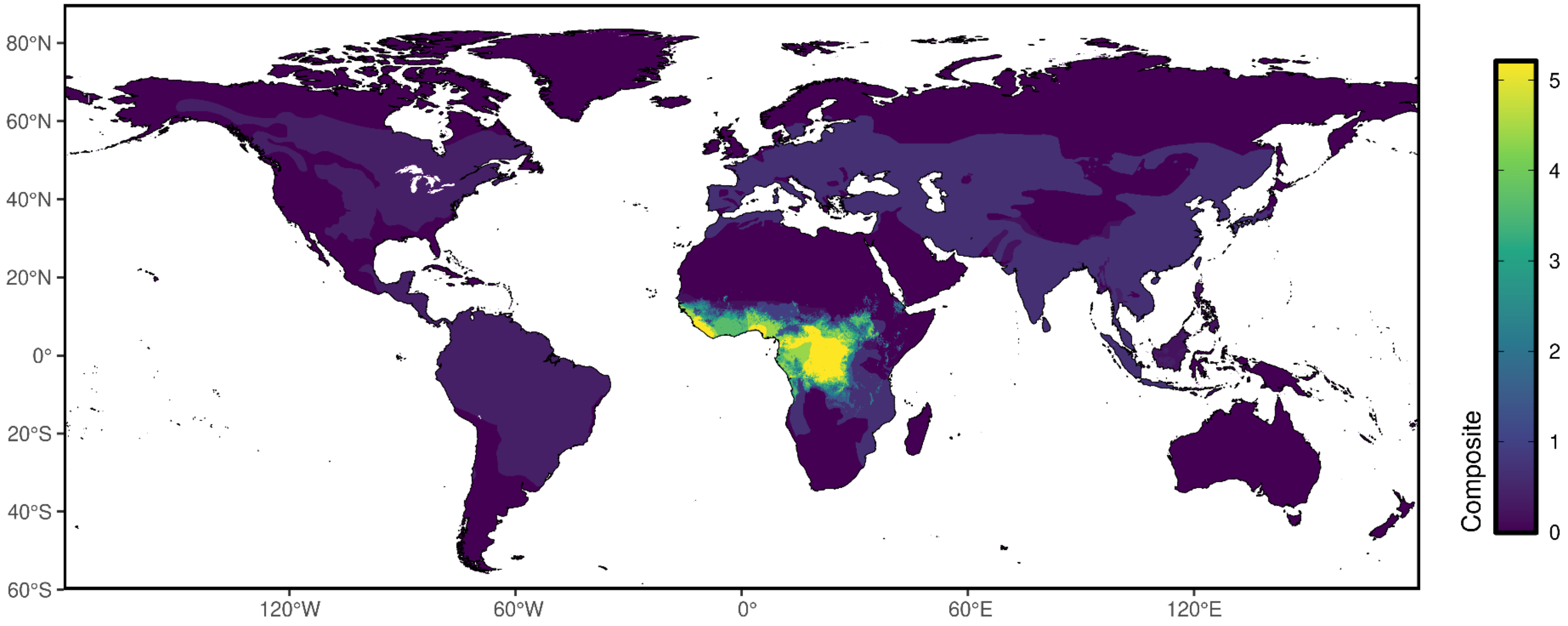

### Reservoir presence

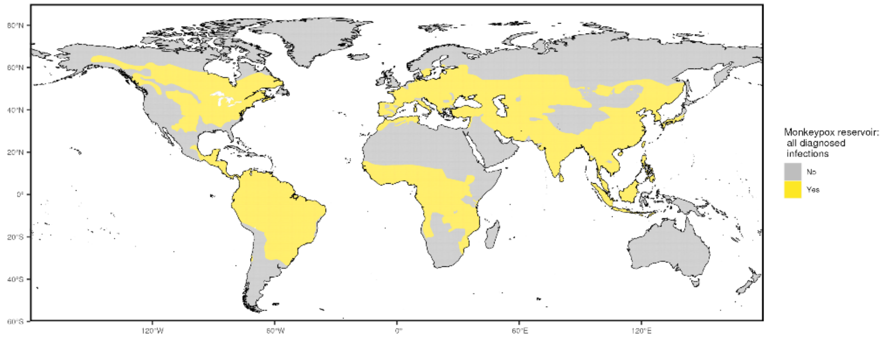


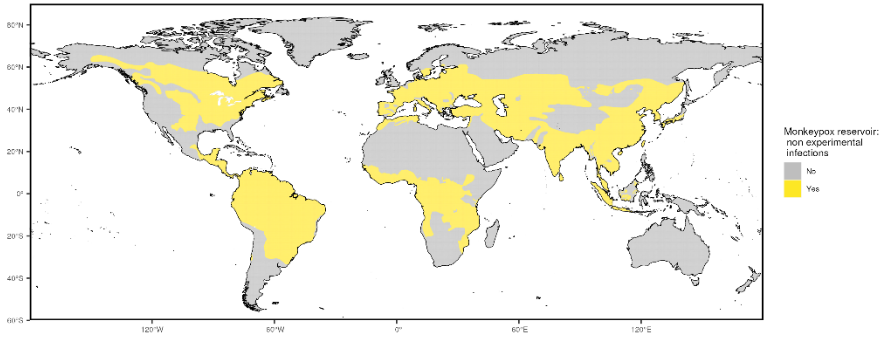


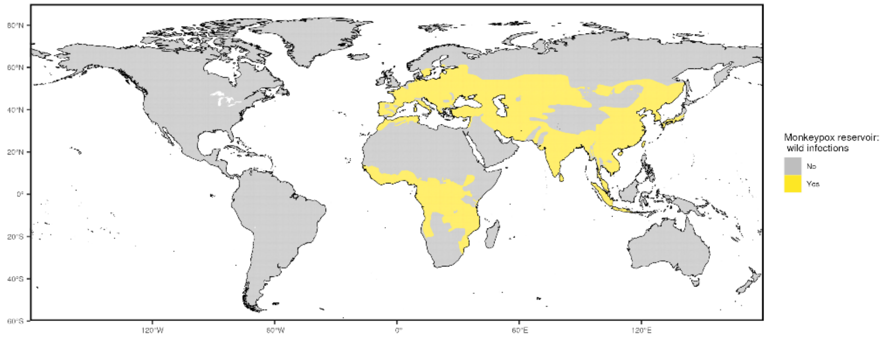


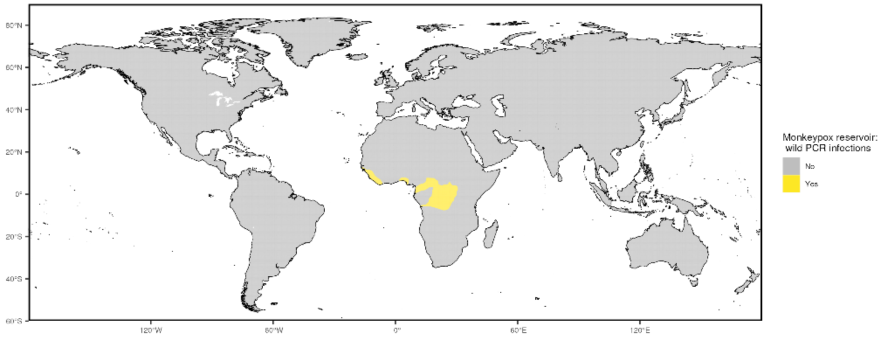

### Niche classifications

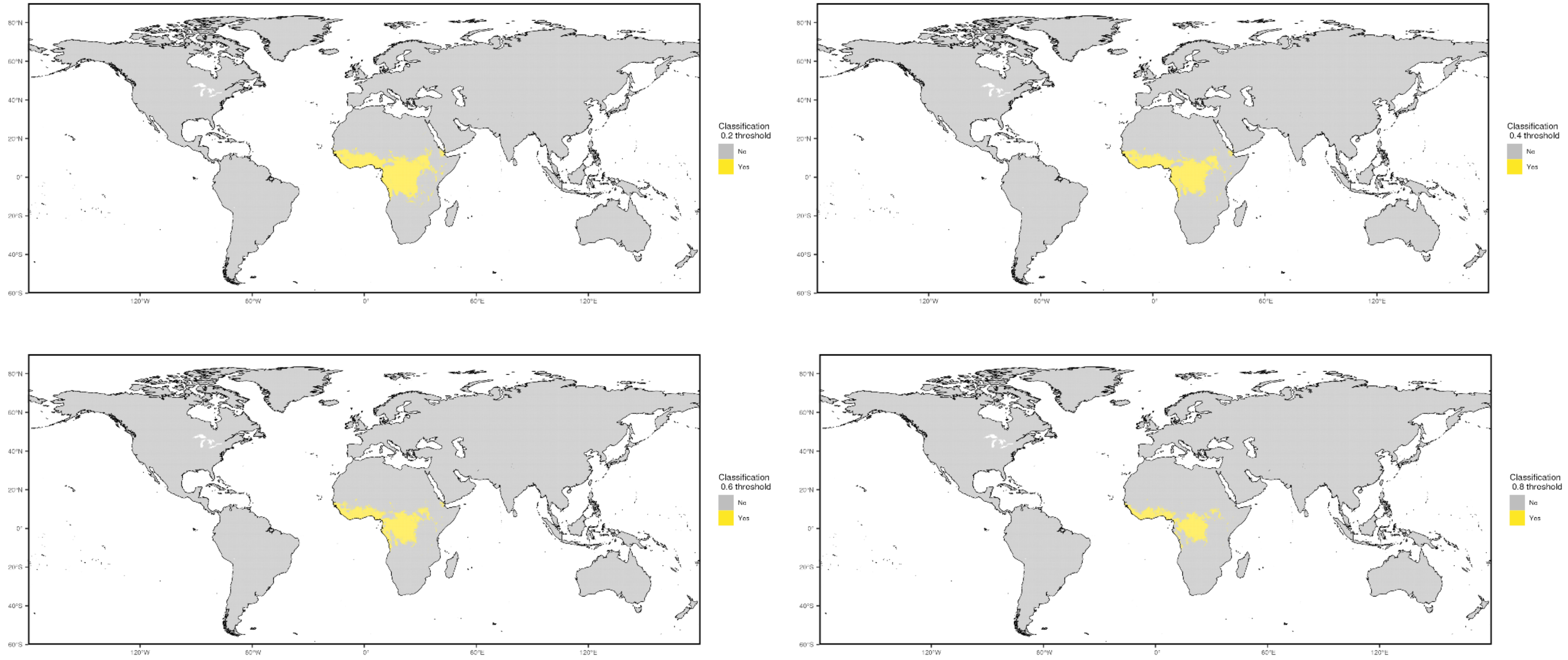

#### Prior cases

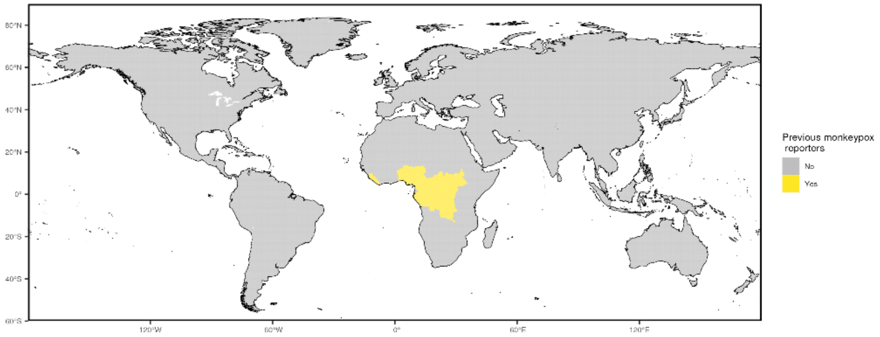


#### Occurrence data

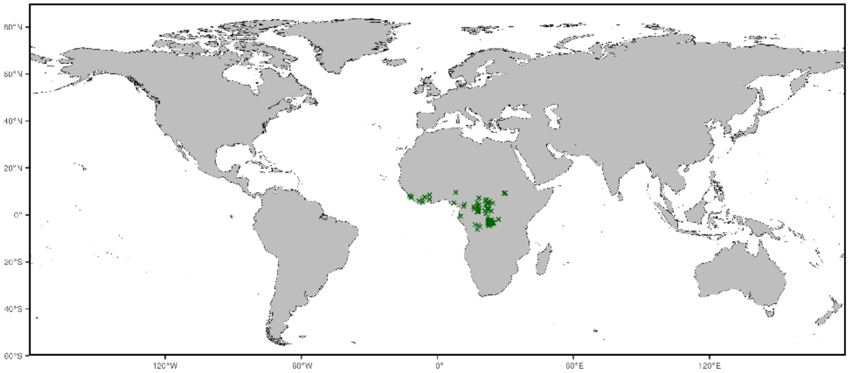


NOTE: centroids of polygons were used to map administrative and buffered occurrences, where relevant

#### Precision weightings

| Precision categories | Precision weightings |
|---|---|
| Prior cases | 0.7452 |
| Niche classifications (0.2) | 0.691 |
| Niche classifications (0.4) | 0.7248 |
| Niche classifications (0.6) | 0.7508 |
| Niche classifications (0.8) | 0.7964 |
| All diagnosed infections | 0.1884 |
| Non-experimental infections | 0.1948 |
| Wild infections | 0.2864 |
| Wild/PCR+ infections | 0.8253 |

#### Data sources

Evidence data for mpox were acquired from three main sources. First, we obtained a list of countries with historic transmission reports of mpox compiled by the WHO. Second, an environmental suitability synoptic niche map with continuous suitability estimates from unpublished research by Hulland et al. was used to create evidence suitability layers for mpox with cutoff scores of 0.2, 0.4, 0.6, and 0.8. This suitability layer was constructed using human and mammalian occurrence data through 2019 and relevant environmental covariates (namely, monthly precipitation, monthly averaged daily mean temperature, monthly averaged daily minimum temperature, monthly averaged daily maximum temperature, monthly averaged diurnal temperature range, enhanced vegetation index, tasseled cap wetness, tasseled cap brightness, elevation, and slope) to predict the estimated environmental suitability for mpox transmission for a synoptic year 2016. Third, mammalian non-human species with evidence of mpox as reported to GIDEON were used in creating a zoonotic transmission layer, and these species were stratified into four groupings: all reported species, all non-experimental reports, all wild infections, and all wild, PCR positive infections. The geographic range for each species was obtained from the International Union for Conservation of Nature's (IUCN) Red List of Threatened Species.

Occurrence data for obtaining precision estimates from each layer were obtained from unpublished Hulland et. al. and Ramshaw et al. and were collated using a comprehensive, systematic review and data extraction process.

| **Evidence category** | **Citation** |
|---|---|
| **Countries with reported human cases** | McCollum, A.M. et al. (2023) "Epidemiology of human monkeypox (mpox) – worldwide, 2018–2021" Weekly Epidemiological Record, No 3, 20 January 2023. https://apps.who.int/iris/bitstream/handle/10665/365630/WER9803-29-36.pdf?sequence=1&isAllowed=y |
| **Environmental suitability** | Hulland, E. N. et al. *Unpublished*. "Mapping the environmental suitability of monkeypox in humans across Africa." |
| **Reservoir status determination** | Berger. (2005) "GIDEON: a comprehensive Web-based resource for geographic medicine" Int. J. Health Geogr. 4, 10 |
| **Human geopositioned occurrences** | Ramshaw, R. E. et al. *Under review.* "Global monkeypox occurrences in the literature, 1972–2019: a geo-tagged database." and Hulland, E. N. et al. *Unpublished*. "Mapping the environmental suitability of monkeypox in humans across Africa." |

### Pathogen-specific Evidence

#### Nipah virus

Overall assessment

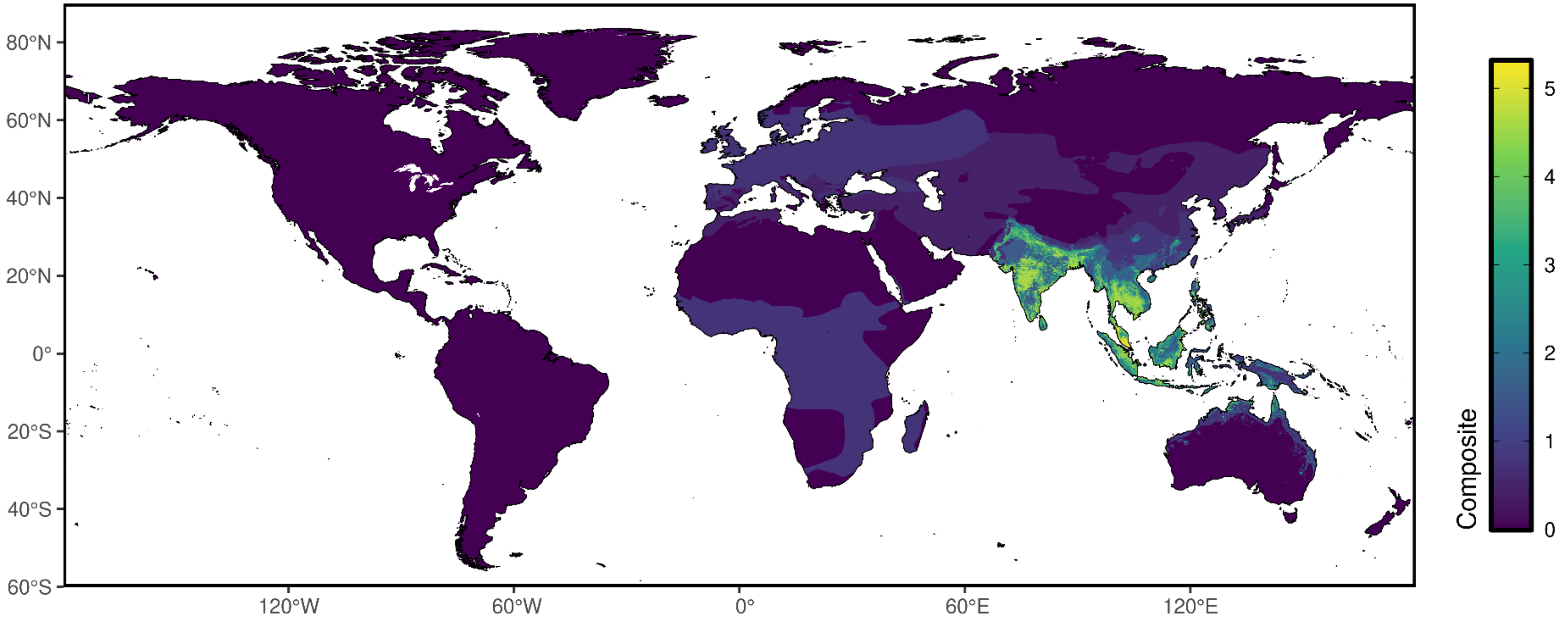

### Reservoir presence

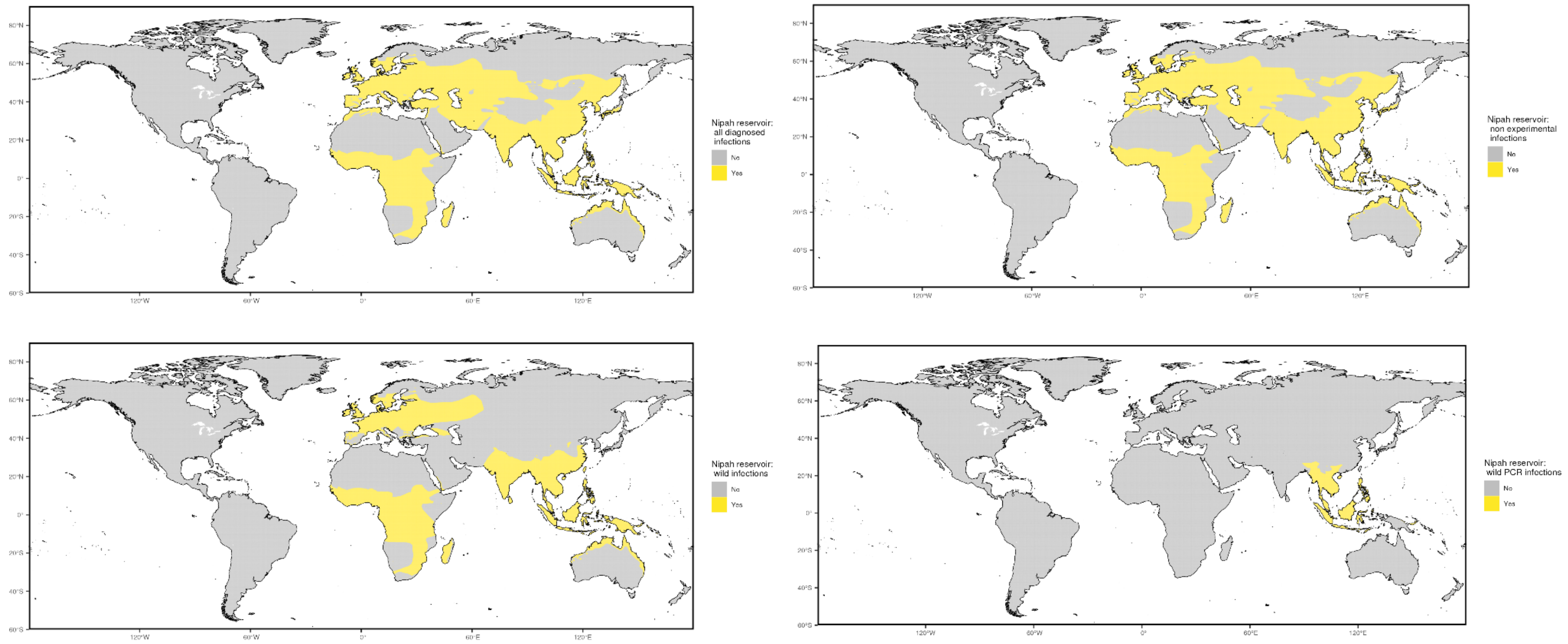

### Niche classifications

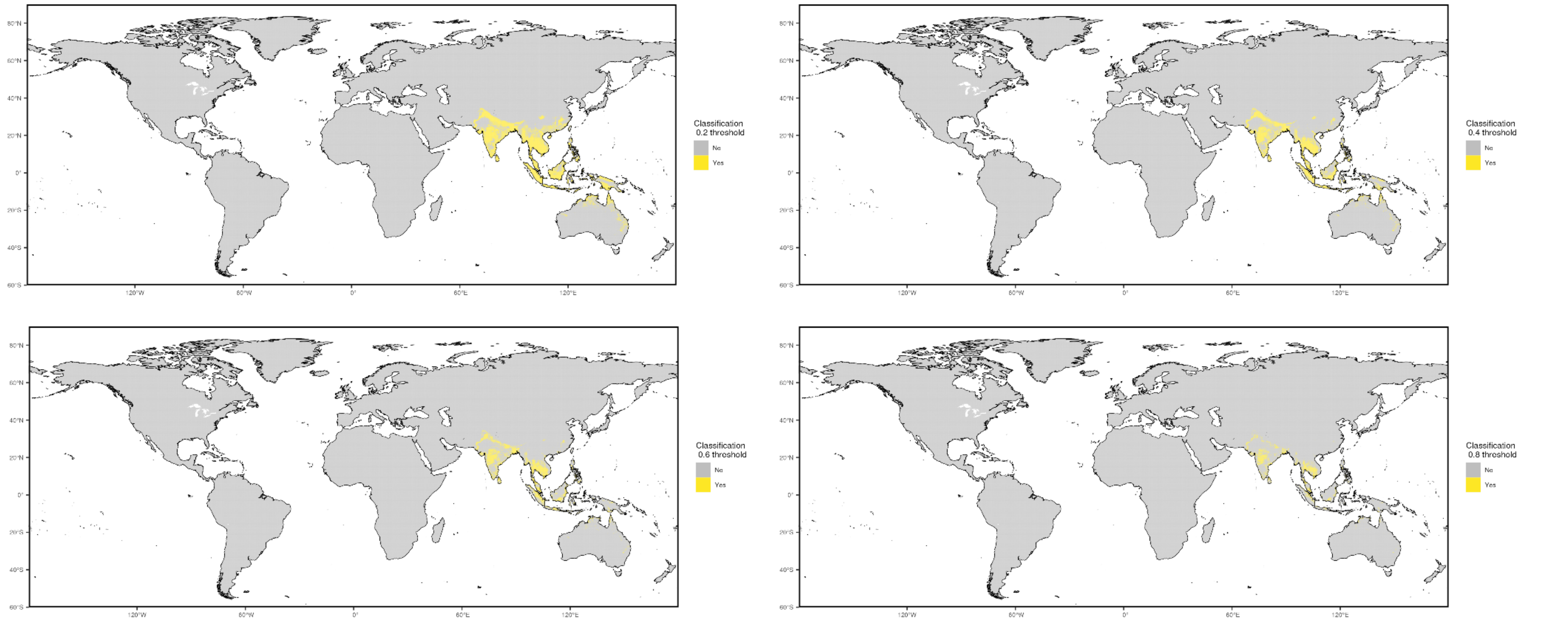

#### Prior cases

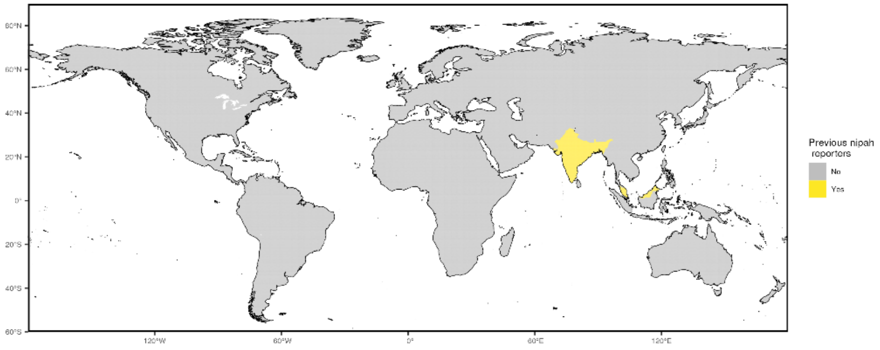


#### Occurrence data

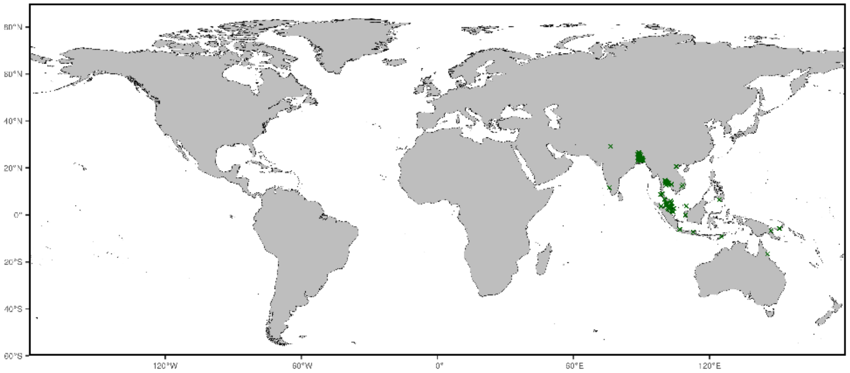


NOTE: centroids of polygons were used to map administrative and buffered occurrences, where relevant

#### Precision weightings

| **Precision categories** | **Precision weightings** |
|---|---|
| Prior cases | 0.7551 |
| Niche classifications (0.2) | 0.6849 |
| Niche classifications (0.4) | 0.7614 |
| Niche classifications (0.6) | 0.8166 |
| Niche classifications (0.8) | 0.8785 |
| All diagnosed infections | 0.2206 |
| Non-experimental infections | 0.2206 |
| Wild infections | 0.2977 |
| Wild/PCR+ infections | 0.6719 |

#### Data sources

**Nipah**
Evidence data for Nipah were acquired from three main sources. First, we obtained a list of countries with historic transmission reports of Nipah compiled by the WHO. Second, an environmental suitability synoptic niche map with continuous suitability estimates from unpublished research by Pigott et al. was used to create evidence suitability layers for Nipah virus with cutoff scores of 0.2, 0.4, 0.6, and 0.8. This suitability layer was constructed using human and mammalian occurrence data through 2017 and relevant environmental covariates (namely, enhanced vegetation index, cumulative precipitation, mean temperature, tasseled cap brightness, tasseled cap wetness, and a consolidated indicator of species distribution models of implicated bat species) to predict the estimated environmental suitability for Nipah transmission for a synoptic year 2016. Third, non-human species with evidence of Nipah as reported to GIDEON were used in creating a zoonotic transmission layer, and these species were stratified into four groupings: all reported species, all non-experimental reports, all wild infections, and all wild, PCR positive infections. The geographic range for each species was obtained from the International Union for Conservation of Nature's (IUCN) Red List of Threatened Species.

Occurrence data for obtaining precision estimates from each layer were obtained from unpublished Pigott et al. and were collated using a comprehensive, systematic review and data extraction process.

| Evidence category | Citation |
|---|---|
| **Countries with reported human cases** | World Health Organization https://www.who.int/news-room/fact-sheets/detail/nipah-virus |
| **Environmental suitability** | Pigott et al. *Unpublished.* "Mapping the environmental suitability for the Henipaviruses: Hendra and Nipah" |
| **Reservoir status determination** | Berger. (2005) "GIDEON: a comprehensive Web-based resource for geographic medicine" Int. J. Health Geogr. 4, 10 |
| **Human geopositioned occurrences** | Pigott et al. *Unpublished.* "Mapping the environmental suitability for the Henipaviruses: Hendra and Nipah" |

### Pathogen-specific Evidence

#### Plague

Overall assessment

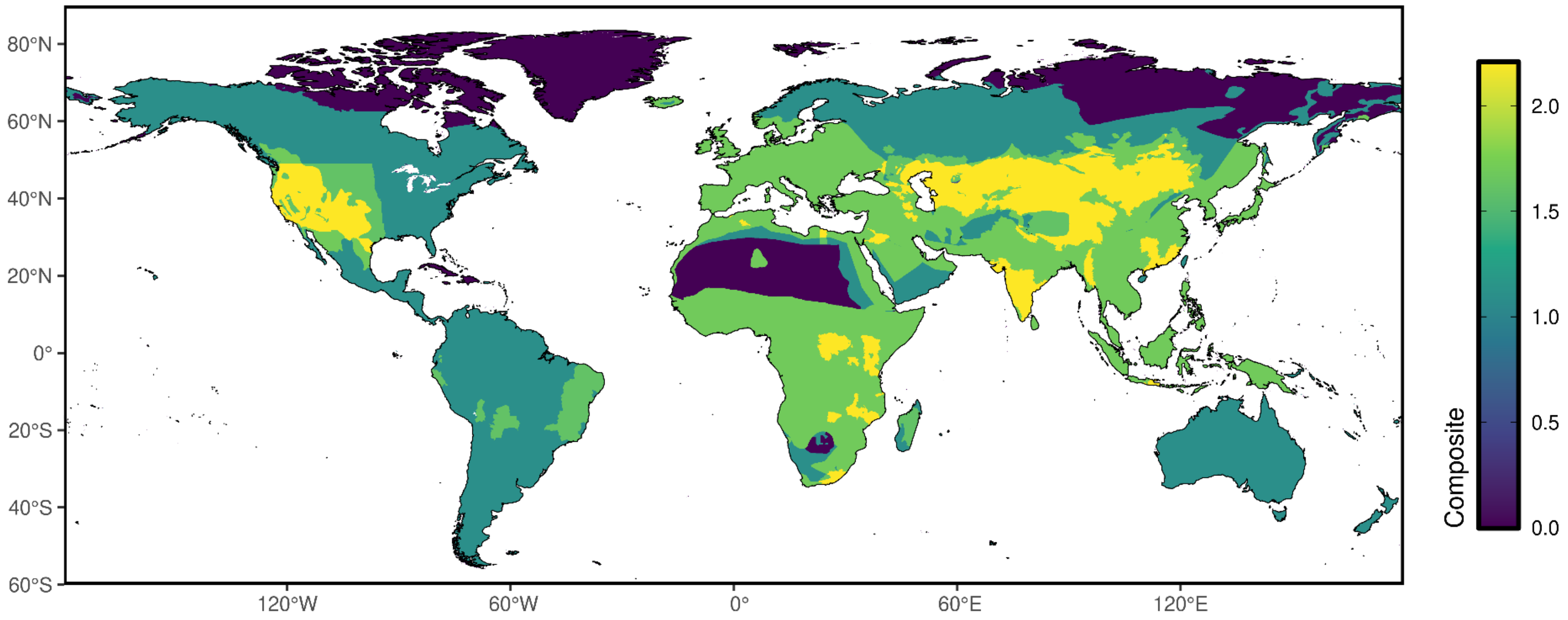

### Reservoir presence

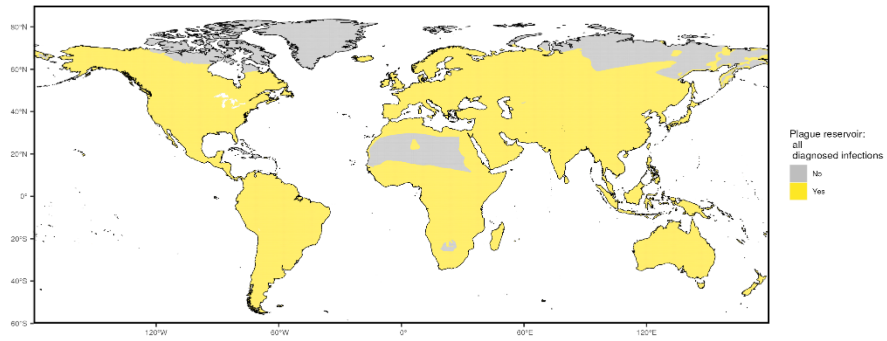


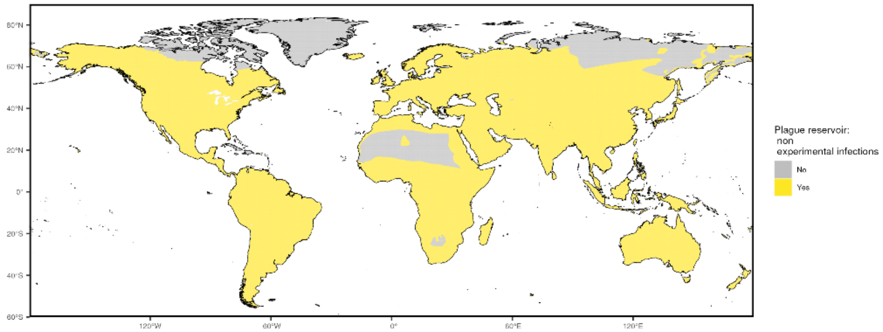


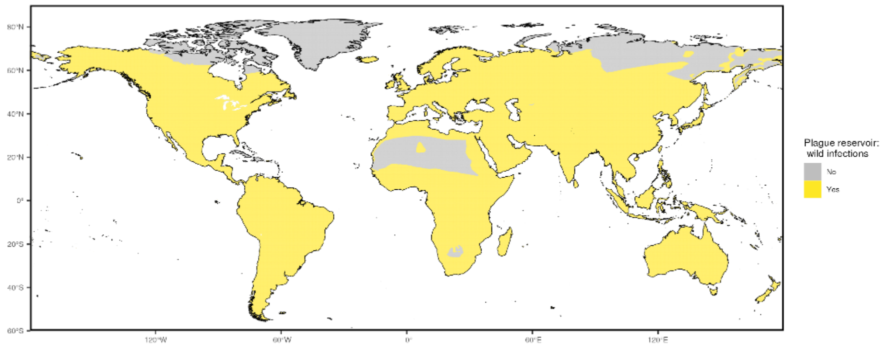


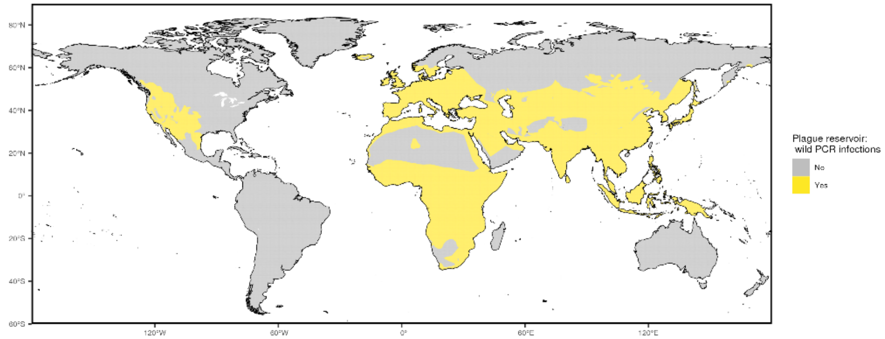

#### Prior cases

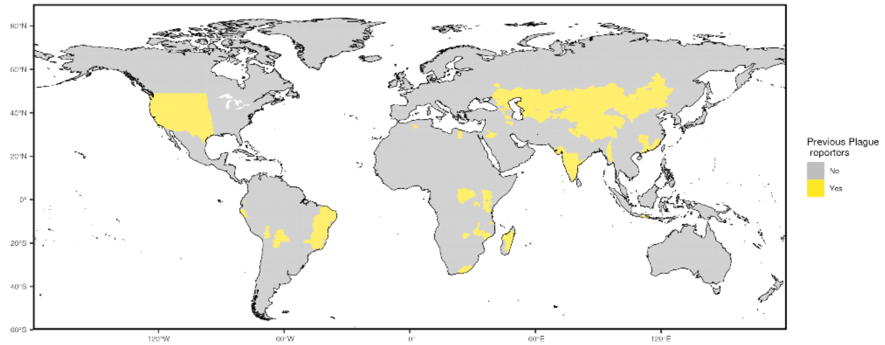


#### Precision weightings

| Precision categories | Precision weightings |
|---|---|
| Prior cases | 0.510473 |
| All diagnosed infections | 0.349955 |
| Non-experimental infections | 0.350536 |
| Wild infections | 0.391327 |
| Wild/PCR+ infections | 0.603233 |

NOTE: Precision weights here are the average of precision weighting for each category across pathogens with occurrence data

#### Data sources

**Plague**
Evidence data for plague were acquired from two main sources. First, we obtained a list of locations with historic transmission reports of plague compiled by the WHO, recorded at the first and second administrative units. Second, non-human species with evidence of Plague as reported in GIDEON were used in creating a zoonotic transmission layer, and these species were stratified into four groupings: all reported species, all non-experimental reports, all wild infections, and all wild, PCR positive infections. The geographic range for each species was obtained from the International Union for Conservation of Nature's (IUCN) Red List of Threatened Species.

| Evidence category | Citation |
|---|---|
| **Countries with reported human cases** | World Health Organization internal database |
| **Reservoir status determination** | Berger. (2005) "GIDEON: a comprehensive Web-based resource for geographic medicine" Int. J. Health Geogr. 4, 10 |

### Pathogen-specific Evidence

#### Rift Valley Fever virus

Overall assessment

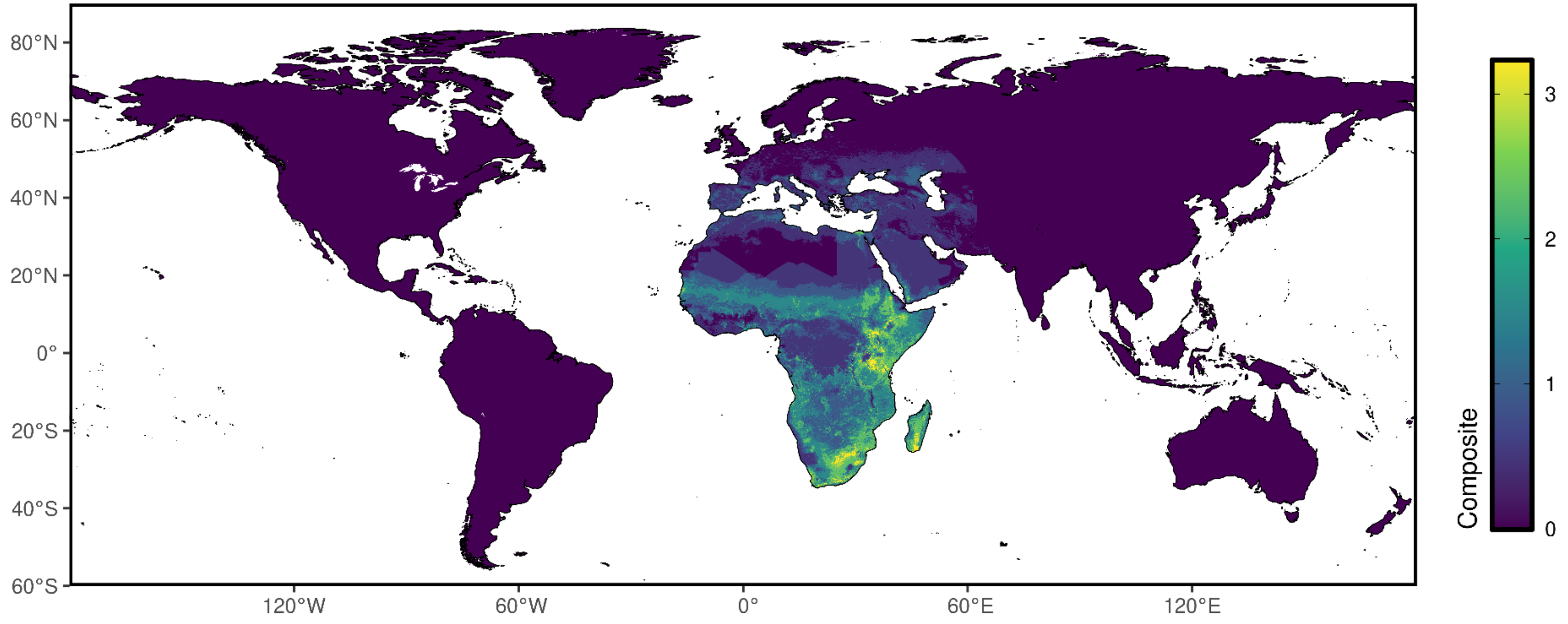

### Reservoir presence

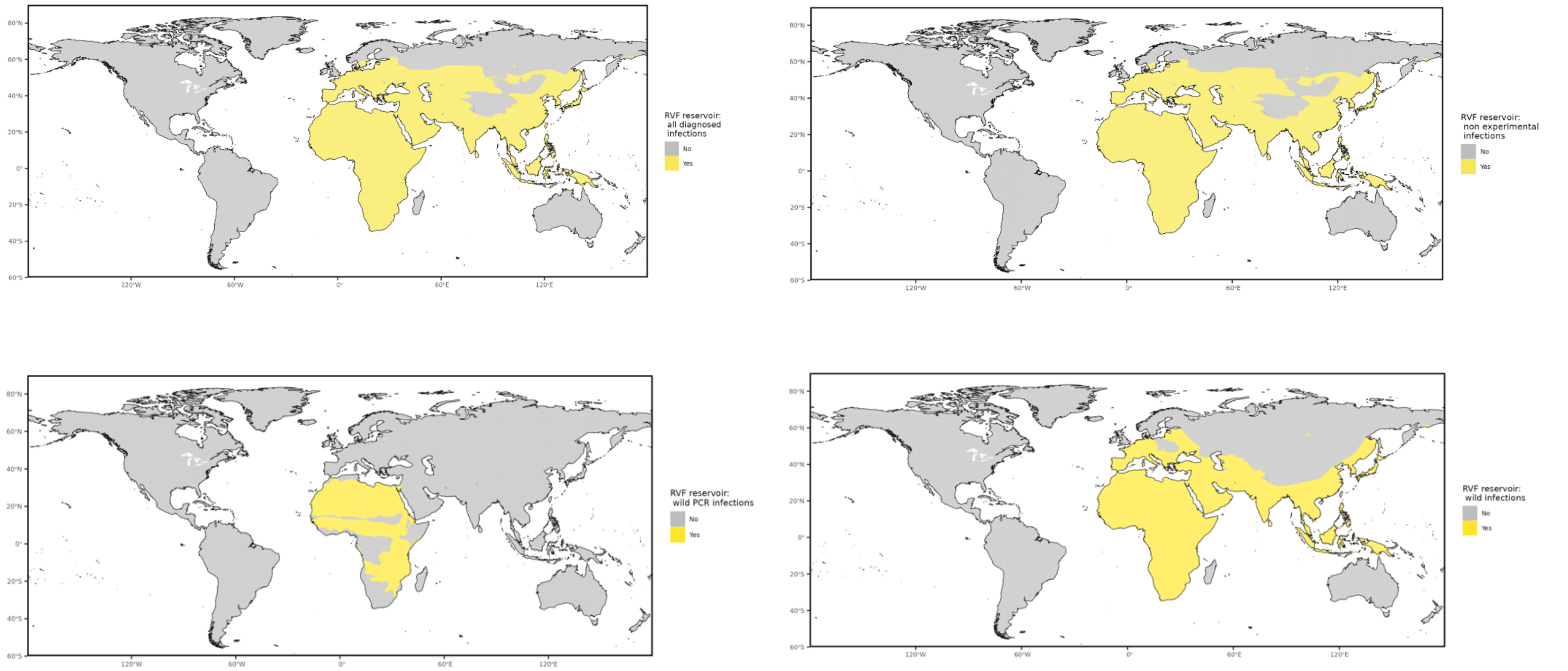

### Niche classifications

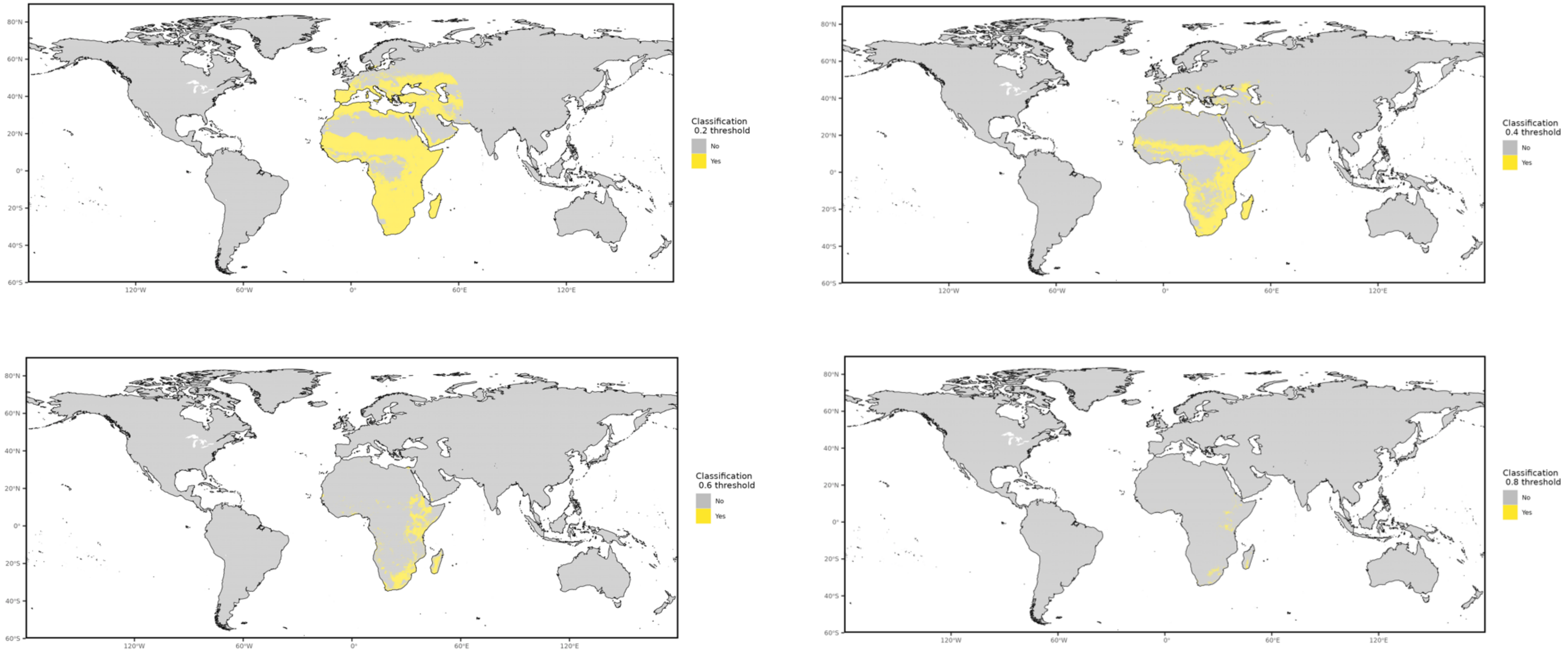

#### Prior cases

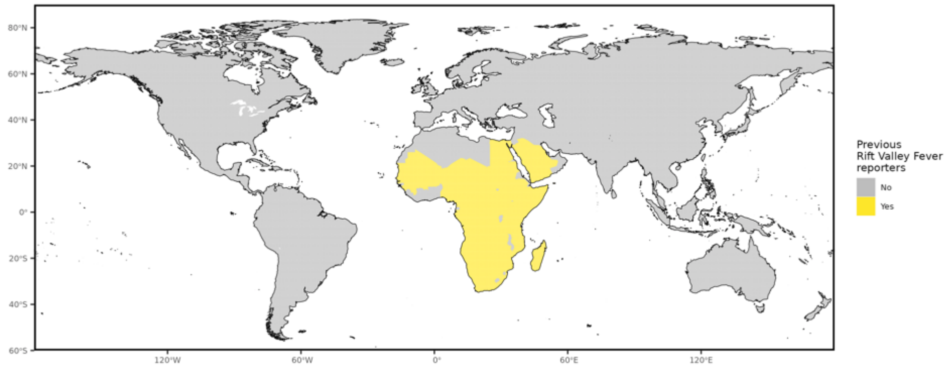


#### Occurrence data

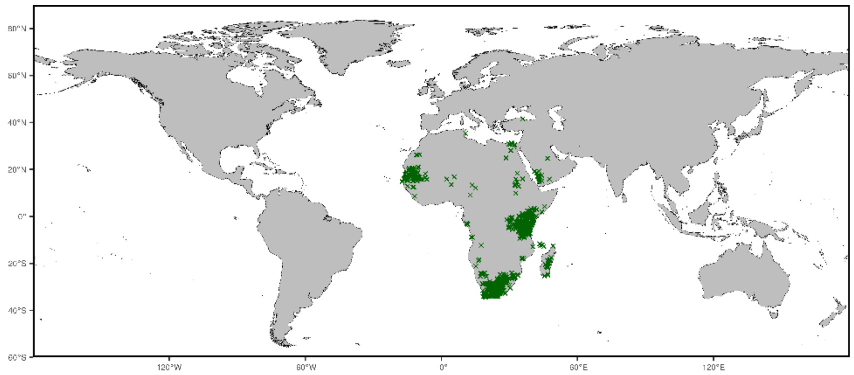


NOTE: centroids of polygons were used to map administrative and buffered occurrences, where relevant

### Precision weightings

| Precision categories | Precision weightings |
|---|---|
| Prior cases | 0.4676 |
| Niche classifications (0.2) | 0.4352 |
| Niche classifications (0.4) | 0.6026 |
| Niche classifications (0.6) | 0.8101 |
| Niche classifications (0.8) | 0.9129 |
| All diagnosed infections | 0.2442 |
| Non-experimental infections | 0.2442 |
| Wild infections | 0.2824 |
| Wild/PCR+ infections | 0.299 |

#### Data sources

Evidence data for Rift Valley Fever (RVF) disease were acquired from three main sources. First, we obtained a list of countries with historic transmission reports of RVF compiled by the CDC. Second, an environmental suitability synoptic niche map with continuous suitability estimates from Hardcastle et al. was used to create evidence suitability layers for RVF with cutoff scores of 0.2, 0.4, 0.6, and 0.8. This suitability layer was constructed using human and mammalian occurrence data and relevant environmental covariates (namely, monthly rainfall, including lags for one- and two-months prior rainfall, distance to closest floodplain, saturated water content of soil, bulk density of soil, monthly mean temperature, enhanced vegetation index, and the standard deviation of monthly precipitation over the years 1995 to 2018) to predict the estimated monthly environmental suitability for RVF. These twelve monthly estimates were then averaged to get the average suitability estimate over the entire year. Third, mammalian non-human species with evidence of Rift Valley Fever infections as reported to GIDEON were used were used in creating a zoonotic transmission layer, and these species were stratified into four groupings: all reported species, all non-experimental reports, all wild infections, and all wild, PCR positive infections. The geographic range for each species was obtained from the International Union for Conservation of Nature's (IUCN) Red List of Threatened Species.

Occurrence data for obtaining precision estimates from each layer were obtained from Hardcastle et al. (2020) and were collated using a comprehensive, systematic review and data extraction process.

| Evidence category | Citation |
|---|---|
| **Countries with reported human cases** | US Centers for Disease Control - https://www.cdc.gov/vhf/rvf/outbreaks/distribution-map.html |
| **Environmental suitability** | Hardcastle, A. N. (2020). Informing Rift Valley Fever preparedness by mapping seasonally varying environmental suitability. International journal of infectious diseases : IJID : official publication of the International Society for Infectious Diseases, 99, 362–372. https://doi.org/10.1016/j.ijid.2020.07.043 |
| **Reservoir status determination** | Berger. (2005) “GIDEON: a comprehensive Web-based resource for geographic medicine” Int. J. Health Geogr. 4, 10 |
| **Human geopositioned occurrences** | Hardcastle, A. N. (2020). Informing Rift Valley Fever preparedness by mapping seasonally varying environmental suitability. International journal of infectious diseases : IJID : official publication of the International Society for Infectious Diseases, 99, 362–372. https://doi.org/10.1016/j.ijid.2020.07.043 |

### Pathogen-specific Evidence

#### SARS-CoV-2

Overall assessment

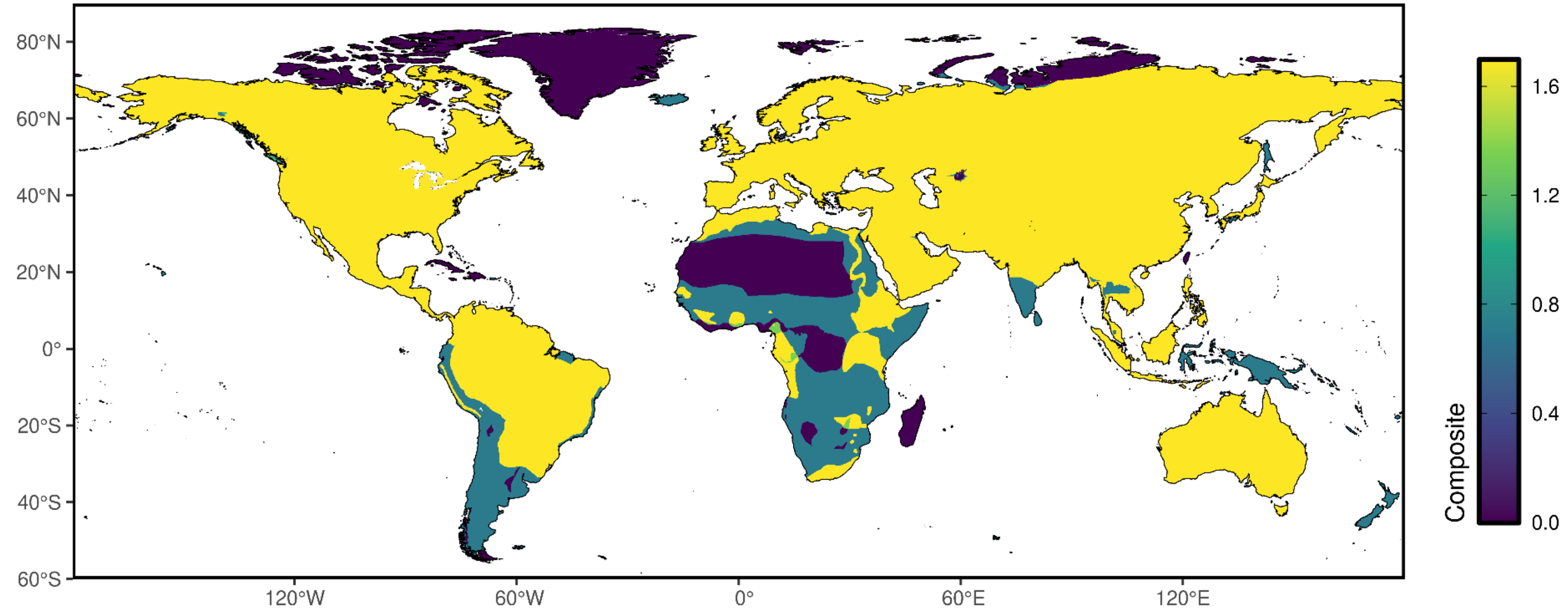

### Reservoir presence

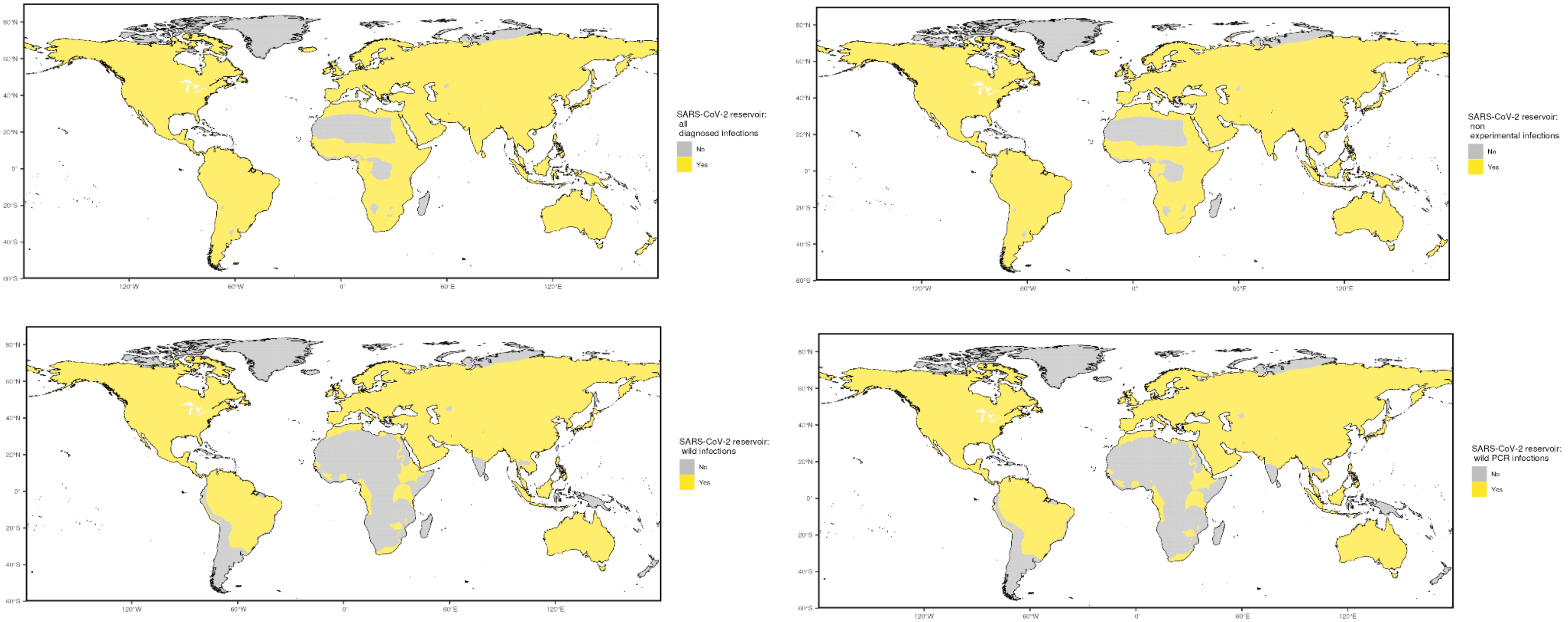

Precision weightings

| Precision categories | Precision weightings |
|---|---|
| All diagnosed infections | 0.349955 |
| Non-experimental infections | 0.350536 |
| Wild infections | 0.391327 |
| Wild/PCR+ infections | 0.603233 |

NOTE: Precision weights here are the average of precision weighting for each category across pathogens with occurrence data

#### Data sources

Evidence data for SARS-CoV-2 were acquired from two main sources. Data reported from FAO and WHO were used to compile a list of mammalian non-human species with evidence of SARS-CoV-2 used to create a zoonotic transmission layer. These species were stratified into three groupings: all reported species, all non-experimental reports, and all wild infections.
The geographic range for each mammalian species was obtained from the International Union for Conservation of Nature's (IUCN) Red List of Threatened Species.

| Evidence category | Citation |
|---|---|
| **Reservoir status determination** | 1. FAO . 2023. SARS-CoV-2 in animals situation update. Available at: https://www.fao.org/animal-health/situation-updates/sars-cov-2-in-animals/en. (Accessed 13 March 2023)<br>2. World Health Organization. 2023. [Internal Documentation] |

### Pathogen-specific Evidence

#### Yellow Fever virus

Overall assessment

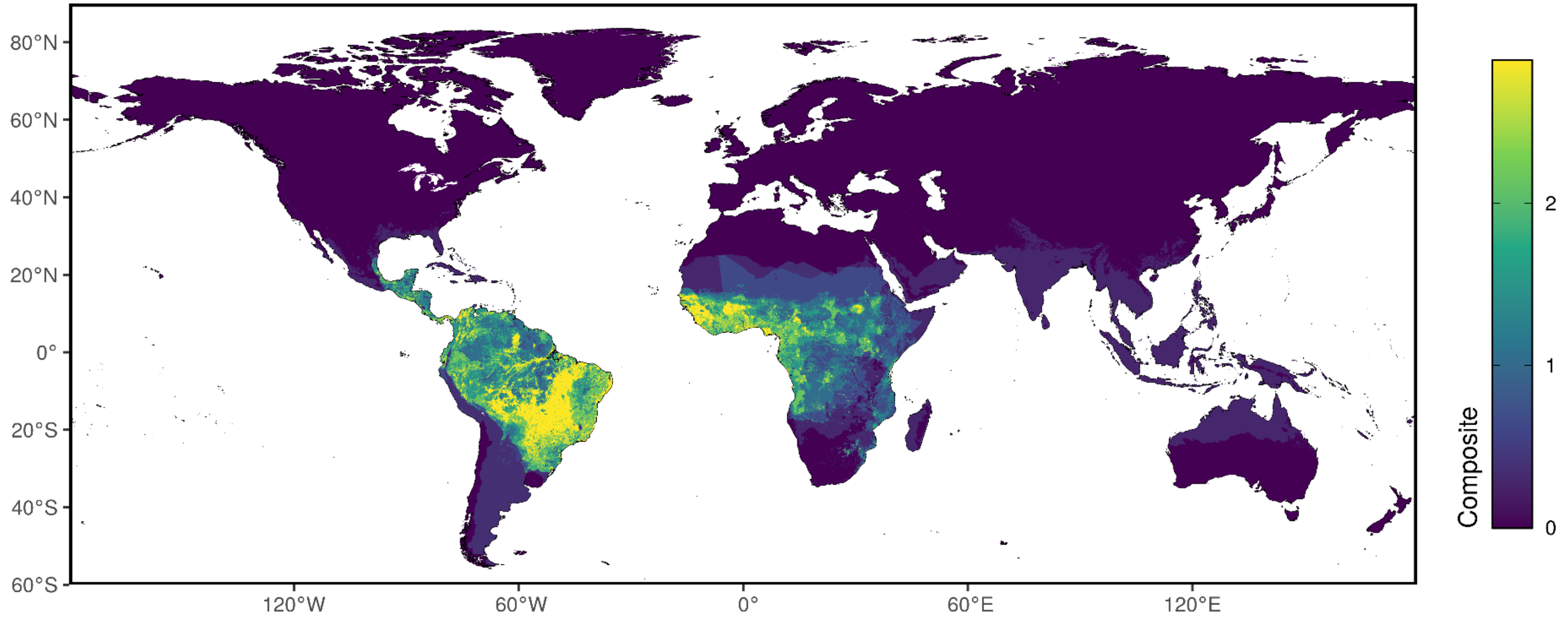

Aedes presence

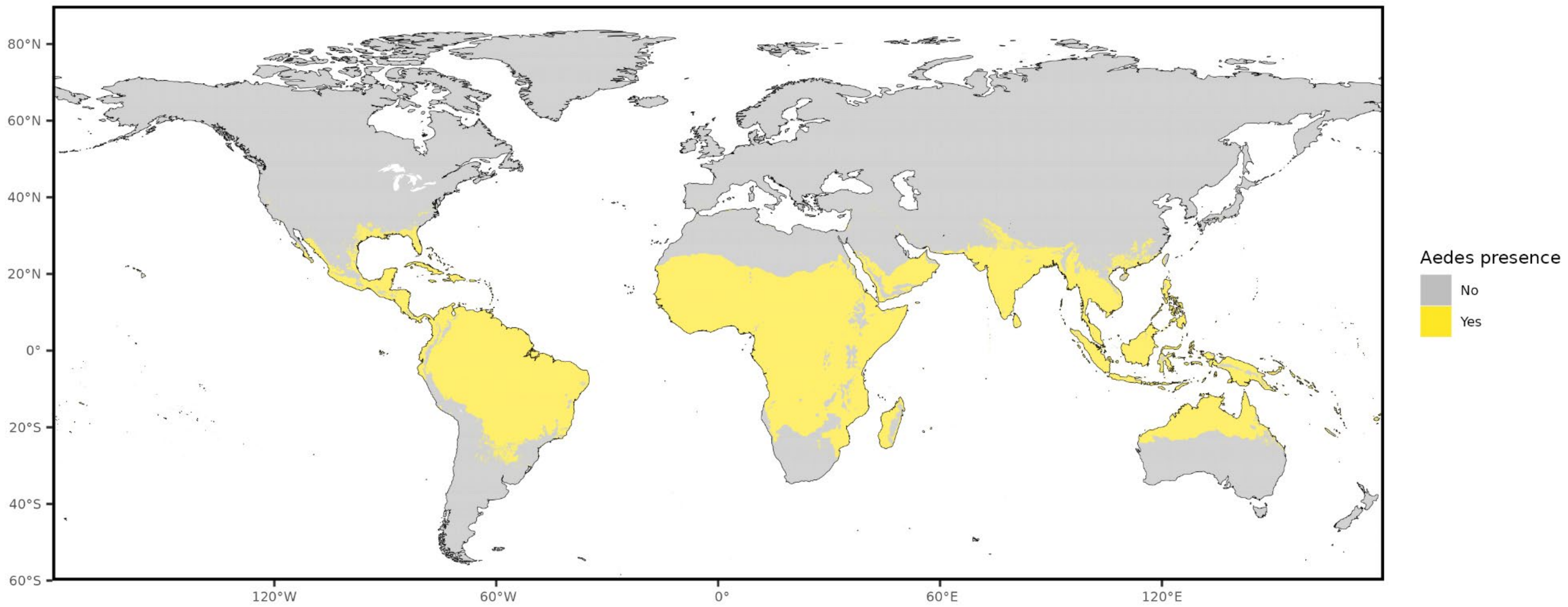

### Niche classifications

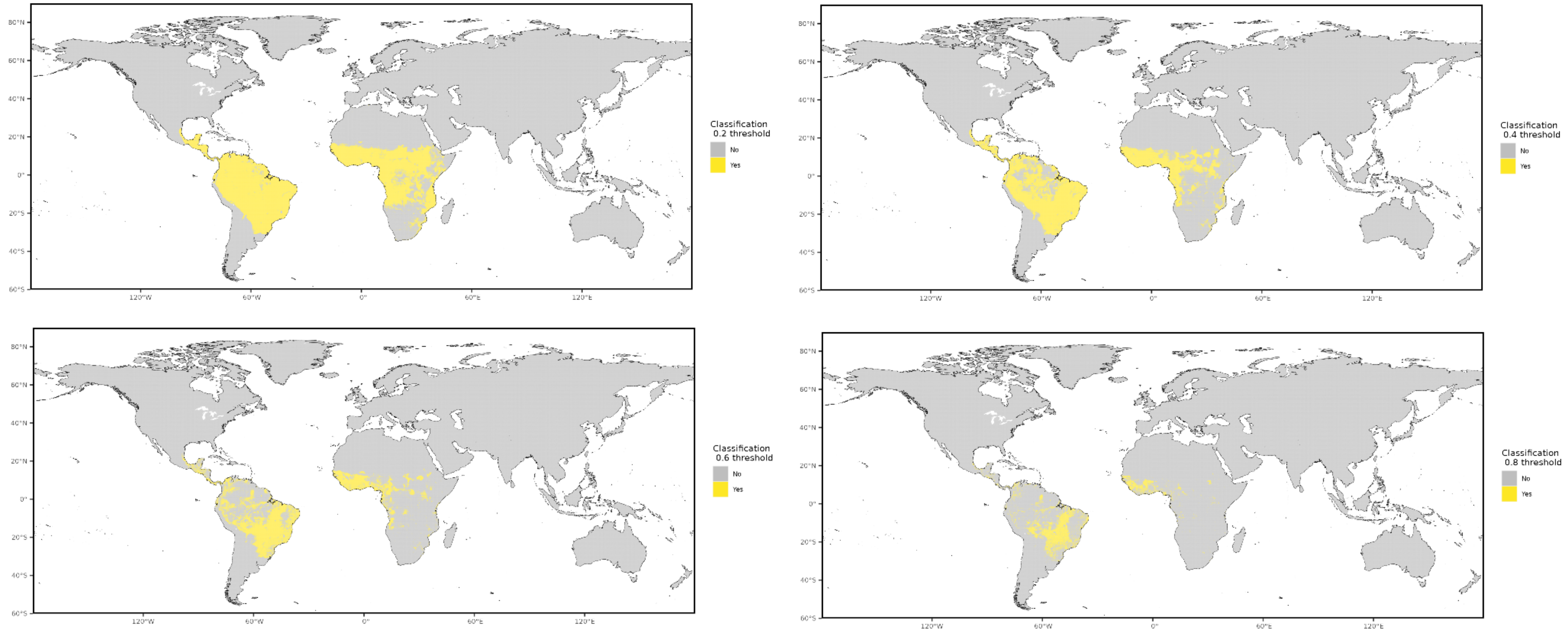

Prior cases

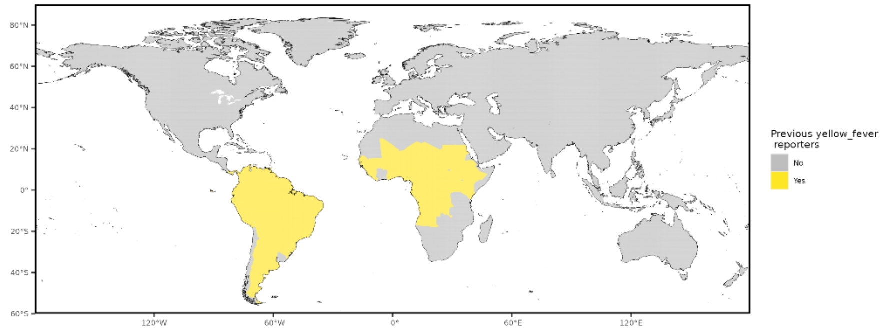


Occurrence data

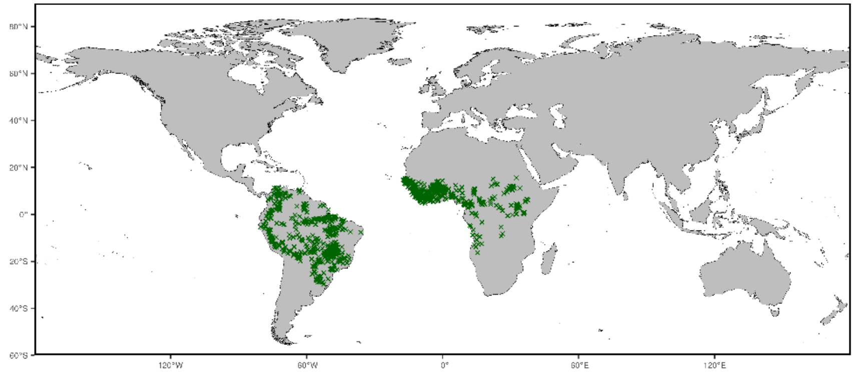


NOTE: centroids of polygons were used to map administrative and buffered occurrences, where relevant

Precision weightings

| Precision categories | Precision weightings |
|---|---|
| Aedes presence | 0.2712 |
| Prior cases | 0.3524 |
| Niche classifications (0.2) | 0.4080 |
| Niche classifications (0.4) | 0.4950 |
| Niche classifications (0.6) | 0.6004 |
| Niche classifications (0.8) | 0.7495 |

#### Data sources

Evidence data for Yellow Fever were acquired from three main sources. First, we obtained a list of countries with historic transmission reports of aedes-borne pathogens, subset specifically to Yellow Fever, compiled by the WHO. Second, an environmental suitability synoptic niche map with continuous suitability estimates from Lim et al. (2025) was used to create evidence suitability layers for Yellow Fever virus with cutoff scores of 0.2, 0.4, 0.6, and 0.8. Third, the global distribution of arbovirus vectors *Aedes aegypti* and *Aedes albopictus* as published by Kraemer et al. in 2015 were included as a vector layer.

Occurrence data for obtaining precision estimates from each layer were obtained from Lim et al. (2025)

| **Evidence category** | **Citation** |
|---|---|
| **Countries with reported human cases** | World Health Organization internal database |
| **Environmental suitability** | Lim, A., Shearer, F.M., Sewalk, K. *et al.* The overlapping global distribution of dengue, chikungunya, Zika and yellow fever. *Nat Commun* **16**, 3418 (2025). https://doi.org/10.1038/s41467-025-58609-5 |
| **Vector distribution** | Kraemer et al. (2015) "The global distribution of the arbovirus vectors Aedes aegypti and Ae. albopictus" eLife https://elifesciences.org/articles/08347 |
| **Human geopositioned occurrences** | Lim, A., Shearer, F.M., Sewalk, K. *et al.* The overlapping global distribution of dengue, chikungunya, Zika and yellow fever. *Nat Commun* **16**, 3418 (2025). https://doi.org/10.1038/s41467-025-58609-5 |

### Pathogen-specific Evidence

#### Zika virus

Overall assessment

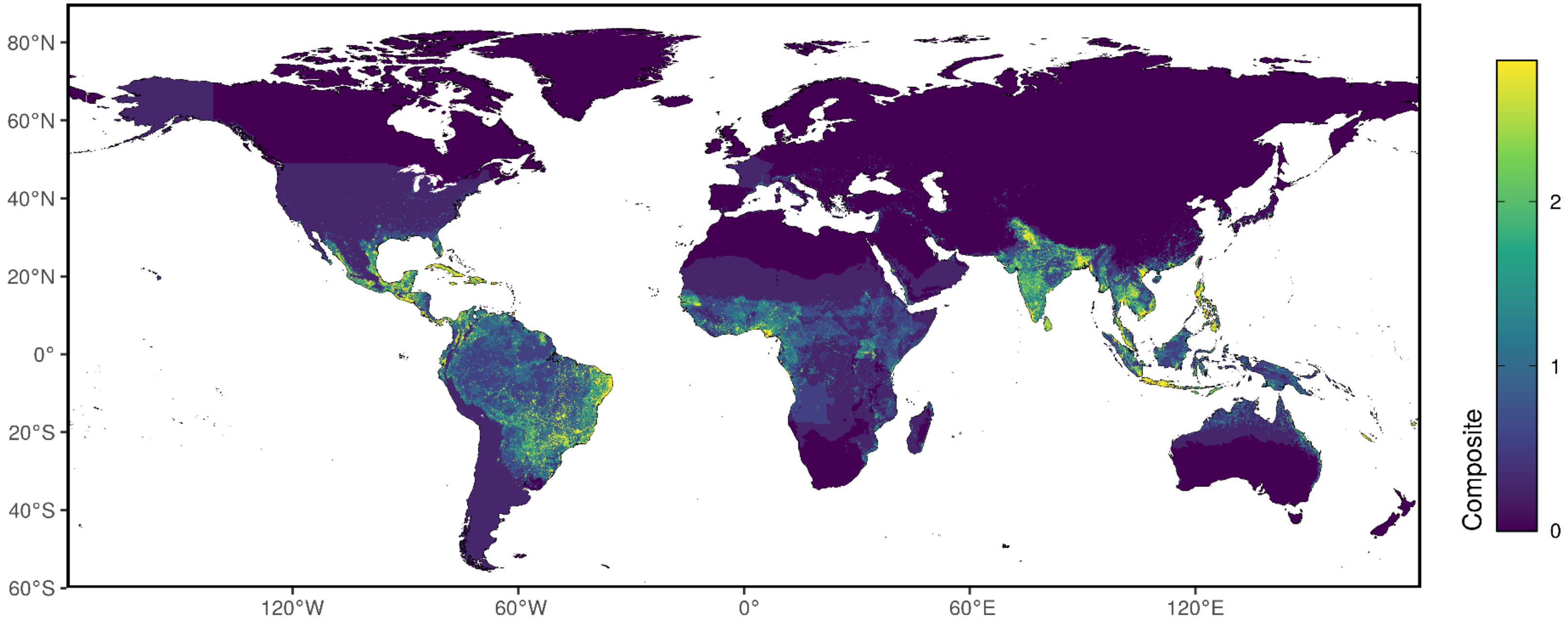

Aedes presence

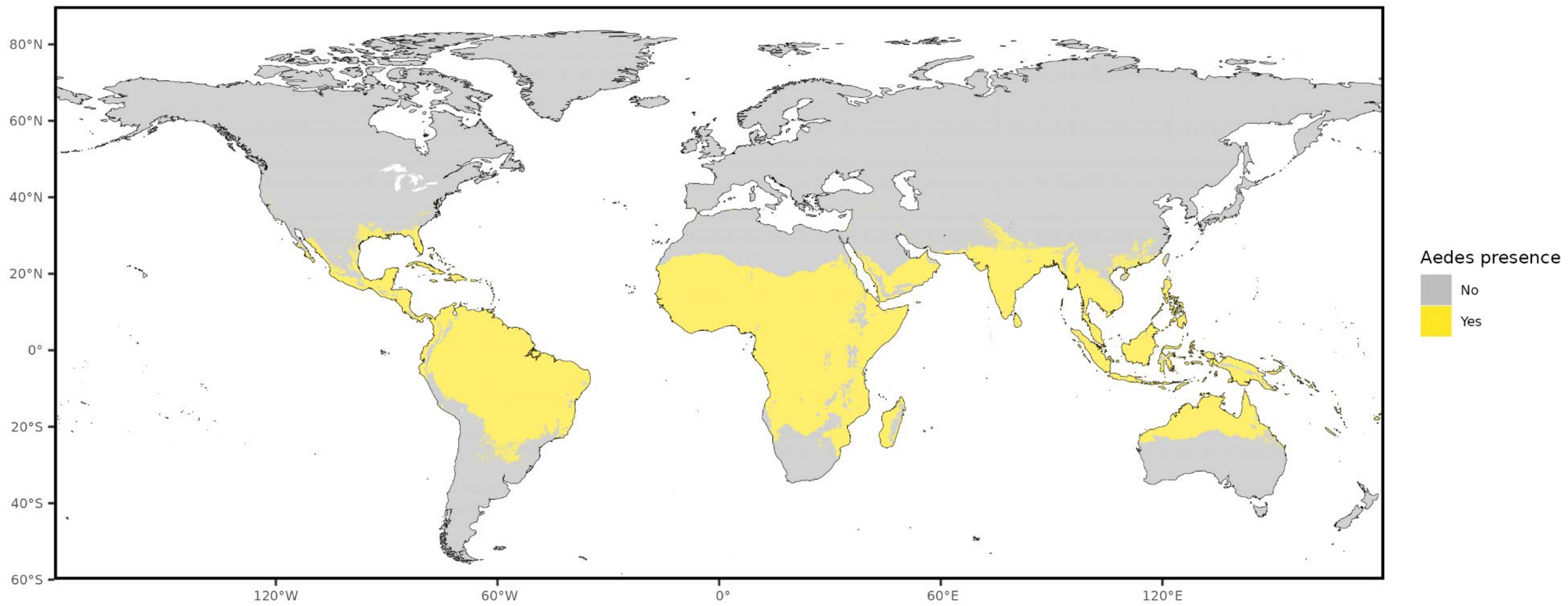

### Niche classifications

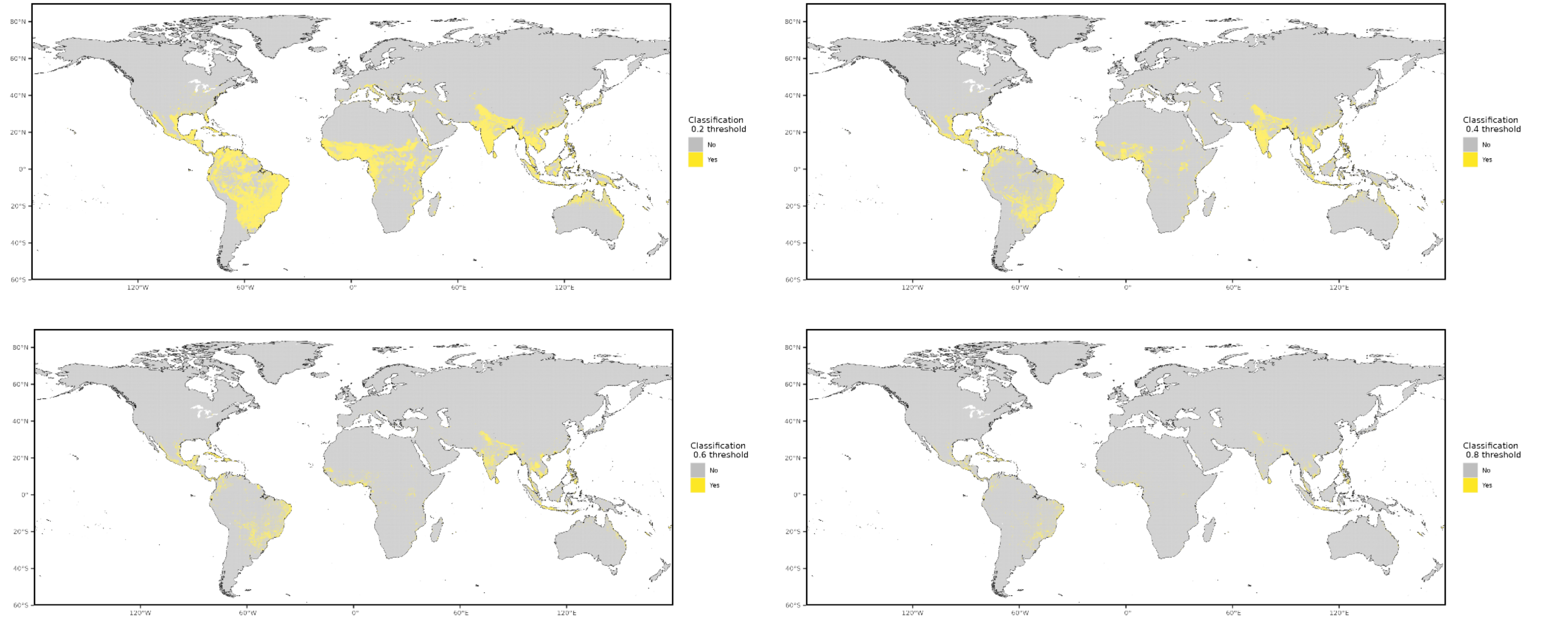

#### Prior cases

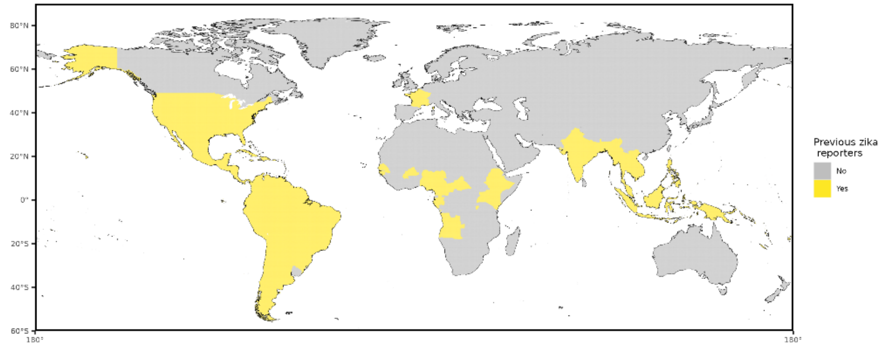


#### Occurrence data

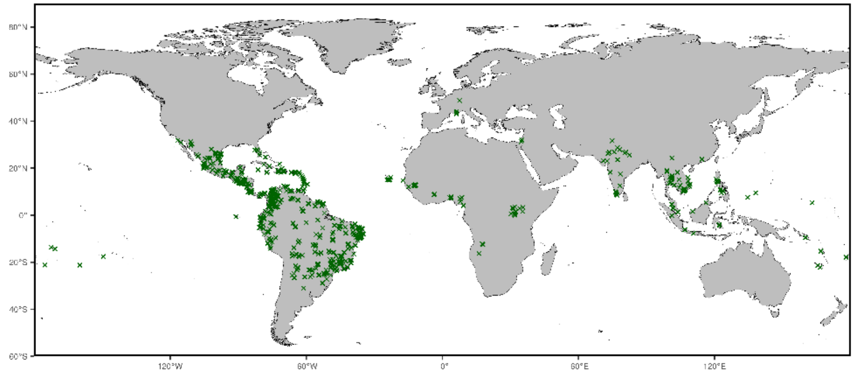


NOTE: centroids of polygons were used to map administrative and buffered occurrences, where relevant

Precision weightings

| **Precision categories** | **Precision weightings** |
|---|---|
| Aedes presence | 0.2537 |
| Prior cases | 0.2726 |
| Niche classifications (0.2) | 0.3662 |
| Niche classifications (0.4) | 0.5056 |
| Niche classifications (0.6) | 0.6468 |
| Niche classifications (0.8) | 0.8079 |

#### Data sources

Evidence data for Zika were acquired from three main sources. First, we obtained a list of countries with historic transmission reports of aedes-borne pathogens, subset specifically to Zika, compiled by the WHO. Second, an environmental suitability synoptic niche map with continuous suitability estimates from Lim et al. (2025) was used to create evidence suitability layers for Zika virus with cutoff scores of 0.2, 0.4, 0.6, and 0.8. Third, the global distribution of arbovirus vectors *Aedes aegypti* and *Aedes albopictus* as published by Kraemer et al. in 2015 were included as a vector layer.

Occurrence data for obtaining precision estimates from each layer were obtained from Lim et al. (2025).

| **Evidence category** | **Citation** |
|---|---|
| **Countries with reported human cases** | World Health Organization internal database |
| **Environmental suitability** | Lim, A., Shearer, F.M., Sewalk, K. *et al.* The overlapping global distribution of dengue, chikungunya, Zika and yellow fever. *Nat Commun* **16**, 3418 (2025). https://doi.org/10.1038/s41467-025-58609-5 |
| **Vector distribution** | Kraemer et al. (2015) "The global distribution of the arbovirus vectors Aedes aegypti and Ae. albopictus" eLife https://elifesciences.org/articles/08347 |
| **Human geopositioned occurrences** | Lim, A., Shearer, F.M., Sewalk, K. *et al.* The overlapping global distribution of dengue, chikungunya, Zika and yellow fever. *Nat Commun* **16**, 3418 (2025). https://doi.org/10.1038/s41467-025-58609-5 |
